# Incidence and prevalence of euthanasia in Belgium. A study using administrative data on all cases of euthanasia reported between 2002 and 2023

**DOI:** 10.1101/2024.10.16.24315619

**Authors:** Jacques Wels, Natasia Hamarat

## Abstract

**Background:** Reported assisted dying cases have increased in countries with such legislation. In Belgium, where euthanasia was legalized in 2002, cases rose from 236 in 2003 to 3,423 in 2023. While most studies focus on occurrence rates, this study examines the magnitude of increase and the contribution of demographic changes observed over the period.

**Methods:** We analysed complete data from the Belgian Federal Commission for the Control and Evaluation of Euthanasia (FCCEE) from 2002-2023 (N=33,604). Using Poisson regression, we calculated Rate Ratios (RR) by age, gender, region, and euthanasia characteristics. We compared estimates with a model that included demographic offsets to calculate Prevalence Rates (PR) and explored interaction effects across sub-categories.

**Results:** The yearly RR is 1.070, while the PR is 1.054, indicating that demographic changes significantly influence the observed increase. The PR for euthanasia among females has slightly risen (PR: 1.034), while psychiatric cases remained a small proportion (PR: 1.002). Cases citing multimorbidity have increased (RR: 1.029), whereas cases related to psychiatric disorders (PR: 0.949) and deaths in care homes (PR: 0.998) have not shown significant increases. Higher prevalence is observed in the Flemish region (PR: 1.983) but the gap has narrowed over the years.

**Interpretation:** A substantial part of the increase in euthanasia cases is attributed to demographic changes, particularly population ageing. Early increases were mainly due to the regulatory onset, while recent trends reflect a growing influence of demographic factors and regional adjustments. Considering demographic shifts is essential and long-term trends should be monitored.

**Funding:** ERC (UHealth), FNRS-CQ

## Background

The number of euthanasia cases has steadily increased in countries with regulations on euthanasia or assisted dying. In the Netherlands, cases rose from 1,933 in 2005 to 6,361 in 2019 ^1^. Euthanasia made up 1.9% of all deaths in 1990, increasing to 4.4% in 2017 ^1^. In Switzerland, between 1991 and 2008, the number of older women requesting assisted dying tripled, while it doubled for men ^2^. This rise is linked to more positive attitudes toward euthanasia ^3^, though opinions vary by country ^4^, and more nations are implementing or discussing assisted dying policies ^5^.

In Belgium, reported euthanasia cases rose from 235 in 2003 to 1,807 in 2013 ^6^, and to 2,700 in 2021 ^7^. Over 20 years, the number increased sevenfold, with euthanasia representing 2.4% of all deaths in 2021 ^7^. Belgium legalized euthanasia in May 2002 ^8^, becoming the second country to do so after the Netherlands. The law allows competent adults suffering from unbearable, untreatable physical or mental suffering due to a serious condition to request euthanasia. The request must be voluntary, written, and approved by independent doctors. In 2014, euthanasia was extended to minors – although concerning only a few cases ^9^ –, making Belgium the first country to allow it for children of any age. The Federal Commission for the Control and Evaluation of Euthanasia (FCCEE) oversees compliance and publishes regular reports to ensure transparency ^10^.

A common criticism of euthanasia is that allowing voluntary active euthanasia for some specific conditions would result in a so-called “slippery slope” ^11^. The “slippery slope” argument is a common criticism of voluntary active euthanasia, suggesting that once euthanasia is permitted for specific conditions, it may lead to broader, less ethically acceptable practices, such as for non-terminal or psychiatric conditions ^12,13^. This concern emphasises the need for rigorous safeguards, particularly in cases involving psychiatric disorders ^14^. Additionally, critics argue that socioeconomically vulnerable groups ^15^, including those in under-resourced healthcare settings ^16^ or deprived care facilities ^13^, could be disproportionately affected, raising concerns about potential coercion. Euthanasia, in this configuration, would not be efficiently monitored and controlled and might lead to error, abuse or violation of the rights of vulnerable patients ^17^. While the slippery slope argument often focuses on potential abuses, evidence from countries where assisted dying is legalized indicates that, although practices have expanded, they have largely remained within the strict legal and ethical frameworks established.^6^. Studies focusing on the slippery slope assumption rarely focus on data ^18^ and empirical investigation do not attest the existence of a slippery slope in the Netherlands ^19^ or Oregon ^20^.

Physical suffering without prospects of improvement was the most common reason given for granting euthanasia ^21^ and euthanasia is mostly linked to chronic or terminal physical conditions – with a large share due to cancers in terminal phase ^22^. Euthanasia for non-terminal illness is allowed in Belgium but the issue remains controversial and highly debated ^23^. It was estimated that, between 2002 and 2021, euthanasia for unbearable suffering caused by psychiatric disorders concerned at total 370 patients, 1.4 percent of the total number of euthanasia cases, most of which occurred after 2010. Research on medical files has found that most (90 percent) of these were diagnosed more than one disorder ^24^. Although concerning a small proportion of the causes of euthanasia in Belgium ^25^, psychiatric disorders are often identified to be a cause of concern ^14^ but data on such a population is sparse ^26^. The majority of the increase in cases in Belgium is particularly pronounced for those aged 80 and over, in a care home, those having a disease other than cancer and those not expected to die in the near future ^6^.

While much research and public reports indicate increasing trends in euthanasia among various population subgroups based on age, gender, or region of residence, they often overlook the demographic characteristics that underpin these trends. A public health approach to euthanasia should prioritize population-level data and propensities rather than focusing solely on individual cases ^27^. The aging population, regional distribution, and the higher proportion of women in older age groups can significantly influence the incidence of euthanasia cases. For instance, older adults may experience higher euthanasia rates due to the prevalence of terminal illnesses, while regional differences may reflect varying access to euthanasia services or differing cultural attitudes towards assisted dying ^4,28^. Ignoring these demographic characteristics could lead to misleading interpretations of euthanasia trends.

Despite the limited application of a demographic approach in this field, some studies have begun to explore euthanasia incidence among specific population subgroups. Gender disparities have been noted in the context of Belgian euthanasia data, which shows a relatively balanced distribution with females representing 49.6 percent of euthanasia cases in 2020 ^29^ and data on euthanasia as the ratio of all deaths by gender show similar rates among genders ^25^. Furthermore, when examining the conditions justifying euthanasia, females are notably overrepresented in psychiatric cases, although these comprise a small fraction of total cases ^30,31^. Regional differences have also been documented. For example, in the Netherlands, unexplained geographical variations in euthanasia incidence were observed across provinces. Factors such as age, church attendance, political orientation, income, self-perceived health, and availability of voluntary workers have been associated with these differences, yet a significant portion of the variation remains unexplained ^1^. In Belgium, official statistics reveal higher propensities for euthanasia in the Flemish region ^32^, with most research predominantly focused on Flanders ^33,34^.

Using Belgian administrative data on full euthanasia cases, this study has three key objectives. First, it seeks to provide prevalence rates of euthanasia categorized by age, gender, region, and health condition since the implementation of the regulation (2002-2023). Second, it aims to compare these prevalence figures with population characteristics and changes, offering incidence rates adjusted for demographic factors. Finally, the study intends to highlight changes observed over the period within specific population subgroups, contributing to a more nuanced understanding of euthanasia trends in Belgium.

## Data and methods

### Data

We use data routinely collected by the Federal Commission for the Control and Evaluation of Euthanasia (FCCEE), derived from individual reports that practitioners are legally required to submit for each case. These data are fully anonymized and encompass all reported euthanasia cases since 2002, including information on the reasons for euthanasia, as well as the patients’ gender, age group, and language. Additionally, we utilize open population data provided by Statbel, the Belgian Office for Statistics, which aggregates figures from administrative sources to provide population sizes by area of residence, gender, and age. The dataset includes 33,647 cases, representing all reported euthanasia cases in Belgium between 2002 and 2023. Since the euthanasia law was implemented in mid-2002, we exclude data from 2002 in our empirical models because the law was implemented in mid-2002 leading to low cases (N=24). Additionally, 43 cases were removed due to incomplete information. No imputations were made to address the missing data, given the small proportion (0.1% of the total) and limited available information. The final sample includes 33,623 cases.

### Euthanasia variables

The study focuses on nine variables available within the FCCEE dataset:

#### Reasons for Euthanasia

The Federal Commission for the Control and Evaluation of Euthanasia (FCCEE) identifies twelve possible medical conditions that can justify euthanasia. In this study, we classify reasons under seven categories: (1) cancer and tumours; (2) multimorbidity; (3) nervous system diseases; (4) specific diseases; (5) psychiatric disorders; (6) cognitive disorders; and (7) others. Specific diseases include diseases of the respiratory system, diseases of the circulatory system, diseases of the genitourinary system, diseases of the digestive system, haematological disorders, endocrine, nutritional, and metabolic diseases, diseases of the eye and its adnexa, diseases of the ear and mastoid process, diseases of the musculoskeletal system, muscles, and connective tissue, diseases of the skin and subcutaneous tissue. Other causes of euthanasia include symptoms, signs, and abnormal clinical and laboratory findings not elsewhere classified, traumatic injuries, poisoning, and certain other consequences of external causes, congenital malformations and chromosomal abnormalities, certain infectious and parasitic diseases. We maintain the FCCEE distinction between psychiatric and cognitive disorders due to their different clinical profiles. The tumours category serves as the reference.

#### Age Group

The FCCEE data include birth and death dates, but to preserve anonymity, eight post-calculated age groups are used: 15-29, 30-39, 40-49, 50-59 (reference category), 60-69, 70-79, 80-89, and 90+.

#### Gender

Gender, as reported by the medical practitioner, is recorded as male or female (reference category).

#### Language

Belgium’s federal structure includes three regions (Wallonia, Flanders, and Brussels). While place of residence was not consistently collected by the FCCEE until recently, language data (Dutch or French) used by the reporting medical practitioner are systematically included. This allows us to impute regional differences by distinguishing euthanasia cases reported in Dutch or French (reference category).

#### Year

The dataset records the year the euthanasia was carried out, ranging from 2003 to 2023 (we exclude 2002).

#### Basis for euthanasia

The dataset distinguishes between euthanasia requests made in advance (advanced request) or at the time of need (actual, reference category).

#### Type of suffering

The dataset includes information on the type of unbearable suffering reported by the practitioner, categorized as physical suffering (reference), mental suffering, or both.

#### Term of death

The dataset records whether the death was expected to occur within a year or over a longer period (reference category), as reported by the health care practitioner.

#### Place of death

The dataset distinguishes several types of places where the euthanasia was performed including home (reference), hospital, care home, palliative care and other.

### Population variables

We generate population figures based on demographic data retrieved from *Statbel*, the Belgian Statistical Office. They include information on the total population as of January 1st for each selected year (2003 to 2023), broken down by age group, sex, and region of residence. We chose to use population figures instead of the number of deaths (or, non-violent deaths), as done in previous studies ^1,21,25,35,36^, because a non-negligible share of euthanasia is performed on patients not expected to die in the foreseeable future, including those with dementia or psychiatric disorders – 14.4 percent of all cases in 2020-2021 ^29^. The figures are calculated for each line of euthanasia counts by year, age, gender, and language, and are then used as an offset in the model. Demographic data do not include information on language. To tackle this issue, the French-speaking population was calculated as the sum of the population residing in Wallonia and 90 percent of the population in Brussels and the Dutch speaking as the sum of the Flanders residents and 10 percent of the Brussels population, reflecting the Belgian language repartition. We additionally do <u>sensitivity</u> <u>analyses</u> using only Wallonia and Flanders and excluding Brussels. The values used for the demographic offset are shown in <u>supplementary file S1</u>.

### Analyses

We conducted a Poisson fixed effects analysis on the count data of euthanasia cases in Belgium, examining the effects of year, age group, gender, language, reason, basis, suffering and term of euthanasia. Since the study examines the total number of euthanasia cases over time – a count variable that has shown an increasing trend – we apply a Poisson model, which is specifically designed for non-negative integer outcomes and accounts for the mean-variance relationship inherent in such data ^37^, as done previously on suicide count data ^38,39^.

We compare two models. The first model does not include an offset for population size, and thus estimates the *occurrence* of euthanasia, providing insights into the raw counts without accounting for differences in population size. This model assumes that the observed counts are representative of the entire population, without adjusting for demographic variations, and is useful for identifying broad trends and associations. However, it may overlook the influence of population size and demographic patterns, potentially leading to biased interpretations. The second model includes an offset for population size by year, age, gender, and region, allowing us to calculate the *prevalence* of euthanasia, i.e., the rate of euthanasia occurrences relative to the population at risk. This approach offers a more accurate picture by normalizing the counts and providing insights into how euthanasia rates vary across different demographic groups. We exponentiate the coefficients to obtain the rate ratios (RR) and prevalence ratios (PR) ^40,41^. As our data include the total number of reported cases within the Belgian population with no possible sampling error, the analysis is not based on a sample, and significance levels are not required. The main article therefore does not show 95% confidence interval but they are included in the <u>supplementary files</u> for transparency.

We replicate the model without adjustment for demographic characteristics (unadjusted) and with full adjustment (age, gender, region). In Poisson regressions, offsets are used to account for exposure differences, such as population size, making it useful when analysing rates. However, adjusting for demographic characteristics is important even when an offset is used because the offset only adjusts for the size of the population at risk, not differences within the population that may influence the outcome. Additional analyses include multiplicative interaction terms between the years of data collection and all covariates, separately. Interaction terms are calculated using year as a numeric variable and marginal effects are calculated to plot change over time. Marginal effects plots are replicated using year as a categorical variable to address the non-linearity of time change (i.e., the fact that the number cases do not increase constantly in the same way across years). We additionally replicate the analyses using year as numeric but using two period-subsets (2002-2015; 2016-2023) to address the non-linearity of time change. Finally, we replicate the analysis using a demographic offset kept constant at baseline (year 2003) (PR_c_) to generate a counter-factual scenario in which demographics would have remained constant over the period to better highlight the contribution of demographic change in explaining prevalence ratios.

## Results

Table 1 exhibits the reported cases of euthanasia by demographic characteristics (year, age group, gender, language) as well as by reason, basis and term of euthanasia and type of suffering. The table include the number of reported cases as well as the percentage distribution by variable.

**Table 1.** Reported cases of euthanasia by year, age group, gender, language, reason, basis, suffering and term of euthanasia.

|  | Count | Percentage |  | Count | Percentage |
| --- | --- | --- | --- | --- | --- |
| **Year** |  |  | **Gender** |  |  |
| 2002 | 24 | 0.07 | Female | 16,711 | 49.67 |
| 2003 | 236 | 0.70 | Male | 16,902 | 50.23 |
| 2004 | 350 | 1.04 | NA | 34 | 0.10 |
| 2005 | 392 | 1.17 | **Language** |  |  |
| 2006 | 432 | 1.28 | FR (French) | 7,718 | 22.94 |
| 2007 | 496 | 1.47 | NL (Dutch) | 25,895 | 76.96 |
| 2008 | 706 | 2.10 | NA | 34 | 0.10 |
| 2009 | 822 | 2.44 | **Reason for euthanasia** |  |  |
| 2010 | 956 | 2.84 | Tumours | 21,919 | 65.14 |
| 2011 | 1,134 | 3.37 | Dementia | 310 | 0.92 |
| 2012 | 1,430 | 4.25 | Multimorbidity | 5,108 | 15.18 |
| 2013 | 1,815 | 5.39 | Nervous system diseases | 2,650 | 7.88 |
| 2014 | 1,933 | 5.74 | Others | 375 | 1.11 |
| 2015 | 2,023 | 6.01 | Psychiatric disorders | 427 | 1.27 |
| 2016 | 2,028 | 6.03 | Specific diseases | 2,832 | 8.42 |
| 2017 | 2,315 | 6.88 | NA | 26 | 0.08 |
| 2018 | 2,362 | 7.02 | **Basis for euthanasia** |  |  |
| 2019 | 2,658 | 7.90 | Actual | 33,169 | 98.58 |
| 2020 | 2,446 | 7.27 | Advanced | 444 | 1.32 |
| 2021 | 2,700 | 8.02 | **Type of suffering** |  |  |
| 2022 | 2,966 | 8.82 | Physical | 7,989 | 23.74 |
| 2023 | 3,423 | 10.17 | Both | 24,627 | 73.19 |
| **Age group** |  |  | Mental | 995 | 2.96 |
| 15-29 | 123 | 0.37 | NA | 36 | 0.11 |
| 30-39 | 401 | 1.19 | **Term of death** |  |  |
| 40-49 | 1,145 | 3.40 | >12 months | 4,913 | 14.60 |
| 50-59 | 3,422 | 10.17 | < 12 months | 28,697 | 85.29 |
| 60-69 | 7,015 | 20.85 | NA | 37 | 0.11 |
| 70-79 | 9,251 | 27.49 | **Place of death** |  |  |
| 80-89 | 9,063 | 26.94 | Home | 15,770 | 46.9 |
| 90+ | 3,184 | 9.46 | Hospital | 12,094 | 35.9 |
| NA | 43 | 0.13 | Care home | 4,352 | 12.9 |
|  |  |  | Palliative care | 778 | 2.31 |
|  |  |  | Other | 617 | 1.83 |
|  |  |  | NA | 36 | 0.11 |

The number of euthanasia increased yearly with, for instance, 24 cases reported in 2002, 1,430 in 2012 and 3,424 in 2023, correspondent to respectively 0.07, 4.25 and 10.17 percent of the total amount of euthanasia over the period. The increase is constant except in 2020, during the early stage of the COVID-19 pandemic, when the figure slightly dropped to 2,446 cases compared to 2,658 the previous year. Age groups 60-69, 70-79 and 80-89 show a higher number of cases, i.e., respectively 20.8, 27.5 and 26.9 percent of all euthanasia cases over the selected period. The number of cases observed within the population aged below 30 accounts for 0.4 percent of the total number of cases. The number of cases is quasi equally distributed amongst genders with 50.23 percent of cases observed among males. The presence of tumour(s) is the main cause of euthanasia, accounting for 65.1 percent of cases, just before multimorbidity that accounts for 15.2 percent of all cases. Euthanasia for psychiatric disorders and dementia ^42^ respectively account for 1.27 and 0.92 percent of all cases, 737 people in total. The basis for euthanasia is mostly actual and not made in advance with 98.6 percent of all cases not planned in advance. Health practitioners report suffering that is mostly both physical and mental (for 73.2 percent of cases), followed by physical only (23.7 percent) and mental only (2.96 percent of all cases). We also observe that in 85.3 percent of cases, the death is expected within the year. In 14.6 percent of cases, the death is not expected to occur within 12 months. Finally, a majority (46.9 percent) of euthanasia is made at home against 35.9 in hospital and 12.9 in case home.

Table 2 presents the rate ratio, prevalence ratio, and counterfactual prevalence ratio derived from the main model without interaction. The estimates indicate that the rate and prevalence ratios are similar in the unadjusted model (RR/PR=1.059) but higher in the counterfactual model (PR_c_=1.069), suggesting that demographic composition has increased the yearly rate of change by one percentage point. In the adjusted model, controlling for demographic characteristics, a difference of 1.6 between the RR (1.070) and PR (1.054) reveals that not accounting for demographics leads to an overestimation of euthanasia cases in Belgium. The counterfactual analysis confirms that without adjusting for demographic change, the prevalence and rate ratios remain similar. Additionally, the population aged 90+ is underrepresented in the RR (0.837) compared to the reference category (tumours), but euthanasia is far more prevalent in this group when adjusted for demographics (PR=13.186). This also affects gender distribution, where the male RR (1.046) contrasts with a higher PR (1.363), indicating higher prevalence among men. While previous studies reported higher euthanasia rates in the Flemish region, our analysis shows that moving from RR (2.451) to PR (1.512) moderates this finding, with euthanasia more prevalent in Flanders, though less so than earlier studies suggested (due to an aging population in the north).

**Table 2.**
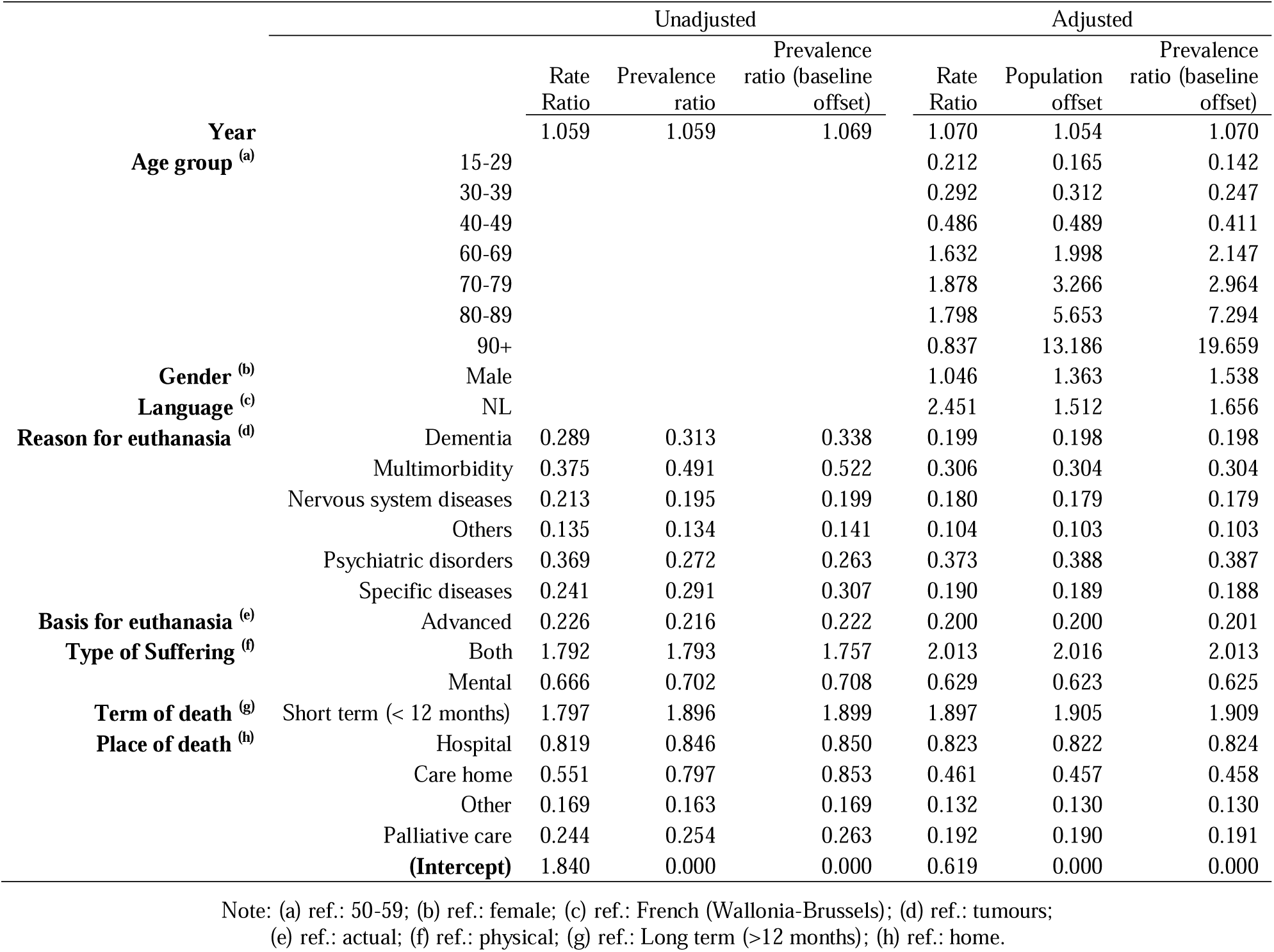
Rate ratio and prevalence ratio (fully adjusted model)

|  |  | Unadjusted |  |  | Adjusted |  |  |
| --- | --- | --- | --- | --- | --- | --- | --- |
|  |  | Rate Ratio | Prevalence ratio | Prevalence ratio (baseline offset) | Rate Ratio | Population offset | Prevalence ratio (baseline offset) |
| **Year** |  | 1.059 | 1.059 | 1.069 | 1.070 | 1.054 | 1.070 |
| **Age group <sup>(a)</sup>** | 15-29 |  |  |  | 0.212 | 0.165 | 0.142 |
|  | 30-39 |  |  |  | 0.292 | 0.312 | 0.247 |
|  | 40-49 |  |  |  | 0.486 | 0.489 | 0.411 |
|  | 60-69 |  |  |  | 1.632 | 1.998 | 2.147 |
|  | 70-79 |  |  |  | 1.878 | 3.266 | 2.964 |
|  | 80-89 |  |  |  | 1.798 | 5.653 | 7.294 |
|  | 90+ |  |  |  | 0.837 | 13.186 | 19.659 |
| **Gender <sup>(b)</sup>** | Male |  |  |  | 1.046 | 1.363 | 1.538 |
| **Language <sup>(c)</sup>** | NL |  |  |  | 2.451 | 1.512 | 1.656 |
| **Reason for euthanasia <sup>(d)</sup>** | Dementia | 0.289 | 0.313 | 0.338 | 0.199 | 0.198 | 0.198 |
|  | Multimorbidity | 0.375 | 0.491 | 0.522 | 0.306 | 0.304 | 0.304 |
|  | Nervous system diseases | 0.213 | 0.195 | 0.199 | 0.180 | 0.179 | 0.179 |
|  | Others | 0.135 | 0.134 | 0.141 | 0.104 | 0.103 | 0.103 |
|  | Psychiatric disorders | 0.369 | 0.272 | 0.263 | 0.373 | 0.388 | 0.387 |
|  | Specific diseases | 0.241 | 0.291 | 0.307 | 0.190 | 0.189 | 0.188 |
| **Basis for euthanasia <sup>(e)</sup>** | Advanced | 0.226 | 0.216 | 0.222 | 0.200 | 0.200 | 0.201 |
| **Type of Suffering <sup>(f)</sup>** | Both | 1.792 | 1.793 | 1.757 | 2.013 | 2.016 | 2.013 |
|  | Mental | 0.666 | 0.702 | 0.708 | 0.629 | 0.623 | 0.625 |
| **Term of death <sup>(g)</sup>** | Short term (< 12 months) | 1.797 | 1.896 | 1.899 | 1.897 | 1.905 | 1.909 |
| **Place of death <sup>(h)</sup>** | Hospital | 0.819 | 0.846 | 0.850 | 0.823 | 0.822 | 0.824 |
|  | Care home | 0.551 | 0.797 | 0.853 | 0.461 | 0.457 | 0.458 |
|  | Other | 0.169 | 0.163 | 0.169 | 0.132 | 0.130 | 0.130 |
|  | Palliative care | 0.244 | 0.254 | 0.263 | 0.192 | 0.190 | 0.191 |
|  | **(Intercept)** | 1.840 | 0.000 | 0.000 | 0.619 | 0.000 | 0.000 |

Note: (a) ref.: 50-59; (b) ref.: female; (c) ref.: French (Wallonia-Brussels); (d) ref.: tumours; (e) ref.: actual; (f) ref.: physical; (g) ref.: Long term (>12 months); (h) ref.: home.

Full results from the main model are shown in <u>supplementary files S2 (model 1) and S3</u> <u>(model 1)</u>. Sensitivity analyses using a region/language variable excluding Brussels shows little difference with, for instance, a PR of 1.054 for the variable year in the fully adjusted model, a difference of 0.001 units.

Figure 1 shows the marginal effect of years using it as a categorical variable to capture non-linear trends. In the unadjusted model, the rate ratio shows a steady increase over time, reflecting a consistent rise in the total number of euthanasia cases. The prevalence ratio, which accounts for population size, also increases but more gradually, suggesting that part of this trend can be explained by population growth. The prevalence ratio with a 2003 baseline, which holds demographic characteristics constant, remains lower than the standard prevalence ratio, indicating that demographic shifts, particularly population aging, have contributed to the rise in euthanasia prevalence. In the adjusted model, which controls for age, gender, and region, a notable pattern emerges. The three lines (rate ratio, prevalence ratio, and 2003 baseline prevalence ratio) overlap closely for the first 15 years, before diverging thereafter. This early overlap suggests that during the initial period, demographic changes did not strongly influence euthanasia rates. The divergence after year 15 indicates that population factors, such as aging, began playing a larger role in driving euthanasia prevalence. This shift may be explained by the time it took for euthanasia regulations to be fully implemented and for the practice to become more normalized. Around year 13 (2015), we might assume the end of the initial onset of the regulation, after which demographic shifts—such as the increasing proportion of elderly individuals—start to have a more pronounced impact on euthanasia trends. We replicated the analyses using year a linear variable on two subsets using 2015 as a cut point and distinguishing the onset period (2003-2015) and the recent period (2016-2023) in <u>supplementary file S6</u>. We observe that the RR for the year variable reaches 1.118 for the onset period against 1.048 in the recent period. Similarly, the PR is 1.100 in the first period and 1.033 in the second.

**Figure 1.**
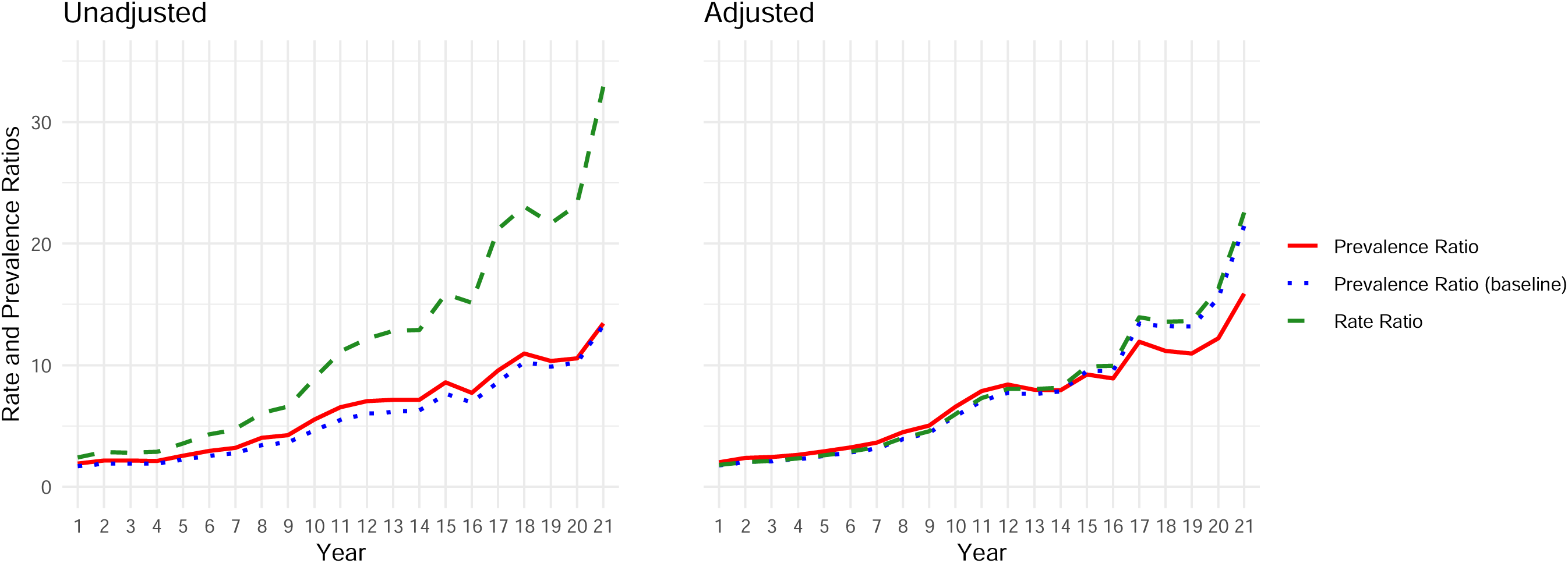
Predicted effect of year – prevalence ratio, rate ratio and sensitivity checks.

**Table 3** replicates the analysis with an interaction between year of data collection (numeric) and each variable, adjusting for all others. The table includes both the main effects and interaction terms (*Year). Results show relatively stable estimates for age groups, with constant rates for those under 40. However, PR has increased by 1.034 and 1.046 for the 70-79 and 80-89 age groups, respectively. Among men, prevalence slightly declined (PR, *Year=0.993), but counterfactual analysis suggests this is largely due to demographic changes (PR_c_, *Year=0.999). Prevalence in Flanders declined compared to Wallonia-Brussels, with counterfactual rates of 0.983 and 0.990, indicating minimal impact of demographic shifts. Regarding reasons for euthanasia, cases due to multimorbidity have increased compared to tumours (1.029), even when controlling for demographic changes (PR_c_: 1.032). In contrast, cases for dementia, psychiatric disorders, or advanced requests have not increased. No increase is seen in cases where death is expected beyond one year, with an interaction of PR=1.010 for short-term deaths. There is an increase in reporting both physical and mental suffering (PR=1.017), while a minor increase is seen for mental suffering alone (PR=1.002). Finally, there has been no increase in euthanasia cases in hospitals (PR=0.968) or care homes (PR=0.999), but a slight increase in palliative care settings (PR=1.018).

**Table 3.**
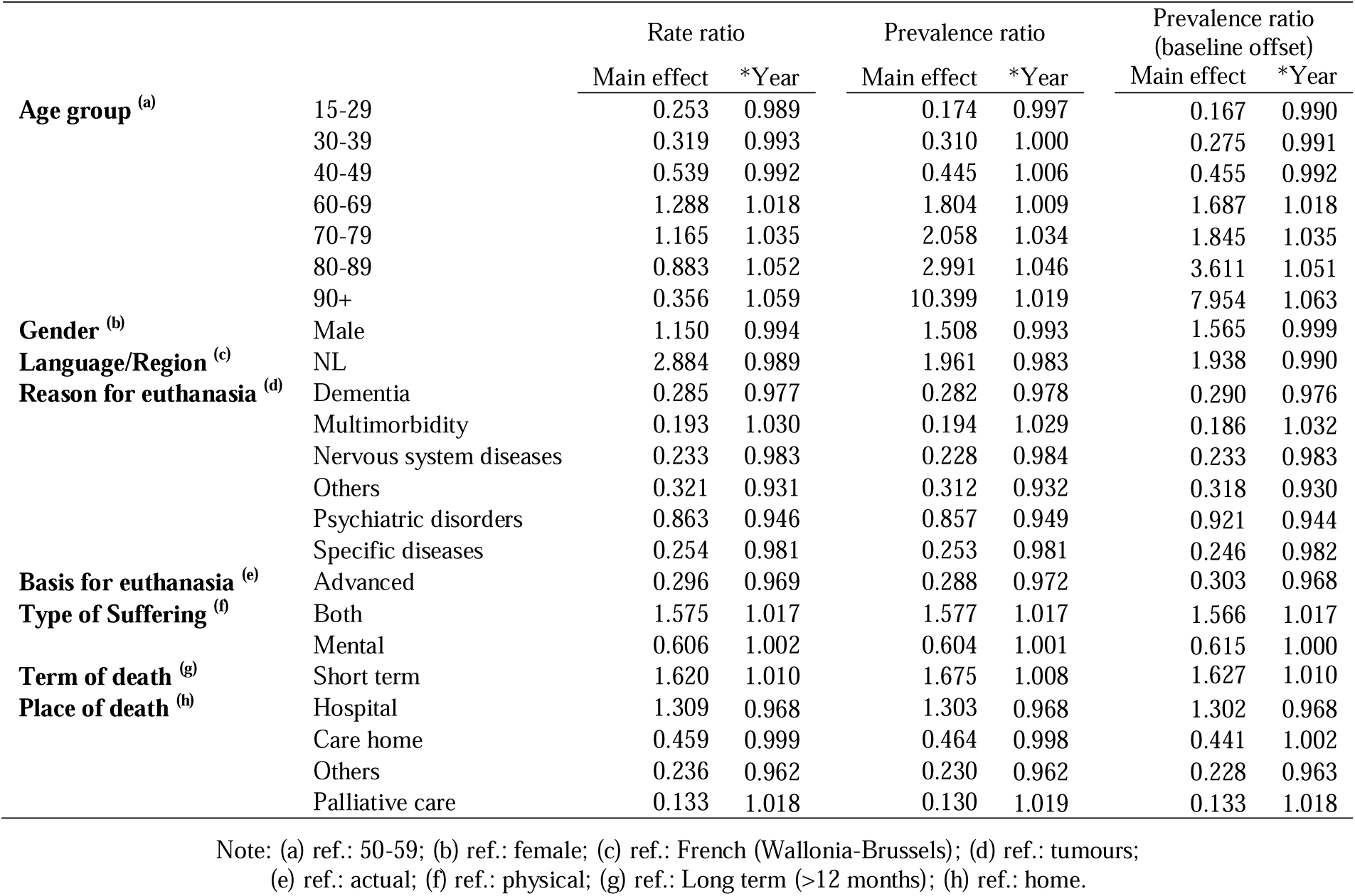
Interaction terms within the fully adjusted model.

Full results for the interaction effects are shown in supplementary <u>file S2 (models 2-6) and S3</u> <u>(models 2-9)</u>. Marginal effects plots calculated for each fully adjusted models are shown in <u>supplementary file S4 (using year as numeric) and S5 (using year as categorical)</u>.

## Discussion

Whilst euthanasia remains a controversial topic, more research is needed to thoroughly understand its prevalence in countries that have implemented it ^43^. This is particularly relevant as several nations are currently debating the potential introduction of assisted dying schemes, with cautionary perspectives often dominating the discourse over calls for much-needed legislation ^44,45^. The amount of discussion not based on scientific evidence ^46,47^ or on purely descriptive data ^48^ using the slippery slope as an assumption is problematic. With euthanasia figures increasing in all states that have implemented such type of legislation, a major question is understanding the demographic characteristics underlying incidence rates and to provide adjusted rates that accurately represent actual changes.

Using administrative data on all reported cases of euthanasia in Belgium since the legislation was implemented, this study confirms a significant rise in euthanasia cases, driven by both demographic changes and evolving societal attitudes. However, this increase requires careful interpretation. First, we find that approximately one-third to one-fourth of the overall rise during this period can be attributed to demographic changes. Second, while there was a steep increase in cases during the first ten years following the implementation of the regulation, the rate of increase slowed down after 2015, suggesting that it took time for both the Belgian population and healthcare practitioners to become familiar with and adopt the practice of euthanasia. As assisted dying policies are relatively recent in Belgium and other nations, it is crucial to account for a period of adjustment before the practice becomes normalised. Early trends might not reflect long-term trends. Third, we observe a long-term shift in euthanasia prevalence, marked by an increase among women and a reduction in regional differences between Dutch- and French-speaking areas. Finally, the rise in cases citing multimorbidity underscores the growing complexity of health conditions among those seeking euthanasia, whereas cases associated with psychiatric disorders and dementia have remained relatively stable since the implementation of the legislation.

This study has limitations. First, the FCCEE’s data collection methods, which exclude patient identifiers, limit our ability to link euthanasia cases with socioeconomic data. It is expected that patient identifiers allowing to link different sources of administrative data to address, for instance, socio-economic inequalities in euthanasia access will be available in the near future but not retroactively. This should be encouraged and we urge the FCCEE and the Belgian Government to facilitate data linkage on this matter. Second, the absence of information on patients’ regions of residence constrains more detailed regional analysis. Specifically, the data include information on patients’ language but information on region of residence is missing whilst there is no information on the area of residence. This, again, limits the ability to run analyses that would consider, for instance, a geographical approach to health inequalities. Third, by focusing solely on reported cases, we acknowledge that unreported instances of assisted dying may exist, potentially affecting the completeness of the data. Finally, since the dataset lacks information on the exact month of each euthanasia case, we were unable to account for a potential reduction in cases during the COVID-19 pandemic, which may have slightly biased the trends observed from 2020 onward. Nevertheless, our analyses provide no concrete evidence to support the notion of a ‘slippery slope’ in the Belgian context, where safeguards appear to function effectively. We recommend that future research focus on making better use of the available data and approach it with greater methodological rigor, rather than relying on crude descriptions that may not accurately reflect actual trends.

## Data Availability

Data access is granted upon request to the Belgian Federal Commission for the Control and Evaluation of Euthanasia (FCCEE), https://consultativebodies.health.belgium.be/en/advisory-and-consultative-bodies/federal-commission-control-and-evaluation-euthanasia

## Ethical statement

Access to data was granted by the ethic committee of the Federal Commission for the Control and Evaluation of Euthanasia (FCCEE) on the 14^th^ of May 2024. Data were used between the 1^st^ of July 2024 and the 14^th^ of October 2024.

## Data access

Data access is granted upon request to the FCCEE, https://consultativebodies.health.belgium.be/en/advisory-and-consultative-bodies/federal-commission-control-and-evaluation-euthanasia

## Authors contribution

Conceptualization: Jacques Wels, Natasia Hamarat; Data curation: Jacques Wels, Natasia Hamarat; Formal analysis: Jacques Wels; Funding acquisition: NA; Investigation: Jacques Wels, Natasia Hamarat; Methodology: Jacques Wels; Project administration: Jacques Wels, Natasia Hamarat; Resources: Jacques Wels, Natasia Hamarat; Software: Jacques Wels; Supervision: Jacques Wels; Validation: Natasia Hamarat; Visualization: Jacques Wels; Writing – original draft: Jacques Wels; Writing – review & editing: Natasia Hamarat

## Supplementary files

**Supplementary file S1. Population count on the 1st of January of years 2002-2023 by language, sex and age-group**

**Supplementary file S2. Poisson regression, not adjusted for demographic characteristics**

Model 1. Main model

Model 2. Interaction between year and reason for euthanasia

Model 3. Interaction between year and basis for euthanasia

Model 4. Interaction between year and type of suffering

Model 5. Interaction between year and expected term of death

Model 6. Interaction between year and place of death

**Supplementary file S3. Poisson regression, adjusted for demographic characteristics**

Model 1. Main model

Model 2. Interaction between year and reason for euthanasia

Model 3. Interaction between year and basis for euthanasia

Model 4. Interaction between year and type of suffering

Model 5. Interaction between year and expected term of death

Model 6. Interaction between year and place of death

Model 7. Interaction between year and age group

Model 8. Interaction between year and gender

Model 9. Interaction between year and language/region

**Supplementary file S4. Marginal effects in the fully adjusted model**

Model 1. Marginal effects of year

Model 2. Marginal effects of the interaction between age group and year

Model 3. Marginal effects of the interaction between gender and year

Model 4. Marginal effects of the interaction between language/region and year

Model 5. Marginal effects of the interaction between reason for euthanasia and year

Model 6. Marginal effects of the interaction between basis of euthanasia and year

Model 7. Marginal effects of the interaction between type of suffering and year

Model 8. Marginal effects of the interaction between term of death and year

Model 9. Marginal effects of the interaction between place of death and year

**Supplementary file S5. Marginal effects (year as categorical)**

Model 1. Marginal effects of year

Model 2. Marginal effects of the interaction between age group and year

Model 3. Marginal effects of the interaction between gender and year

Model 4. Marginal effects of the interaction between language/region and year

Model 5. Marginal effects of the interaction between reason for euthanasia and year

Model 6. Marginal effects of the interaction between basis of euthanasia and year

Model 7. Marginal effects of the interaction between type of suffering and year

Model 8. Marginal effects of the interaction between term of death and year

Model 9. Marginal effects of the interaction between place of death and year

**Supplementary file S6. Linear trends of year by time period (2003-2015; 2016-2022), fully adjusted model without interaction**

