## Supplementary files S1-S6 for "Incidence and prevalence of euthanasia in Belgium. A study using administrative data on all cases of euthanasia reported between 2002 and 2023"

### Supplementary file S1. Population count on the 1^st^ of January of years 2002-2023 by language, sex and age-group

| Language | Gender | Age group |  | Year | Brussels included | Brussels excluded |  | Year | Brussels included | Brussels excluded |
| --- | --- | --- | --- | --- | --- | --- | --- | --- | --- | --- |
| FR | female | 15-29 |  | **2002** | 399,434 | 306,945 |  | **2014** | 440,861 | 330,067 |
| NL | female | 15-29 |  | **2002** | 541,731 | 531,454 |  | **2014** | 566,178 | 553,868 |
| FR | female | 30-39 |  | **2002** | 312,059 | 241,998 |  | **2014** | 310,041 | 221,870 |
| NL | female | 30-39 |  | **2002** | 453,726 | 445,941 |  | **2014** | 411,256 | 401,459 |
| FR | female | 40-49 |  | **2002** | 308,436 | 248,853 |  | **2014** | 321,780 | 251,412 |
| NL | female | 40-49 |  | **2002** | 450,034 | 443,414 |  | **2014** | 458,862 | 451,043 |
| FR | female | 50-59 |  | **2002** | 257,702 | 207,844 |  | **2014** | 310,225 | 250,802 |
| NL | female | 50-59 |  | **2002** | 373,668 | 368,128 |  | **2014** | 462,914 | 456,311 |
| FR | female | 60-69 |  | **2002** | 202,791 | 163,746 |  | **2014** | 256,771 | 211,232 |
| NL | female | 60-69 |  | **2002** | 321,237 | 316,899 |  | **2014** | 374,373 | 369,313 |
| FR | female | 70-79 |  | **2002** | 207,283 | 166,338 |  | **2014** | 168,883 | 136,998 |
| NL | female | 70-79 |  | **2002** | 284,023 | 279,474 |  | **2014** | 281,269 | 277,726 |
| FR | female | 80-89 |  | **2002** | 98,347 | 75,437 |  | **2014** | 127,342 | 103,141 |
| NL | female | 80-89 |  | **2002** | 126,474 | 123,928 |  | **2014** | 193,749 | 191,060 |
| FR | female | 90+ |  | **2002** | 20,841 | 15,137 |  | **2014** | 28,098 | 21,549 |
| NL | female | 90+ |  | **2002** | 27,079 | 26,445 |  | **2014** | 37,774 | 37,046 |
| FR | male | 15-29 |  | **2002** | 407,269 | 317,578 |  | **2014** | 442,485 | 338,898 |
| NL | male | 15-29 |  | **2002** | 560,747 | 550,781 |  | **2014** | 579,511 | 568,001 |
| FR | male | 30-39 |  | **2002** | 319,100 | 244,472 |  | **2014** | 312,265 | 222,512 |
| NL | male | 30-39 |  | **2002** | 468,597 | 460,305 |  | **2014** | 416,679 | 406,706 |
| FR | male | 40-49 |  | **2002** | 307,122 | 248,814 |  | **2014** | 329,633 | 252,557 |
| NL | male | 40-49 |  | **2002** | 464,637 | 458,158 |  | **2014** | 471,849 | 463,285 |
| FR | male | 50-59 |  | **2002** | 251,055 | 204,457 |  | **2014** | 301,841 | 244,569 |
| NL | male | 50-59 |  | **2002** | 381,891 | 376,713 |  | **2014** | 471,215 | 464,851 |
| FR | male | 60-69 |  | **2002** | 176,655 | 143,328 |  | **2014** | 234,217 | 195,161 |
| NL | male | 60-69 |  | **2002** | 300,022 | 296,319 |  | **2014** | 367,283 | 362,943 |
| FR | male | 70-79 |  | **2002** | 141,457 | 115,104 |  | **2014** | 128,838 | 105,554 |
| NL | male | 70-79 |  | **2002** | 218,525 | 215,597 |  | **2014** | 238,964 | 236,377 |
| FR | male | 80-89 |  | **2002** | 43,610 | 33,673 |  | **2014** | 67,883 | 55,260 |
| NL | male | 80-89 |  | **2002** | 66,391 | 65,287 |  | **2014** | 120,435 | 119,032 |
| FR | male | 90+ |  | **2002** | 4,681 | 3,384 |  | **2014** | 8,520 | 6,450 |
| NL | male | 90+ |  | **2002** | 7,856 | 7,712 |  | **2014** | 14,089 | 13,859 |
| FR | female | 15-29 |  | **2003** | 399,733 | 305,687 |  | **2015** | 441,636 | 330,236 |
| NL | female | 15-29 |  | **2003** | 539,673 | 529,223 |  | **2015** | 566,157 | 553,779 |
| FR | female | 30-39 |  | **2003** | 312,554 | 240,967 |  | **2015** | 310,929 | 221,787 |
| NL | female | 30-39 |  | **2003** | 447,916 | 439,962 |  | **2015** | 413,496 | 403,591 |
| FR | female | 40-49 |  | **2003** | 310,509 | 250,321 |  | **2015** | 319,385 | 248,334 |
| NL | female | 40-49 |  | **2003** | 457,094 | 450,406 |  | **2015** | 449,627 | 441,732 |
| FR | female | 50-59 |  | **2003** | 267,213 | 215,990 |  | **2015** | 313,237 | 253,008 |
| NL | female | 50-59 |  | **2003** | 384,247 | 378,556 |  | **2015** | 469,902 | 463,210 |
| FR | female | 60-69 |  | **2003** | 197,625 | 159,222 |  | **2015** | 262,771 | 216,798 |
| NL | female | 60-69 |  | **2003** | 315,590 | 311,323 |  | **2015** | 380,802 | 375,694 |
| FR | female | 70-79 |  | **2003** | 205,480 | 165,380 |  | **2015** | 169,204 | 137,160 |
| NL | female | 70-79 |  | **2003** | 287,846 | 283,390 |  | **2015** | 282,881 | 279,321 |
| FR | female | 80-89 |  | **2003** | 102,107 | 78,789 |  | **2015** | 126,353 | 102,480 |
| NL | female | 80-89 |  | **2003** | 131,180 | 128,589 |  | **2015** | 196,917 | 194,264 |
| FR | female | 90+ |  | **2003** | 21,151 | 15,406 |  | **2015** | 29,713 | 22,889 |
| NL | female | 90+ |  | **2003** | 27,805 | 27,167 |  | **2015** | 40,683 | 39,925 |
| FR | male | 15-29 |  | **2003** | 407,180 | 316,364 |  | **2015** | 444,104 | 339,691 |
| NL | male | 15-29 |  | **2003** | 558,654 | 548,563 |  | **2015** | 579,699 | 568,098 |
| FR | male | 30-39 |  | **2003** | 320,259 | 243,375 |  | **2015** | 311,809 | 221,820 |
| NL | male | 30-39 |  | **2003** | 461,647 | 453,104 |  | **2015** | 417,500 | 407,501 |
| FR | male | 40-49 |  | **2003** | 309,497 | 249,813 |  | **2015** | 327,617 | 249,645 |
| NL | male | 40-49 |  | **2003** | 471,220 | 464,588 |  | **2015** | 461,680 | 453,016 |
| FR | male | 50-59 |  | **2003** | 260,342 | 212,523 |  | **2015** | 305,738 | 246,932 |
| NL | male | 50-59 |  | **2003** | 393,209 | 387,896 |  | **2015** | 478,687 | 472,153 |
| FR | male | 60-69 |  | **2003** | 172,827 | 139,966 |  | **2015** | 239,893 | 200,250 |
| NL | male | 60-69 |  | **2003** | 296,415 | 292,764 |  | **2015** | 373,706 | 369,301 |
| FR | male | 70-79 |  | **2003** | 141,432 | 115,331 |  | **2015** | 130,337 | 106,957 |
| NL | male | 70-79 |  | **2003** | 222,887 | 219,987 |  | **2015** | 242,562 | 239,964 |
| FR | male | 80-89 |  | **2003** | 46,305 | 35,967 |  | **2015** | 68,527 | 55,936 |
| NL | male | 80-89 |  | **2003** | 70,594 | 69,445 |  | **2015** | 124,424 | 123,025 |
| FR | male | 90+ |  | **2003** | 4,844 | 3,480 |  | **2015** | 9,269 | 7,063 |
| NL | male | 90+ |  | **2003** | 8,113 | 7,961 |  | **2015** | 15,473 | 15,228 |
| FR | female | 15-29 |  | **2004** | 401,945 | 306,687 |  | **2016** | 443,461 | 331,183 |
| NL | female | 15-29 |  | **2004** | 539,931 | 529,347 |  | **2016** | 567,415 | 554,940 |
| FR | female | 30-39 |  | **2004** | 311,257 | 238,822 |  | **2016** | 312,754 | 222,617 |
| NL | female | 30-39 |  | **2004** | 439,160 | 431,112 |  | **2016** | 416,499 | 406,484 |
| FR | female | 40-49 |  | **2004** | 313,177 | 252,455 |  | **2016** | 317,422 | 245,716 |
| NL | female | 40-49 |  | **2004** | 464,600 | 457,853 |  | **2016** | 441,489 | 433,522 |
| FR | female | 50-59 |  | **2004** | 274,492 | 222,523 |  | **2016** | 314,960 | 254,035 |
| NL | female | 50-59 |  | **2004** | 391,198 | 385,424 |  | **2016** | 473,796 | 467,027 |
| FR | female | 60-69 |  | **2004** | 196,366 | 157,846 |  | **2016** | 268,874 | 222,303 |
| NL | female | 60-69 |  | **2004** | 315,525 | 311,245 |  | **2016** | 388,343 | 383,168 |
| FR | female | 70-79 |  | **2004** | 202,075 | 163,107 |  | **2016** | 169,960 | 137,898 |
| NL | female | 70-79 |  | **2004** | 289,086 | 284,756 |  | **2016** | 285,038 | 281,476 |
| FR | female | 80-89 |  | **2004** | 105,700 | 81,937 |  | **2016** | 124,355 | 100,839 |
| NL | female | 80-89 |  | **2004** | 136,299 | 133,659 |  | **2016** | 198,628 | 196,015 |
| FR | female | 90+ |  | **2004** | 21,224 | 15,481 |  | **2016** | 30,787 | 23,916 |
| NL | female | 90+ |  | **2004** | 28,372 | 27,734 |  | **2016** | 42,489 | 41,726 |
| FR | male | 15-29 |  | **2004** | 408,377 | 317,123 |  | **2016** | 446,973 | 341,010 |
| NL | male | 15-29 |  | **2004** | 557,809 | 547,670 |  | **2016** | 581,899 | 570,125 |
| FR | male | 30-39 |  | **2004** | 319,018 | 241,450 |  | **2016** | 312,678 | 222,454 |
| NL | male | 30-39 |  | **2004** | 451,600 | 442,981 |  | **2016** | 419,570 | 409,545 |
| FR | male | 40-49 |  | **2004** | 311,838 | 251,337 |  | **2016** | 325,688 | 246,744 |
| NL | male | 40-49 |  | **2004** | 478,382 | 471,660 |  | **2016** | 452,695 | 443,923 |
| FR | male | 50-59 |  | **2004** | 267,531 | 219,160 |  | **2016** | 309,193 | 249,005 |
| NL | male | 50-59 |  | **2004** | 400,377 | 395,002 |  | **2016** | 483,608 | 476,920 |
| FR | male | 60-69 |  | **2004** | 172,419 | 139,398 |  | **2016** | 245,850 | 205,440 |
| NL | male | 60-69 |  | **2004** | 297,828 | 294,159 |  | **2016** | 381,327 | 376,837 |
| FR | male | 70-79 |  | **2004** | 140,484 | 114,839 |  | **2016** | 131,714 | 108,286 |
| NL | male | 70-79 |  | **2004** | 225,871 | 223,022 |  | **2016** | 246,139 | 243,536 |
| FR | male | 80-89 |  | **2004** | 48,770 | 38,070 |  | **2016** | 68,431 | 55,907 |
| NL | male | 80-89 |  | **2004** | 75,009 | 73,820 |  | **2016** | 127,426 | 126,034 |
| FR | male | 90+ |  | **2004** | 5,030 | 3,631 |  | **2016** | 9,797 | 7,542 |
| NL | male | 90+ |  | **2004** | 8,233 | 8,078 |  | **2016** | 16,680 | 16,429 |
| FR | female | 15-29 |  | **2005** | 405,356 | 308,637 |  | **2017** | 442,457 | 330,812 |
| NL | female | 15-29 |  | **2005** | 542,003 | 531,256 |  | **2017** | 567,099 | 554,694 |
| FR | female | 30-39 |  | **2005** | 308,505 | 235,750 |  | **2017** | 314,021 | 224,086 |
| NL | female | 30-39 |  | **2005** | 428,817 | 420,733 |  | **2017** | 419,255 | 409,262 |
| FR | female | 40-49 |  | **2005** | 315,745 | 254,414 |  | **2017** | 315,964 | 243,696 |
| NL | female | 40-49 |  | **2005** | 471,589 | 464,774 |  | **2017** | 435,640 | 427,610 |
| FR | female | 50-59 |  | **2005** | 281,193 | 228,808 |  | **2017** | 315,690 | 254,394 |
| NL | female | 50-59 |  | **2005** | 397,976 | 392,155 |  | **2017** | 476,227 | 469,416 |
| FR | female | 60-69 |  | **2005** | 196,684 | 157,888 |  | **2017** | 270,055 | 223,238 |
| NL | female | 60-69 |  | **2005** | 317,089 | 312,778 |  | **2017** | 392,860 | 387,658 |
| FR | female | 70-79 |  | **2005** | 198,718 | 160,902 |  | **2017** | 175,912 | 143,358 |
| NL | female | 70-79 |  | **2005** | 290,885 | 286,683 |  | **2017** | 290,869 | 287,252 |
| FR | female | 80-89 |  | **2005** | 109,487 | 85,336 |  | **2017** | 123,105 | 99,933 |
| NL | female | 80-89 |  | **2005** | 142,035 | 139,352 |  | **2017** | 200,625 | 198,050 |
| FR | female | 90+ |  | **2005** | 21,880 | 16,102 |  | **2017** | 31,706 | 24,737 |
| NL | female | 90+ |  | **2005** | 29,200 | 28,558 |  | **2017** | 44,853 | 44,079 |
| FR | male | 15-29 |  | **2005** | 410,365 | 318,797 |  | **2017** | 447,934 | 341,484 |
| NL | male | 15-29 |  | **2005** | 558,986 | 548,812 |  | **2017** | 583,015 | 571,187 |
| FR | male | 30-39 |  | **2005** | 315,616 | 238,166 |  | **2017** | 313,345 | 224,070 |
| NL | male | 30-39 |  | **2005** | 440,278 | 431,672 |  | **2017** | 421,843 | 411,924 |
| FR | male | 40-49 |  | **2005** | 314,995 | 253,319 |  | **2017** | 323,552 | 244,252 |
| NL | male | 40-49 |  | **2005** | 485,126 | 478,273 |  | **2017** | 445,915 | 437,104 |
| FR | male | 50-59 |  | **2005** | 273,561 | 224,879 |  | **2017** | 311,344 | 250,117 |
| NL | male | 50-59 |  | **2005** | 407,249 | 401,840 |  | **2017** | 487,053 | 480,250 |
| FR | male | 60-69 |  | **2005** | 173,413 | 140,225 |  | **2017** | 247,039 | 206,224 |
| NL | male | 60-69 |  | **2005** | 300,941 | 297,253 |  | **2017** | 385,365 | 380,830 |
| FR | male | 70-79 |  | **2005** | 139,333 | 114,074 |  | **2017** | 137,907 | 114,085 |
| NL | male | 70-79 |  | **2005** | 228,922 | 226,115 |  | **2017** | 253,603 | 250,956 |
| FR | male | 80-89 |  | **2005** | 51,528 | 40,381 |  | **2017** | 68,786 | 56,250 |
| NL | male | 80-89 |  | **2005** | 79,556 | 78,317 |  | **2017** | 130,778 | 129,385 |
| FR | male | 90+ |  | **2005** | 5,336 | 3,913 |  | **2017** | 10,159 | 7,895 |
| NL | male | 90+ |  | **2005** | 8,631 | 8,473 |  | **2017** | 17,862 | 17,610 |
| FR | female | 15-29 |  | **2006** | 410,716 | 311,863 |  | **2018** | 441,224 | 329,694 |
| NL | female | 15-29 |  | **2006** | 548,237 | 537,253 |  | **2018** | 566,867 | 554,475 |
| FR | female | 30-39 |  | **2006** | 306,897 | 233,281 |  | **2018** | 315,763 | 225,740 |
| NL | female | 30-39 |  | **2006** | 419,927 | 411,747 |  | **2018** | 421,129 | 411,126 |
| FR | female | 40-49 |  | **2006** | 317,585 | 255,622 |  | **2018** | 315,202 | 241,787 |
| NL | female | 40-49 |  | **2006** | 475,754 | 468,869 |  | **2018** | 432,982 | 424,825 |
| FR | female | 50-59 |  | **2006** | 288,139 | 235,010 |  | **2018** | 315,820 | 254,018 |
| NL | female | 50-59 |  | **2006** | 405,969 | 400,066 |  | **2018** | 476,545 | 469,678 |
| FR | female | 60-69 |  | **2006** | 197,600 | 158,814 |  | **2018** | 271,291 | 224,183 |
| NL | female | 60-69 |  | **2006** | 319,350 | 315,040 |  | **2018** | 398,444 | 393,210 |
| FR | female | 70-79 |  | **2006** | 194,878 | 157,890 |  | **2018** | 182,114 | 149,059 |
| NL | female | 70-79 |  | **2006** | 291,514 | 287,404 |  | **2018** | 295,568 | 291,895 |
| FR | female | 80-89 |  | **2006** | 113,872 | 89,319 |  | **2018** | 121,863 | 98,989 |
| NL | female | 80-89 |  | **2006** | 148,942 | 146,214 |  | **2018** | 202,879 | 200,337 |
| FR | female | 90+ |  | **2006** | 21,396 | 15,783 |  | **2018** | 32,426 | 25,392 |
| NL | female | 90+ |  | **2006** | 28,829 | 28,205 |  | **2018** | 46,656 | 45,874 |
| FR | male | 15-29 |  | **2006** | 414,501 | 321,844 |  | **2018** | 448,148 | 340,604 |
| NL | male | 15-29 |  | **2006** | 564,083 | 553,788 |  | **2018** | 584,294 | 572,345 |
| FR | male | 30-39 |  | **2006** | 312,894 | 234,834 |  | **2018** | 314,625 | 225,883 |
| NL | male | 30-39 |  | **2006** | 430,497 | 421,824 |  | **2018** | 423,522 | 413,662 |
| FR | male | 40-49 |  | **2006** | 317,906 | 254,771 |  | **2018** | 321,980 | 242,270 |
| NL | male | 40-49 |  | **2006** | 489,836 | 482,821 |  | **2018** | 441,496 | 432,639 |
| FR | male | 50-59 |  | **2006** | 280,044 | 230,615 |  | **2018** | 312,946 | 250,319 |
| NL | male | 50-59 |  | **2006** | 414,942 | 409,450 |  | **2018** | 488,047 | 481,088 |
| FR | male | 60-69 |  | **2006** | 174,830 | 141,618 |  | **2018** | 248,286 | 207,124 |
| NL | male | 60-69 |  | **2006** | 304,383 | 300,693 |  | **2018** | 391,631 | 387,057 |
| FR | male | 70-79 |  | **2006** | 137,571 | 112,679 |  | **2018** | 144,232 | 119,903 |
| NL | male | 70-79 |  | **2006** | 231,376 | 228,610 |  | **2018** | 259,368 | 256,665 |
| FR | male | 80-89 |  | **2006** | 54,293 | 42,795 |  | **2018** | 69,248 | 56,789 |
| NL | male | 80-89 |  | **2006** | 84,607 | 83,329 |  | **2018** | 134,399 | 133,015 |
| FR | male | 90+ |  | **2006** | 5,238 | 3,840 |  | **2018** | 10,481 | 8,165 |
| NL | male | 90+ |  | **2006** | 8,675 | 8,520 |  | **2018** | 18,856 | 18,599 |
| FR | female | 15-29 |  | **2007** | 416,313 | 316,117 |  | **2019** | 440,168 | 327,842 |
| NL | female | 15-29 |  | **2007** | 554,122 | 542,989 |  | **2019** | 566,257 | 553,776 |
| FR | female | 30-39 |  | **2007** | 305,860 | 231,257 |  | **2019** | 318,666 | 228,318 |
| NL | female | 30-39 |  | **2007** | 413,206 | 404,917 |  | **2019** | 422,981 | 412,942 |
| FR | female | 40-49 |  | **2007** | 318,667 | 256,196 |  | **2019** | 314,454 | 239,966 |
| NL | female | 40-49 |  | **2007** | 478,243 | 471,302 |  | **2019** | 431,684 | 423,408 |
| FR | female | 50-59 |  | **2007** | 289,718 | 236,348 |  | **2019** | 315,468 | 253,110 |
| NL | female | 50-59 |  | **2007** | 410,681 | 404,751 |  | **2019** | 476,284 | 469,355 |
| FR | female | 60-69 |  | **2007** | 204,444 | 165,025 |  | **2019** | 273,084 | 225,541 |
| NL | female | 60-69 |  | **2007** | 325,536 | 321,156 |  | **2019** | 404,210 | 398,927 |
| FR | female | 70-79 |  | **2007** | 191,546 | 155,322 |  | **2019** | 187,787 | 154,348 |
| NL | female | 70-79 |  | **2007** | 292,260 | 288,235 |  | **2019** | 299,830 | 296,115 |
| FR | female | 80-89 |  | **2007** | 118,656 | 93,477 |  | **2019** | 121,366 | 98,827 |
| NL | female | 80-89 |  | **2007** | 156,675 | 153,877 |  | **2019** | 205,390 | 202,886 |
| FR | female | 90+ |  | **2007** | 20,444 | 15,106 |  | **2019** | 32,932 | 25,871 |
| NL | female | 90+ |  | **2007** | 27,704 | 27,111 |  | **2019** | 48,506 | 47,721 |
| FR | male | 15-29 |  | **2007** | 419,216 | 325,446 |  | **2019** | 447,531 | 338,914 |
| NL | male | 15-29 |  | **2007** | 569,244 | 558,825 |  | **2019** | 584,879 | 572,810 |
| FR | male | 30-39 |  | **2007** | 311,907 | 232,771 |  | **2019** | 317,027 | 228,060 |
| NL | male | 30-39 |  | **2007** | 423,405 | 414,612 |  | **2019** | 424,884 | 414,999 |
| FR | male | 40-49 |  | **2007** | 320,102 | 255,519 |  | **2019** | 321,333 | 240,783 |
| NL | male | 40-49 |  | **2007** | 492,914 | 485,738 |  | **2019** | 439,423 | 430,473 |
| FR | male | 50-59 |  | **2007** | 281,323 | 231,494 |  | **2019** | 314,015 | 249,876 |
| NL | male | 50-59 |  | **2007** | 418,935 | 413,398 |  | **2019** | 487,731 | 480,604 |
| FR | male | 60-69 |  | **2007** | 182,315 | 148,453 |  | **2019** | 250,171 | 208,377 |
| NL | male | 60-69 |  | **2007** | 312,361 | 308,599 |  | **2019** | 398,263 | 393,619 |
| FR | male | 70-79 |  | **2007** | 136,518 | 111,920 |  | **2019** | 150,334 | 125,516 |
| NL | male | 70-79 |  | **2007** | 234,330 | 231,597 |  | **2019** | 264,853 | 262,095 |
| FR | male | 80-89 |  | **2007** | 57,152 | 45,231 |  | **2019** | 69,868 | 57,344 |
| NL | male | 80-89 |  | **2007** | 89,550 | 88,225 |  | **2019** | 138,028 | 136,636 |
| FR | male | 90+ |  | **2007** | 5,012 | 3,696 |  | **2019** | 10,932 | 8,579 |
| NL | male | 90+ |  | **2007** | 8,447 | 8,301 |  | **2019** | 19,968 | 19,707 |
| FR | female | 15-29 |  | **2008** | 421,879 | 319,858 |  | **2020** | 439,713 | 326,189 |
| NL | female | 15-29 |  | **2008** | 558,878 | 547,542 |  | **2020** | 567,530 | 554,916 |
| FR | female | 30-39 |  | **2008** | 306,324 | 229,712 |  | **2020** | 321,606 | 230,802 |
| NL | female | 30-39 |  | **2008** | 410,479 | 401,967 |  | **2020** | 425,263 | 415,174 |
| FR | female | 40-49 |  | **2008** | 319,061 | 255,906 |  | **2020** | 313,970 | 238,519 |
| NL | female | 40-49 |  | **2008** | 478,623 | 471,606 |  | **2020** | 431,486 | 423,103 |
| FR | female | 50-59 |  | **2008** | 291,209 | 237,543 |  | **2020** | 314,865 | 251,896 |
| NL | female | 50-59 |  | **2008** | 416,604 | 410,641 |  | **2020** | 474,420 | 467,423 |
| FR | female | 60-69 |  | **2008** | 211,647 | 171,389 |  | **2020** | 276,162 | 228,028 |
| NL | female | 60-69 |  | **2008** | 330,615 | 326,142 |  | **2020** | 411,983 | 406,635 |
| FR | female | 70-79 |  | **2008** | 188,446 | 153,053 |  | **2020** | 192,831 | 159,050 |
| NL | female | 70-79 |  | **2008** | 293,953 | 290,020 |  | **2020** | 304,301 | 300,548 |
| FR | female | 80-89 |  | **2008** | 122,835 | 97,209 |  | **2020** | 120,747 | 98,583 |
| NL | female | 80-89 |  | **2008** | 164,418 | 161,571 |  | **2020** | 207,144 | 204,681 |
| FR | female | 90+ |  | **2008** | 19,478 | 14,412 |  | **2020** | 33,820 | 26,704 |
| NL | female | 90+ |  | **2008** | 26,204 | 25,641 |  | **2020** | 50,848 | 50,057 |
| FR | male | 15-29 |  | **2008** | 424,032 | 328,542 |  | **2020** | 447,830 | 337,647 |
| NL | male | 15-29 |  | **2008** | 573,963 | 563,353 |  | **2020** | 586,223 | 573,980 |
| FR | male | 30-39 |  | **2008** | 311,808 | 230,927 |  | **2020** | 319,310 | 230,039 |
| NL | male | 30-39 |  | **2008** | 419,418 | 410,431 |  | **2020** | 426,111 | 416,192 |
| FR | male | 40-49 |  | **2008** | 321,361 | 255,209 |  | **2020** | 320,355 | 239,406 |
| NL | male | 40-49 |  | **2008** | 494,062 | 486,712 |  | **2020** | 438,728 | 429,734 |
| FR | male | 50-59 |  | **2008** | 282,458 | 232,262 |  | **2020** | 315,007 | 249,389 |
| NL | male | 50-59 |  | **2008** | 425,394 | 419,817 |  | **2020** | 486,337 | 479,046 |
| FR | male | 60-69 |  | **2008** | 189,765 | 155,376 |  | **2020** | 253,028 | 210,495 |
| NL | male | 60-69 |  | **2008** | 318,374 | 314,553 |  | **2020** | 405,966 | 401,240 |
| FR | male | 70-79 |  | **2008** | 135,834 | 111,475 |  | **2020** | 155,544 | 130,417 |
| NL | male | 70-79 |  | **2008** | 238,043 | 235,336 |  | **2020** | 271,615 | 268,823 |
| FR | male | 80-89 |  | **2008** | 59,765 | 47,566 |  | **2020** | 70,708 | 58,166 |
| NL | male | 80-89 |  | **2008** | 94,821 | 93,466 |  | **2020** | 140,792 | 139,398 |
| FR | male | 90+ |  | **2008** | 4,810 | 3,545 |  | **2020** | 11,483 | 9,039 |
| NL | male | 90+ |  | **2008** | 8,004 | 7,863 |  | **2020** | 21,247 | 20,975 |
| FR | female | 15-29 |  | **2009** | 426,819 | 322,301 |  | **2021** | 438,480 | 324,553 |
| NL | female | 15-29 |  | **2009** | 563,022 | 551,409 |  | **2021** | 565,525 | 552,866 |
| FR | female | 30-39 |  | **2009** | 307,072 | 227,957 |  | **2021** | 323,140 | 232,545 |
| NL | female | 30-39 |  | **2009** | 408,960 | 400,169 |  | **2021** | 427,375 | 417,309 |
| FR | female | 40-49 |  | **2009** | 319,067 | 254,949 |  | **2021** | 313,992 | 237,636 |
| NL | female | 40-49 |  | **2009** | 477,915 | 470,791 |  | **2021** | 431,648 | 423,164 |
| FR | female | 50-59 |  | **2009** | 293,471 | 239,024 |  | **2021** | 314,868 | 251,453 |
| NL | female | 50-59 |  | **2009** | 423,137 | 417,087 |  | **2021** | 473,161 | 466,115 |
| FR | female | 60-69 |  | **2009** | 218,138 | 177,147 |  | **2021** | 278,529 | 229,721 |
| NL | female | 60-69 |  | **2009** | 335,096 | 330,541 |  | **2021** | 419,238 | 413,815 |
| FR | female | 70-79 |  | **2009** | 186,408 | 151,652 |  | **2021** | 198,548 | 164,501 |
| NL | female | 70-79 |  | **2009** | 295,813 | 291,951 |  | **2021** | 311,085 | 307,302 |
| FR | female | 80-89 |  | **2009** | 126,616 | 100,589 |  | **2021** | 116,710 | 95,231 |
| NL | female | 80-89 |  | **2009** | 172,069 | 169,177 |  | **2021** | 203,502 | 201,115 |
| FR | female | 90+ |  | **2009** | 18,633 | 13,800 |  | **2021** | 33,390 | 26,463 |
| NL | female | 90+ |  | **2009** | 25,077 | 24,540 |  | **2021** | 52,103 | 51,333 |
| FR | male | 15-29 |  | **2009** | 428,336 | 330,887 |  | **2021** | 447,530 | 336,736 |
| NL | male | 15-29 |  | **2009** | 577,595 | 566,767 |  | **2021** | 586,040 | 573,730 |
| FR | male | 30-39 |  | **2009** | 312,528 | 229,681 |  | **2021** | 320,968 | 231,937 |
| NL | male | 30-39 |  | **2009** | 417,325 | 408,120 |  | **2021** | 427,800 | 417,908 |
| FR | male | 40-49 |  | **2009** | 322,104 | 254,068 |  | **2021** | 319,167 | 238,035 |
| NL | male | 40-49 |  | **2009** | 493,289 | 485,729 |  | **2021** | 437,572 | 428,557 |
| FR | male | 50-59 |  | **2009** | 284,380 | 233,278 |  | **2021** | 315,936 | 249,166 |
| NL | male | 50-59 |  | **2009** | 432,580 | 426,902 |  | **2021** | 485,646 | 478,227 |
| FR | male | 60-69 |  | **2009** | 196,986 | 161,904 |  | **2021** | 255,963 | 212,668 |
| NL | male | 60-69 |  | **2009** | 323,873 | 319,975 |  | **2021** | 413,509 | 408,698 |
| FR | male | 70-79 |  | **2009** | 135,626 | 111,385 |  | **2021** | 160,325 | 134,943 |
| NL | male | 70-79 |  | **2009** | 241,957 | 239,264 |  | **2021** | 278,924 | 276,104 |
| FR | male | 80-89 |  | **2009** | 61,970 | 49,456 |  | **2021** | 69,037 | 56,882 |
| NL | male | 80-89 |  | **2009** | 99,971 | 98,581 |  | **2021** | 139,310 | 137,959 |
| FR | male | 90+ |  | **2009** | 4,686 | 3,439 |  | **2021** | 11,361 | 8,983 |
| NL | male | 90+ |  | **2009** | 7,688 | 7,549 |  | **2021** | 22,168 | 21,904 |
| FR | female | 15-29 |  | **2010** | 430,201 | 324,278 |  | **2022** | 439,854 | 324,485 |
| NL | female | 15-29 |  | **2010** | 563,872 | 552,103 |  | **2022** | 568,373 | 555,554 |
| FR | female | 30-39 |  | **2010** | 308,095 | 226,639 |  | **2022** | 325,080 | 234,747 |
| NL | female | 30-39 |  | **2010** | 408,477 | 399,426 |  | **2022** | 431,150 | 421,113 |
| FR | female | 40-49 |  | **2010** | 319,308 | 253,821 |  | **2022** | 313,192 | 236,462 |
| NL | female | 40-49 |  | **2010** | 475,865 | 468,589 |  | **2022** | 432,406 | 423,880 |
| FR | female | 50-59 |  | **2010** | 297,580 | 241,879 |  | **2022** | 315,144 | 251,229 |
| NL | female | 50-59 |  | **2010** | 431,753 | 425,564 |  | **2022** | 470,650 | 463,548 |
| FR | female | 60-69 |  | **2010** | 224,191 | 182,435 |  | **2022** | 281,587 | 232,408 |
| NL | female | 60-69 |  | **2010** | 339,707 | 335,067 |  | **2022** | 427,733 | 422,269 |
| FR | female | 70-79 |  | **2010** | 184,361 | 150,055 |  | **2022** | 205,856 | 171,148 |
| NL | female | 70-79 |  | **2010** | 296,630 | 292,818 |  | **2022** | 320,453 | 316,597 |
| FR | female | 80-89 |  | **2010** | 128,464 | 102,430 |  | **2022** | 113,460 | 92,681 |
| NL | female | 80-89 |  | **2010** | 177,702 | 174,809 |  | **2022** | 199,425 | 197,116 |
| FR | female | 90+ |  | **2010** | 19,628 | 14,697 |  | **2022** | 34,460 | 27,408 |
| NL | female | 90+ |  | **2010** | 26,067 | 25,519 |  | **2022** | 55,453 | 54,669 |
| FR | male | 15-29 |  | **2010** | 431,283 | 332,641 |  | **2022** | 448,855 | 336,779 |
| NL | male | 15-29 |  | **2010** | 577,703 | 566,743 |  | **2022** | 589,283 | 576,830 |
| FR | male | 30-39 |  | **2010** | 313,248 | 228,478 |  | **2022** | 322,915 | 234,233 |
| NL | male | 30-39 |  | **2010** | 415,962 | 406,543 |  | **2022** | 431,121 | 421,267 |
| FR | male | 40-49 |  | **2010** | 323,556 | 253,574 |  | **2022** | 317,103 | 236,392 |
| NL | male | 40-49 |  | **2010** | 491,092 | 483,316 |  | **2022** | 438,375 | 429,407 |
| FR | male | 50-59 |  | **2010** | 287,481 | 235,321 |  | **2022** | 317,147 | 249,540 |
| NL | male | 50-59 |  | **2010** | 440,164 | 434,368 |  | **2022** | 483,060 | 475,548 |
| FR | male | 60-69 |  | **2010** | 203,707 | 168,002 |  | **2022** | 259,644 | 215,427 |
| NL | male | 60-69 |  | **2010** | 330,413 | 326,446 |  | **2022** | 421,835 | 416,922 |
| FR | male | 70-79 |  | **2010** | 135,566 | 111,407 |  | **2022** | 166,257 | 140,433 |
| NL | male | 70-79 |  | **2010** | 244,309 | 241,625 |  | **2022** | 287,992 | 285,123 |
| FR | male | 80-89 |  | **2010** | 63,632 | 51,017 |  | **2022** | 67,702 | 55,856 |
| NL | male | 80-89 |  | **2010** | 104,043 | 102,641 |  | **2022** | 137,345 | 136,029 |
| FR | male | 90+ |  | **2010** | 5,173 | 3,847 |  | **2022** | 11,900 | 9,469 |
| NL | male | 90+ |  | **2010** | 8,440 | 8,293 |  | **2022** | 23,977 | 23,707 |
| FR | female | 15-29 |  | **2011** | 434,396 | 326,090 |  | **2023** | 444,924 | 325,085 |
| NL | female | 15-29 |  | **2011** | 565,728 | 553,694 |  | **2023** | 577,241 | 563,926 |
| FR | female | 30-39 |  | **2011** | 310,676 | 226,458 |  | **2023** | 329,543 | 237,569 |
| NL | female | 30-39 |  | **2011** | 409,047 | 399,689 |  | **2023** | 440,060 | 429,841 |
| FR | female | 40-49 |  | **2011** | 320,734 | 253,565 |  | **2023** | 314,463 | 236,377 |
| NL | female | 40-49 |  | **2011** | 475,250 | 467,787 |  | **2023** | 437,771 | 429,095 |
| FR | female | 50-59 |  | **2011** | 301,434 | 244,232 |  | **2023** | 316,990 | 251,879 |
| NL | female | 50-59 |  | **2011** | 439,982 | 433,626 |  | **2023** | 468,445 | 461,210 |
| FR | female | 60-69 |  | **2011** | 232,055 | 189,152 |  | **2023** | 284,197 | 234,483 |
| NL | female | 60-69 |  | **2011** | 347,845 | 343,078 |  | **2023** | 436,107 | 430,583 |
| FR | female | 70-79 |  | **2011** | 180,312 | 146,640 |  | **2023** | 212,988 | 177,483 |
| NL | female | 70-79 |  | **2011** | 292,626 | 288,885 |  | **2023** | 329,775 | 325,830 |
| FR | female | 80-89 |  | **2011** | 129,291 | 103,396 |  | **2023** | 110,983 | 90,596 |
| NL | female | 80-89 |  | **2011** | 182,417 | 179,540 |  | **2023** | 197,044 | 194,779 |
| FR | female | 90+ |  | **2011** | 22,319 | 16,824 |  | **2023** | 34,657 | 27,678 |
| NL | female | 90+ |  | **2011** | 29,260 | 28,649 |  | **2023** | 57,027 | 56,252 |
| FR | male | 15-29 |  | **2011** | 435,536 | 334,535 |  | **2023** | 452,795 | 337,633 |
| NL | male | 15-29 |  | **2011** | 579,702 | 568,480 |  | **2023** | 596,278 | 583,482 |
| FR | male | 30-39 |  | **2011** | 315,695 | 228,040 |  | **2023** | 326,341 | 236,316 |
| NL | male | 30-39 |  | **2011** | 416,275 | 406,536 |  | **2023** | 437,987 | 427,984 |
| FR | male | 40-49 |  | **2011** | 326,251 | 253,615 |  | **2023** | 316,074 | 234,963 |
| NL | male | 40-49 |  | **2011** | 490,571 | 482,500 |  | **2023** | 441,787 | 432,775 |
| FR | male | 50-59 |  | **2011** | 291,578 | 237,818 |  | **2023** | 318,778 | 250,051 |
| NL | male | 50-59 |  | **2011** | 448,568 | 442,595 |  | **2023** | 479,069 | 471,433 |
| FR | male | 60-69 |  | **2011** | 211,423 | 174,831 |  | **2023** | 262,945 | 217,770 |
| NL | male | 60-69 |  | **2011** | 339,222 | 335,156 |  | **2023** | 429,841 | 424,822 |
| FR | male | 70-79 |  | **2011** | 134,080 | 110,007 |  | **2023** | 172,208 | 145,706 |
| NL | male | 70-79 |  | **2011** | 242,772 | 240,097 |  | **2023** | 297,438 | 294,493 |
| FR | male | 80-89 |  | **2011** | 64,896 | 52,306 |  | **2023** | 67,064 | 55,322 |
| NL | male | 80-89 |  | **2011** | 108,648 | 107,249 |  | **2023** | 137,541 | 136,236 |
| FR | male | 90+ |  | **2011** | 6,140 | 4,596 |  | **2023** | 12,335 | 9,780 |
| NL | male | 90+ |  | **2011** | 10,011 | 9,839 |  | **2023** | 25,057 | 24,773 |
| FR | female | 15-29 |  | **2012** | 436,979 | 327,371 |  | 2,024 | 448,555 | 325,904 |
| NL | female | 15-29 |  | **2012** | 566,942 | 554,763 |  | 2,024 | 582,847 | 569,219 |
| FR | female | 30-39 |  | **2012** | 311,289 | 225,217 |  | 2,024 | 331,614 | 238,655 |
| NL | female | 30-39 |  | **2012** | 409,066 | 399,502 |  | 2,024 | 444,010 | 433,681 |
| FR | female | 40-49 |  | **2012** | 321,823 | 253,268 |  | 2,024 | 313,871 | 235,246 |
| NL | female | 40-49 |  | **2012** | 471,983 | 464,366 |  | 2,024 | 442,102 | 433,366 |
| FR | female | 50-59 |  | **2012** | 305,265 | 246,931 |  | 2,024 | 316,247 | 250,595 |
| NL | female | 50-59 |  | **2012** | 448,824 | 442,342 |  | 2,024 | 461,915 | 454,620 |
| FR | female | 60-69 |  | **2012** | 241,431 | 197,320 |  | 2,024 | 286,949 | 236,810 |
| NL | female | 60-69 |  | **2012** | 358,667 | 353,766 |  | 2,024 | 443,949 | 438,378 |
| FR | female | 70-79 |  | **2012** | 174,536 | 141,826 |  | 2,024 | 218,682 | 182,620 |
| NL | female | 70-79 |  | **2012** | 285,864 | 282,230 |  | 2,024 | 335,866 | 331,859 |
| FR | female | 80-89 |  | **2012** | 129,436 | 103,997 |  | 2,024 | 110,832 | 90,622 |
| NL | female | 80-89 |  | **2012** | 187,188 | 184,361 |  | 2,024 | 198,600 | 196,354 |
| FR | female | 90+ |  | **2012** | 24,763 | 18,832 |  | 2,024 | 34,707 | 27,740 |
| NL | female | 90+ |  | **2012** | 32,562 | 31,903 |  | 2,024 | 58,946 | 58,172 |
| FR | male | 15-29 |  | **2012** | 438,279 | 336,242 |  | 2,024 | 457,536 | 339,840 |
| NL | male | 15-29 |  | **2012** | 579,537 | 568,200 |  | 2,024 | 605,183 | 592,106 |
| FR | male | 30-39 |  | **2012** | 315,134 | 226,066 |  | 2,024 | 328,519 | 237,749 |
| NL | male | 30-39 |  | **2012** | 416,713 | 406,817 |  | 2,024 | 442,323 | 432,237 |
| FR | male | 40-49 |  | **2012** | 328,431 | 253,813 |  | 2,024 | 314,518 | 233,863 |
| NL | male | 40-49 |  | **2012** | 487,094 | 478,803 |  | 2,024 | 444,153 | 435,191 |
| FR | male | 50-59 |  | **2012** | 295,667 | 240,592 |  | 2,024 | 319,055 | 249,305 |
| NL | male | 50-59 |  | **2012** | 456,722 | 450,603 |  | 2,024 | 472,504 | 464,754 |
| FR | male | 60-69 |  | **2012** | 219,823 | 182,165 |  | 2,024 | 266,569 | 220,404 |
| NL | male | 60-69 |  | **2012** | 350,493 | 346,309 |  | 2,024 | 438,259 | 433,130 |
| FR | male | 70-79 |  | **2012** | 130,894 | 107,363 |  | 2,024 | 177,422 | 150,552 |
| NL | male | 70-79 |  | **2012** | 238,653 | 236,038 |  | 2,024 | 304,137 | 301,151 |
| FR | male | 80-89 |  | **2012** | 66,334 | 53,702 |  | 2,024 | 67,979 | 56,016 |
| NL | male | 80-89 |  | **2012** | 113,409 | 112,005 |  | 2,024 | 140,713 | 139,384 |
| FR | male | 90+ |  | **2012** | 7,246 | 5,416 |  | 2,024 | 12,611 | 10,084 |
| NL | male | 90+ |  | **2012** | 11,577 | 11,374 |  | 2,024 | 26,362 | 26,081 |
| FR | female | 15-29 |  | **2013** | 438,947 | 328,484 |  |  |  |  |
| NL | female | 15-29 |  | **2013** | 566,981 | 554,707 |  |  |  |  |
| FR | female | 30-39 |  | **2013** | 311,298 | 223,667 |  |  |  |  |
| NL | female | 30-39 |  | **2013** | 409,415 | 399,678 |  |  |  |  |
| FR | female | 40-49 |  | **2013** | 322,958 | 253,181 |  |  |  |  |
| NL | female | 40-49 |  | **2013** | 466,790 | 459,037 |  |  |  |  |
| FR | female | 50-59 |  | **2013** | 307,468 | 248,587 |  |  |  |  |
| NL | female | 50-59 |  | **2013** | 455,637 | 449,095 |  |  |  |  |
| FR | female | 60-69 |  | **2013** | 250,050 | 204,949 |  |  |  |  |
| NL | female | 60-69 |  | **2013** | 368,349 | 363,338 |  |  |  |  |
| FR | female | 70-79 |  | **2013** | 170,053 | 138,000 |  |  |  |  |
| NL | female | 70-79 |  | **2013** | 281,105 | 277,544 |  |  |  |  |
| FR | female | 80-89 |  | **2013** | 128,999 | 104,163 |  |  |  |  |
| NL | female | 80-89 |  | **2013** | 191,218 | 188,458 |  |  |  |  |
| FR | female | 90+ |  | **2013** | 26,597 | 20,384 |  |  |  |  |
| NL | female | 90+ |  | **2013** | 34,963 | 34,273 |  |  |  |  |
| FR | male | 15-29 |  | **2013** | 440,970 | 337,726 |  |  |  |  |
| NL | male | 15-29 |  | **2013** | 579,677 | 568,205 |  |  |  |  |
| FR | male | 30-39 |  | **2013** | 314,351 | 224,258 |  |  |  |  |
| NL | male | 30-39 |  | **2013** | 416,501 | 406,491 |  |  |  |  |
| FR | male | 40-49 |  | **2013** | 330,085 | 253,886 |  |  |  |  |
| NL | male | 40-49 |  | **2013** | 480,832 | 472,365 |  |  |  |  |
| FR | male | 50-59 |  | **2013** | 298,539 | 242,341 |  |  |  |  |
| NL | male | 50-59 |  | **2013** | 463,576 | 457,332 |  |  |  |  |
| FR | male | 60-69 |  | **2013** | 227,882 | 189,264 |  |  |  |  |
| NL | male | 60-69 |  | **2013** | 360,541 | 356,250 |  |  |  |  |
| FR | male | 70-79 |  | **2013** | 128,760 | 105,553 |  |  |  |  |
| NL | male | 70-79 |  | **2013** | 236,978 | 234,399 |  |  |  |  |
| FR | male | 80-89 |  | **2013** | 67,260 | 54,616 |  |  |  |  |
| NL | male | 80-89 |  | **2013** | 117,269 | 115,864 |  |  |  |  |
| FR | male | 90+ |  | **2013** | 7,914 | 5,920 |  |  |  |  |
| NL | male | 90+ |  | **2013** | 12,838 | 12,616 |  |  |  |  |

### Supplementary file S2. Poisson regression, not adjusted for demographic characteristics

| Model 1. Main model | | | | | | | | | | | | | | | | | | | |
| --- | --- | --- | --- | --- | --- | --- | --- | --- | --- | --- | --- | --- | --- | --- | --- | --- | --- | --- | --- |
|  | Incidence rate ratio | | |  | Prevalence ratio | | |  | Prevalence ratio (sensitivity check) | | | | | | | | | | |
|  |  |  |  |  | Includng Brussels | | |  | 1. excluding Brussels | | |  | 2. baseline values, including Brussels | | |  | 3. baseline values, excluding Brussels | | |
|  | RR | 2.5 % | 97.5 % |  | PR | 2.5 % | 97.5 % |  | PR | 2.5 % | 97.5 % |  | PR | 2.5 % | 97.5 % |  | PR | 2.5 % | 97.5 % |
| (Intercept) | 1.840 | 1.744 | 1.941 |  | 0.000 | 0.000 | 0.000 |  | 0.000 | 0.000 | 0.000 |  | 0.000 | 0.000 | 0.000 |  | 0.000 | 0.000 | 0.000 |
| year | 1.059 | 1.057 | 1.061 |  | 1.059 | 1.056 | 1.061 |  | 1.060 | 1.058 | 1.063 |  | 1.069 | 1.066 | 1.071 |  | 1.070 | 1.068 | 1.073 |
| Reason: Dementia | 0.289 | 0.257 | 0.325 |  | 0.313 | 0.278 | 0.351 |  | 0.301 | 0.267 | 0.338 |  | 0.338 | 0.300 | 0.380 |  | 0.327 | 0.290 | 0.367 |
| Reason: Multimorbidity | 0.375 | 0.363 | 0.387 |  | 0.491 | 0.475 | 0.507 |  | 0.479 | 0.464 | 0.495 |  | 0.522 | 0.505 | 0.539 |  | 0.514 | 0.498 | 0.531 |
| Reason: Nervous system diseases | 0.213 | 0.204 | 0.222 |  | 0.195 | 0.187 | 0.203 |  | 0.193 | 0.185 | 0.201 |  | 0.199 | 0.191 | 0.208 |  | 0.197 | 0.189 | 0.206 |
| Reason: Others | 0.135 | 0.122 | 0.149 |  | 0.134 | 0.121 | 0.149 |  | 0.129 | 0.117 | 0.143 |  | 0.141 | 0.127 | 0.156 |  | 0.136 | 0.122 | 0.150 |
| Reason: Psychiatric disorders | 0.369 | 0.332 | 0.409 |  | 0.272 | 0.244 | 0.301 |  | 0.259 | 0.233 | 0.288 |  | 0.263 | 0.237 | 0.293 |  | 0.251 | 0.225 | 0.279 |
| Reason: Specific diseases | 0.241 | 0.232 | 0.251 |  | 0.291 | 0.279 | 0.303 |  | 0.283 | 0.272 | 0.295 |  | 0.307 | 0.295 | 0.320 |  | 0.302 | 0.290 | 0.314 |
| Basis: advanced | 0.226 | 0.205 | 0.248 |  | 0.216 | 0.196 | 0.237 |  | 0.215 | 0.195 | 0.235 |  | 0.222 | 0.202 | 0.243 |  | 0.221 | 0.201 | 0.243 |
| Suffering: both | 1.792 | 1.746 | 1.839 |  | 1.793 | 1.747 | 1.840 |  | 1.823 | 1.776 | 1.871 |  | 1.757 | 1.713 | 1.804 |  | 1.784 | 1.738 | 1.830 |
| Suffering: mental | 0.666 | 0.620 | 0.715 |  | 0.702 | 0.653 | 0.753 |  | 0.705 | 0.655 | 0.757 |  | 0.708 | 0.659 | 0.761 |  | 0.713 | 0.663 | 0.766 |
| Term: Short term | 1.797 | 1.737 | 1.859 |  | 1.896 | 1.832 | 1.962 |  | 1.901 | 1.837 | 1.968 |  | 1.899 | 1.835 | 1.966 |  | 1.903 | 1.838 | 1.970 |
| Place: Hospital | 0.819 | 0.800 | 0.838 |  | 0.846 | 0.826 | 0.867 |  | 0.855 | 0.835 | 0.875 |  | 0.850 | 0.831 | 0.871 |  | 0.859 | 0.839 | 0.880 |
| Place: Nursing home | 0.551 | 0.533 | 0.570 |  | 0.797 | 0.771 | 0.825 |  | 0.791 | 0.765 | 0.818 |  | 0.853 | 0.825 | 0.883 |  | 0.855 | 0.826 | 0.884 |
| Place: Other | 0.169 | 0.155 | 0.183 |  | 0.163 | 0.150 | 0.177 |  | 0.159 | 0.147 | 0.173 |  | 0.169 | 0.156 | 0.183 |  | 0.165 | 0.152 | 0.179 |
| Place: Palliative care | 0.244 | 0.227 | 0.262 |  | 0.254 | 0.236 | 0.272 |  | 0.248 | 0.231 | 0.267 |  | 0.263 | 0.244 | 0.282 |  | 0.258 | 0.240 | 0.278 |

| Model 2. Interaction between year and reason for euthanasia | | | | | | | | | | | | | | | | | | | |
| --- | --- | --- | --- | --- | --- | --- | --- | --- | --- | --- | --- | --- | --- | --- | --- | --- | --- | --- | --- |
|  | Incidence rate ratio | | |  | Prevalence ratio | | |  | Prevalence ratio (sensitivity check) | | | | | | | | | | |
|  |  |  |  |  | Includng Brussels | | |  | 1. excluding Brussels | | |  | 2. baseline values, including Brussels | | |  | 3. baseline values, excluding Brussels | | |
|  | RR | 2.5 % | 97.5 % |  | PR | 2.5 % | 97.5 % |  | PR | 2.5 % | 97.5 % |  | PR | 2.5 % | 97.5 % |  | PR | 2.5 % | 97.5 % |
| (Intercept) | 1.809 | 1.710 | 1.913 |  | 0.000 | 0.000 | 0.000 |  | 0.000 | 0.000 | 0.000 |  | 0.000 | 0.000 | 0.000 |  | 0.000 | 0.000 | 0.000 |
| year | 1.060 | 1.058 | 1.063 |  | 1.060 | 1.058 | 1.063 |  | 1.062 | 1.059 | 1.065 |  | 1.070 | 1.067 | 1.072 |  | 1.071 | 1.069 | 1.074 |
| Reason: Dementia | 0.437 | 0.292 | 0.638 |  | 0.579 | 0.391 | 0.839 |  | 0.556 | 0.376 | 0.805 |  | 0.595 | 0.403 | 0.860 |  | 0.568 | 0.385 | 0.821 |
| Reason: Multimorbidity | 0.263 | 0.236 | 0.294 |  | 0.365 | 0.327 | 0.407 |  | 0.355 | 0.318 | 0.396 |  | 0.371 | 0.332 | 0.413 |  | 0.360 | 0.322 | 0.401 |
| Reason: Nervous system diseases | 0.260 | 0.228 | 0.296 |  | 0.238 | 0.209 | 0.271 |  | 0.233 | 0.204 | 0.265 |  | 0.238 | 0.209 | 0.271 |  | 0.233 | 0.204 | 0.265 |
| Reason: Others | 0.338 | 0.238 | 0.472 |  | 0.311 | 0.221 | 0.430 |  | 0.298 | 0.212 | 0.412 |  | 0.322 | 0.229 | 0.444 |  | 0.308 | 0.219 | 0.425 |
| Reason: Psychiatric disorders | 0.757 | 0.548 | 1.035 |  | 0.544 | 0.393 | 0.744 |  | 0.511 | 0.369 | 0.700 |  | 0.550 | 0.397 | 0.753 |  | 0.516 | 0.372 | 0.706 |
| Reason: Specific diseases | 0.346 | 0.304 | 0.394 |  | 0.431 | 0.378 | 0.490 |  | 0.415 | 0.364 | 0.472 |  | 0.431 | 0.378 | 0.490 |  | 0.415 | 0.364 | 0.471 |
| Basis: advanced | 0.226 | 0.205 | 0.248 |  | 0.216 | 0.196 | 0.237 |  | 0.215 | 0.195 | 0.236 |  | 0.222 | 0.202 | 0.243 |  | 0.221 | 0.201 | 0.243 |
| Suffering: both | 1.792 | 1.747 | 1.839 |  | 1.792 | 1.746 | 1.839 |  | 1.822 | 1.776 | 1.870 |  | 1.757 | 1.712 | 1.803 |  | 1.784 | 1.738 | 1.831 |
| Suffering: mental | 0.667 | 0.621 | 0.716 |  | 0.703 | 0.654 | 0.755 |  | 0.706 | 0.657 | 0.758 |  | 0.710 | 0.661 | 0.763 |  | 0.715 | 0.665 | 0.768 |
| Term: Short term | 1.801 | 1.741 | 1.863 |  | 1.897 | 1.833 | 1.964 |  | 1.903 | 1.839 | 1.970 |  | 1.902 | 1.838 | 1.969 |  | 1.906 | 1.842 | 1.974 |
| Place: Hospital | 0.819 | 0.799 | 0.838 |  | 0.847 | 0.827 | 0.867 |  | 0.856 | 0.836 | 0.876 |  | 0.851 | 0.831 | 0.872 |  | 0.861 | 0.840 | 0.881 |
| Place: Nursing home | 0.551 | 0.532 | 0.569 |  | 0.797 | 0.770 | 0.824 |  | 0.790 | 0.764 | 0.818 |  | 0.853 | 0.824 | 0.882 |  | 0.854 | 0.826 | 0.884 |
| Place: Other | 0.168 | 0.155 | 0.182 |  | 0.163 | 0.150 | 0.177 |  | 0.159 | 0.147 | 0.172 |  | 0.169 | 0.155 | 0.183 |  | 0.165 | 0.152 | 0.179 |
| Place: Palliative care | 0.243 | 0.225 | 0.261 |  | 0.252 | 0.234 | 0.271 |  | 0.247 | 0.229 | 0.265 |  | 0.261 | 0.243 | 0.281 |  | 0.257 | 0.239 | 0.276 |
| Reason: Dementia * Year | 0.974 | 0.952 | 0.998 |  | 0.962 | 0.940 | 0.985 |  | 0.962 | 0.940 | 0.985 |  | 0.965 | 0.943 | 0.988 |  | 0.966 | 0.944 | 0.989 |
| Reason: Multimorbidity * Year | 1.022 | 1.016 | 1.029 |  | 1.019 | 1.012 | 1.025 |  | 1.019 | 1.012 | 1.026 |  | 1.022 | 1.015 | 1.029 |  | 1.023 | 1.016 | 1.030 |
| Reason: Nervous system diseases * Year | 0.987 | 0.979 | 0.995 |  | 0.987 | 0.979 | 0.995 |  | 0.987 | 0.979 | 0.996 |  | 0.988 | 0.980 | 0.996 |  | 0.989 | 0.981 | 0.997 |
| Reason: Others * Year | 0.943 | 0.922 | 0.964 |  | 0.947 | 0.927 | 0.968 |  | 0.947 | 0.928 | 0.968 |  | 0.948 | 0.928 | 0.968 |  | 0.948 | 0.929 | 0.969 |
| Reason: Psychiatric disorders * Year | 0.953 | 0.934 | 0.973 |  | 0.955 | 0.935 | 0.975 |  | 0.956 | 0.936 | 0.976 |  | 0.952 | 0.932 | 0.972 |  | 0.953 | 0.933 | 0.973 |
| Reason: Specific diseases * Year | 0.976 | 0.968 | 0.985 |  | 0.974 | 0.966 | 0.982 |  | 0.975 | 0.967 | 0.983 |  | 0.978 | 0.970 | 0.986 |  | 0.979 | 0.971 | 0.987 |

| Model 3. Interaction between year and basis for euthanasia | | | | | | | | | | | | | | | | | | | |
| --- | --- | --- | --- | --- | --- | --- | --- | --- | --- | --- | --- | --- | --- | --- | --- | --- | --- | --- | --- |
|  | Incidence rate ratio | | |  | Prevalence ratio | | |  | Prevalence ratio (sensitivity check) | | | | | | | | | | |
|  |  |  |  |  | Includng Brussels | | |  | 1. excluding Brussels | | |  | 2. baseline values, including Brussels | | |  | 3. baseline values, excluding Brussels | | |
|  | RR | 2.5 % | 97.5 % |  | PR | 2.5 % | 97.5 % |  | PR | 2.5 % | 97.5 % |  | PR | 2.5 % | 97.5 % |  | PR | 2.5 % | 97.5 % |
| (Intercept) | 1.831 | 1.735 | 1.931 |  | 0.000 | 0.000 | 0.000 |  | 0.000 | 0.000 | 0.000 |  | 0.000 | 0.000 | 0.000 |  | 0.000 | 0.000 | 0.000 |
| year | 1.060 | 1.057 | 1.062 |  | 1.059 | 1.057 | 1.061 |  | 1.061 | 1.058 | 1.063 |  | 1.069 | 1.067 | 1.071 |  | 1.071 | 1.068 | 1.073 |
| Basis: advanced | 0.364 | 0.283 | 0.463 |  | 0.269 | 0.209 | 0.343 |  | 0.258 | 0.200 | 0.329 |  | 0.266 | 0.206 | 0.340 |  | 0.255 | 0.198 | 0.326 |
| Reason: Dementia | 0.289 | 0.257 | 0.325 |  | 0.313 | 0.278 | 0.351 |  | 0.301 | 0.267 | 0.338 |  | 0.338 | 0.300 | 0.380 |  | 0.327 | 0.290 | 0.367 |
| Reason: Multimorbidity | 0.375 | 0.363 | 0.387 |  | 0.491 | 0.475 | 0.507 |  | 0.479 | 0.464 | 0.495 |  | 0.522 | 0.505 | 0.539 |  | 0.514 | 0.498 | 0.531 |
| Reason: Nervous system diseases | 0.213 | 0.204 | 0.222 |  | 0.195 | 0.187 | 0.203 |  | 0.193 | 0.185 | 0.201 |  | 0.199 | 0.191 | 0.208 |  | 0.197 | 0.189 | 0.206 |
| Reason: Others | 0.135 | 0.122 | 0.149 |  | 0.134 | 0.121 | 0.149 |  | 0.129 | 0.117 | 0.143 |  | 0.140 | 0.126 | 0.156 |  | 0.136 | 0.122 | 0.150 |
| Reason: Psychiatric disorders | 0.369 | 0.332 | 0.409 |  | 0.272 | 0.244 | 0.302 |  | 0.259 | 0.233 | 0.288 |  | 0.263 | 0.237 | 0.293 |  | 0.251 | 0.226 | 0.279 |
| Reason: Specific diseases | 0.241 | 0.232 | 0.251 |  | 0.291 | 0.279 | 0.303 |  | 0.283 | 0.272 | 0.295 |  | 0.308 | 0.295 | 0.320 |  | 0.302 | 0.290 | 0.314 |
| Suffering: both | 1.787 | 1.742 | 1.834 |  | 1.791 | 1.745 | 1.838 |  | 1.821 | 1.775 | 1.869 |  | 1.756 | 1.711 | 1.802 |  | 1.782 | 1.737 | 1.829 |
| Suffering: mental | 0.664 | 0.618 | 0.713 |  | 0.701 | 0.652 | 0.753 |  | 0.704 | 0.655 | 0.756 |  | 0.708 | 0.658 | 0.760 |  | 0.713 | 0.663 | 0.765 |
| Term: Short term | 1.797 | 1.737 | 1.860 |  | 1.896 | 1.832 | 1.963 |  | 1.901 | 1.837 | 1.968 |  | 1.899 | 1.835 | 1.966 |  | 1.903 | 1.838 | 1.970 |
| Place: Hospital | 0.819 | 0.799 | 0.838 |  | 0.846 | 0.826 | 0.867 |  | 0.855 | 0.835 | 0.875 |  | 0.850 | 0.830 | 0.871 |  | 0.859 | 0.839 | 0.880 |
| Place: Nursing home | 0.551 | 0.533 | 0.570 |  | 0.797 | 0.771 | 0.825 |  | 0.791 | 0.764 | 0.818 |  | 0.853 | 0.825 | 0.883 |  | 0.855 | 0.826 | 0.884 |
| Place: Other | 0.169 | 0.155 | 0.183 |  | 0.163 | 0.150 | 0.177 |  | 0.159 | 0.147 | 0.173 |  | 0.169 | 0.156 | 0.183 |  | 0.165 | 0.152 | 0.179 |
| Place: Palliative care | 0.244 | 0.227 | 0.262 |  | 0.253 | 0.235 | 0.272 |  | 0.248 | 0.231 | 0.267 |  | 0.263 | 0.244 | 0.282 |  | 0.258 | 0.240 | 0.278 |
| Basis: advanced * Year | 0.963 | 0.946 | 0.981 |  | 0.983 | 0.964 | 1.001 |  | 0.985 | 0.967 | 1.004 |  | 0.985 | 0.967 | 1.004 |  | 0.989 | 0.970 | 1.007 |

| Model 4. Interaction between year and type of suffering | | | | | | | | | | | | | | | | | | | |
| --- | --- | --- | --- | --- | --- | --- | --- | --- | --- | --- | --- | --- | --- | --- | --- | --- | --- | --- | --- |
|  | Incidence rate ratio | | |  | Prevalence ratio | | |  | Prevalence ratio (sensitivity check) | | | | | | | | | | |
|  |  |  |  |  | Includng Brussels | | |  | 1. excluding Brussels | | |  | 2. baseline values, including Brussels | | |  | 3. baseline values, excluding Brussels | | |
|  | RR | 2.5 % | 97.5 % |  | PR | 2.5 % | 97.5 % |  | PR | 2.5 % | 97.5 % |  | PR | 2.5 % | 97.5 % |  | PR | 2.5 % | 97.5 % |
| (Intercept) | 2.164 | 2.013 | 2.326 |  | 0.000 | 0.000 | 0.000 |  | 0.000 | 0.000 | 0.000 |  | 0.000 | 0.000 | 0.000 |  | 0.000 | 0.000 | 0.000 |
| year | 1.047 | 1.043 | 1.051 |  | 1.051 | 1.046 | 1.055 |  | 1.052 | 1.048 | 1.057 |  | 1.061 | 1.057 | 1.065 |  | 1.063 | 1.058 | 1.067 |
| Suffering: both | 1.419 | 1.318 | 1.528 |  | 1.532 | 1.424 | 1.648 |  | 1.553 | 1.444 | 1.671 |  | 1.510 | 1.404 | 1.625 |  | 1.530 | 1.422 | 1.646 |
| Suffering: mental | 0.682 | 0.574 | 0.806 |  | 0.762 | 0.643 | 0.898 |  | 0.758 | 0.640 | 0.894 |  | 0.762 | 0.644 | 0.898 |  | 0.759 | 0.641 | 0.894 |
| Reason: Dementia | 0.293 | 0.260 | 0.329 |  | 0.318 | 0.282 | 0.358 |  | 0.306 | 0.271 | 0.343 |  | 0.344 | 0.305 | 0.386 |  | 0.332 | 0.294 | 0.373 |
| Reason: Multimorbidity | 0.375 | 0.363 | 0.387 |  | 0.491 | 0.475 | 0.507 |  | 0.479 | 0.464 | 0.495 |  | 0.522 | 0.505 | 0.539 |  | 0.514 | 0.498 | 0.531 |
| Reason: Nervous system diseases | 0.213 | 0.204 | 0.222 |  | 0.195 | 0.187 | 0.203 |  | 0.193 | 0.185 | 0.201 |  | 0.199 | 0.191 | 0.208 |  | 0.197 | 0.189 | 0.206 |
| Reason: Others | 0.134 | 0.121 | 0.149 |  | 0.134 | 0.121 | 0.148 |  | 0.129 | 0.116 | 0.143 |  | 0.140 | 0.126 | 0.155 |  | 0.135 | 0.122 | 0.150 |
| Reason: Psychiatric disorders | 0.374 | 0.336 | 0.415 |  | 0.276 | 0.248 | 0.307 |  | 0.263 | 0.237 | 0.293 |  | 0.268 | 0.240 | 0.298 |  | 0.255 | 0.229 | 0.284 |
| Reason: Specific diseases | 0.242 | 0.232 | 0.251 |  | 0.291 | 0.280 | 0.303 |  | 0.284 | 0.272 | 0.295 |  | 0.308 | 0.296 | 0.320 |  | 0.302 | 0.290 | 0.315 |
| Basis: advanced | 0.227 | 0.206 | 0.249 |  | 0.216 | 0.196 | 0.237 |  | 0.215 | 0.195 | 0.236 |  | 0.222 | 0.202 | 0.244 |  | 0.221 | 0.201 | 0.243 |
| Term: Short term | 1.802 | 1.742 | 1.865 |  | 1.900 | 1.836 | 1.967 |  | 1.905 | 1.841 | 1.972 |  | 1.903 | 1.839 | 1.970 |  | 1.907 | 1.842 | 1.974 |
| Place: Hospital | 0.819 | 0.799 | 0.838 |  | 0.846 | 0.826 | 0.866 |  | 0.855 | 0.834 | 0.875 |  | 0.850 | 0.830 | 0.871 |  | 0.859 | 0.839 | 0.880 |
| Place: Nursing home | 0.551 | 0.533 | 0.570 |  | 0.797 | 0.771 | 0.825 |  | 0.791 | 0.764 | 0.818 |  | 0.853 | 0.825 | 0.883 |  | 0.855 | 0.826 | 0.884 |
| Place: Other | 0.168 | 0.155 | 0.182 |  | 0.163 | 0.150 | 0.177 |  | 0.159 | 0.147 | 0.172 |  | 0.169 | 0.155 | 0.183 |  | 0.165 | 0.152 | 0.179 |
| Place: Palliative care | 0.243 | 0.225 | 0.261 |  | 0.252 | 0.235 | 0.271 |  | 0.247 | 0.230 | 0.266 |  | 0.262 | 0.243 | 0.281 |  | 0.257 | 0.239 | 0.277 |
| Suffering: both * Year | 1.016 | 1.011 | 1.021 |  | 1.011 | 1.006 | 1.016 |  | 1.011 | 1.006 | 1.016 |  | 1.011 | 1.006 | 1.015 |  | 1.011 | 1.006 | 1.015 |
| Suffering: mental * Year | 0.997 | 0.985 | 1.009 |  | 0.993 | 0.981 | 1.005 |  | 0.994 | 0.982 | 1.005 |  | 0.994 | 0.982 | 1.005 |  | 0.994 | 0.983 | 1.006 |

| Model 5. Interaction between year and expected term of death | | | | | | | | | | | | | | | | | | | |
| --- | --- | --- | --- | --- | --- | --- | --- | --- | --- | --- | --- | --- | --- | --- | --- | --- | --- | --- | --- |
|  | Incidence rate ratio | | |  | Prevalence ratio | | |  | Prevalence ratio (sensitivity check) | | | | | | | | | | |
|  |  |  |  |  | Includng Brussels | | |  | 1. excluding Brussels | | |  | 2. baseline values, including Brussels | | |  | 3. baseline values, excluding Brussels | | |
|  | RR | 2.5 % | 97.5 % |  | PR | 2.5 % | 97.5 % |  | PR | 2.5 % | 97.5 % |  | PR | 2.5 % | 97.5 % |  | PR | 2.5 % | 97.5 % |
| (Intercept) | 2.173 | 1.946 | 2.422 |  | 0.000 | 0.000 | 0.000 |  | 0.000 | 0.000 | 0.000 |  | 0.000 | 0.000 | 0.000 |  | 0.000 | 0.000 | 0.000 |
| year | 1.048 | 1.041 | 1.055 |  | 1.044 | 1.037 | 1.050 |  | 1.046 | 1.039 | 1.052 |  | 1.054 | 1.048 | 1.061 |  | 1.057 | 1.050 | 1.063 |
| Term: Short term | 1.496 | 1.340 | 1.673 |  | 1.474 | 1.321 | 1.647 |  | 1.490 | 1.335 | 1.665 |  | 1.495 | 1.340 | 1.670 |  | 1.517 | 1.359 | 1.695 |
| Reason: Dementia | 0.291 | 0.258 | 0.326 |  | 0.316 | 0.281 | 0.355 |  | 0.304 | 0.270 | 0.341 |  | 0.341 | 0.303 | 0.383 |  | 0.330 | 0.293 | 0.371 |
| Reason: Multimorbidity | 0.375 | 0.363 | 0.387 |  | 0.491 | 0.475 | 0.507 |  | 0.479 | 0.464 | 0.495 |  | 0.522 | 0.506 | 0.539 |  | 0.514 | 0.498 | 0.531 |
| Reason: Nervous system diseases | 0.212 | 0.204 | 0.221 |  | 0.195 | 0.187 | 0.203 |  | 0.192 | 0.184 | 0.200 |  | 0.199 | 0.191 | 0.207 |  | 0.197 | 0.189 | 0.205 |
| Reason: Others | 0.135 | 0.122 | 0.149 |  | 0.134 | 0.121 | 0.149 |  | 0.129 | 0.116 | 0.143 |  | 0.141 | 0.127 | 0.156 |  | 0.136 | 0.122 | 0.150 |
| Reason: Psychiatric disorders | 0.368 | 0.331 | 0.408 |  | 0.270 | 0.243 | 0.300 |  | 0.258 | 0.232 | 0.286 |  | 0.262 | 0.236 | 0.291 |  | 0.250 | 0.225 | 0.278 |
| Reason: Specific diseases | 0.241 | 0.231 | 0.251 |  | 0.290 | 0.279 | 0.302 |  | 0.283 | 0.271 | 0.294 |  | 0.307 | 0.295 | 0.319 |  | 0.301 | 0.289 | 0.314 |
| Basis: advanced | 0.226 | 0.206 | 0.248 |  | 0.217 | 0.197 | 0.238 |  | 0.215 | 0.195 | 0.236 |  | 0.222 | 0.202 | 0.244 |  | 0.222 | 0.201 | 0.243 |
| Suffering: both | 1.792 | 1.746 | 1.839 |  | 1.792 | 1.747 | 1.840 |  | 1.823 | 1.776 | 1.870 |  | 1.757 | 1.712 | 1.803 |  | 1.783 | 1.738 | 1.830 |
| Suffering: mental | 0.666 | 0.620 | 0.715 |  | 0.701 | 0.652 | 0.753 |  | 0.704 | 0.655 | 0.756 |  | 0.708 | 0.658 | 0.760 |  | 0.712 | 0.662 | 0.765 |
| Place: Hospital | 0.818 | 0.799 | 0.838 |  | 0.845 | 0.826 | 0.866 |  | 0.854 | 0.834 | 0.875 |  | 0.850 | 0.830 | 0.870 |  | 0.859 | 0.839 | 0.880 |
| Place: Nursing home | 0.551 | 0.532 | 0.570 |  | 0.797 | 0.770 | 0.825 |  | 0.791 | 0.764 | 0.818 |  | 0.853 | 0.825 | 0.883 |  | 0.855 | 0.826 | 0.884 |
| Place: Other | 0.169 | 0.155 | 0.183 |  | 0.163 | 0.150 | 0.177 |  | 0.159 | 0.147 | 0.173 |  | 0.169 | 0.156 | 0.183 |  | 0.165 | 0.152 | 0.179 |
| Place: Palliative care | 0.244 | 0.226 | 0.262 |  | 0.253 | 0.235 | 0.272 |  | 0.248 | 0.230 | 0.266 |  | 0.262 | 0.243 | 0.282 |  | 0.258 | 0.239 | 0.277 |
| Term: Short term * Year | 1.012 | 1.005 | 1.019 |  | 1.016 | 1.009 | 1.023 |  | 1.016 | 1.009 | 1.023 |  | 1.015 | 1.009 | 1.022 |  | 1.015 | 1.008 | 1.021 |

| Model 6. Interaction between year and place of death | | | | | | | | | | | | | | |  |  |  |  |  |
| --- | --- | --- | --- | --- | --- | --- | --- | --- | --- | --- | --- | --- | --- | --- | --- | --- | --- | --- | --- |
|  | Incidence rate ratio | | |  | Prevalence ratio | | |  | Prevalence ratio (sensitivity check) | | | | | | | | | | |
|  |  |  |  |  | Includng Brussels | | |  | 1. excluding Brussels | | |  | 2. baseline values, including Brussels | | |  | 3. baseline values, excluding Brussels | | |
|  | RR | 2.5 % | 97.5 % |  | PR | 2.5 % | 97.5 % |  | PR | 2.5 % | 97.5 % |  | PR | 2.5 % | 97.5 % |  | PR | 2.5 % | 97.5 % |
| (Intercept) | 1.496 | 1.403 | 1.594 |  | 0.000 | 0.000 | 0.000 |  | 0.000 | 0.000 | 0.000 |  | 0.000 | 0.000 | 0.000 |  | 0.000 | 0.000 | 0.000 |
| year | 1.074 | 1.070 | 1.077 |  | 1.074 | 1.071 | 1.077 |  | 1.076 | 1.072 | 1.079 |  | 1.084 | 1.080 | 1.087 |  | 1.085 | 1.082 | 1.088 |
| Place: Hospital | 1.278 | 1.194 | 1.368 |  | 1.298 | 1.213 | 1.389 |  | 1.305 | 1.219 | 1.396 |  | 1.292 | 1.208 | 1.383 |  | 1.298 | 1.213 | 1.389 |
| Place: Nursing home | 0.594 | 0.525 | 0.671 |  | 1.082 | 0.957 | 1.221 |  | 1.080 | 0.956 | 1.219 |  | 1.102 | 0.975 | 1.243 |  | 1.098 | 0.972 | 1.240 |
| Place: Other | 0.327 | 0.252 | 0.420 |  | 0.285 | 0.219 | 0.367 |  | 0.274 | 0.211 | 0.353 |  | 0.288 | 0.221 | 0.371 |  | 0.277 | 0.213 | 0.357 |
| Place: Palliative care | 0.235 | 0.168 | 0.322 |  | 0.226 | 0.162 | 0.307 |  | 0.210 | 0.151 | 0.287 |  | 0.235 | 0.169 | 0.319 |  | 0.219 | 0.157 | 0.298 |
| Reason: Dementia | 0.291 | 0.258 | 0.326 |  | 0.315 | 0.279 | 0.353 |  | 0.302 | 0.268 | 0.339 |  | 0.340 | 0.301 | 0.381 |  | 0.328 | 0.291 | 0.369 |
| Reason: Multimorbidity | 0.375 | 0.364 | 0.387 |  | 0.490 | 0.475 | 0.506 |  | 0.479 | 0.463 | 0.494 |  | 0.522 | 0.505 | 0.539 |  | 0.513 | 0.497 | 0.530 |
| Reason: Nervous system diseases | 0.213 | 0.204 | 0.222 |  | 0.195 | 0.187 | 0.204 |  | 0.193 | 0.185 | 0.201 |  | 0.200 | 0.191 | 0.208 |  | 0.197 | 0.189 | 0.206 |
| Reason: Others | 0.136 | 0.122 | 0.150 |  | 0.135 | 0.122 | 0.150 |  | 0.130 | 0.117 | 0.144 |  | 0.142 | 0.128 | 0.157 |  | 0.137 | 0.123 | 0.152 |
| Reason: Psychiatric disorders | 0.369 | 0.332 | 0.409 |  | 0.271 | 0.244 | 0.301 |  | 0.259 | 0.233 | 0.288 |  | 0.263 | 0.236 | 0.292 |  | 0.251 | 0.225 | 0.279 |
| Reason: Specific diseases | 0.241 | 0.232 | 0.251 |  | 0.291 | 0.279 | 0.303 |  | 0.283 | 0.272 | 0.295 |  | 0.308 | 0.295 | 0.320 |  | 0.302 | 0.290 | 0.314 |
| Basis: advanced | 0.226 | 0.205 | 0.248 |  | 0.215 | 0.196 | 0.236 |  | 0.214 | 0.194 | 0.235 |  | 0.221 | 0.201 | 0.243 |  | 0.221 | 0.200 | 0.242 |
| Suffering: both | 1.792 | 1.746 | 1.839 |  | 1.793 | 1.747 | 1.840 |  | 1.823 | 1.777 | 1.871 |  | 1.757 | 1.712 | 1.803 |  | 1.784 | 1.738 | 1.831 |
| Suffering: mental | 0.660 | 0.614 | 0.708 |  | 0.697 | 0.648 | 0.749 |  | 0.700 | 0.651 | 0.752 |  | 0.705 | 0.655 | 0.757 |  | 0.709 | 0.659 | 0.762 |
| Term: Short term | 1.806 | 1.746 | 1.868 |  | 1.904 | 1.839 | 1.971 |  | 1.909 | 1.844 | 1.976 |  | 1.906 | 1.841 | 1.973 |  | 1.909 | 1.844 | 1.977 |
| Place: Hospital * Year | 0.969 | 0.965 | 0.973 |  | 0.970 | 0.966 | 0.975 |  | 0.971 | 0.966 | 0.975 |  | 0.971 | 0.967 | 0.975 |  | 0.971 | 0.967 | 0.976 |
| Place: Nursing home * Year | 0.994 | 0.987 | 1.002 |  | 0.980 | 0.973 | 0.987 |  | 0.980 | 0.972 | 0.987 |  | 0.983 | 0.976 | 0.990 |  | 0.983 | 0.976 | 0.991 |
| Place: Other * Year | 0.956 | 0.941 | 0.972 |  | 0.963 | 0.947 | 0.980 |  | 0.964 | 0.948 | 0.980 |  | 0.965 | 0.949 | 0.981 |  | 0.966 | 0.950 | 0.982 |
| Place: Palliative care * Year | 0.999 | 0.983 | 1.017 |  | 1.004 | 0.987 | 1.021 |  | 1.006 | 0.990 | 1.024 |  | 1.004 | 0.987 | 1.021 |  | 1.007 | 0.990 | 1.024 |

### Supplementary file S3. Poisson regression, adjusted for demographic characteristics

| Model 1. Main model | | | | | | | | | | | | | | | | | | | |
| --- | --- | --- | --- | --- | --- | --- | --- | --- | --- | --- | --- | --- | --- | --- | --- | --- | --- | --- | --- |
|  | Incidence rate ratio | | |  | Prevalence ratio | | |  | Prevalence ratio (sensitivity check) | | | | | | | | | | |
|  |  |  |  |  | Includng Brussels | | |  | 1. excluding Brussels | | |  | 2. baseline values, including Brussels | | |  | 3. baseline values, excluding Brussels | | |
|  | RR | 2.5 % | 97.5 % |  | PR | 2.5 % | 97.5 % |  | PR | 2.5 % | 97.5 % |  | PR | 2.5 % | 97.5 % |  | PR | 2.5 % | 97.5 % |
| (Intercept) | 0.619 | 0.579 | 0.661 |  | 0.000 | 0.000 | 0.000 |  | 0.000 | 0.000 | 0.000 |  | 0.000 | 0.000 | 0.000 |  | 0.000 | 0.000 | 0.000 |
| year | 1.070 | 1.067 | 1.072 |  | 1.054 | 1.051 | 1.056 |  | 1.053 | 1.051 | 1.055 |  | 1.070 | 1.068 | 1.073 |  | 1.070 | 1.068 | 1.073 |
| Age group: 15-29 | 0.212 | 0.176 | 0.253 |  | 0.165 | 0.137 | 0.197 |  | 0.168 | 0.139 | 0.200 |  | 0.142 | 0.118 | 0.170 |  | 0.144 | 0.120 | 0.172 |
| Age group: 30-39 | 0.292 | 0.263 | 0.323 |  | 0.312 | 0.281 | 0.346 |  | 0.320 | 0.288 | 0.355 |  | 0.247 | 0.223 | 0.274 |  | 0.251 | 0.226 | 0.278 |
| Age group: 40-49 | 0.486 | 0.455 | 0.520 |  | 0.489 | 0.457 | 0.523 |  | 0.495 | 0.463 | 0.529 |  | 0.411 | 0.385 | 0.440 |  | 0.412 | 0.386 | 0.441 |
| Age group: 60-69 | 1.632 | 1.566 | 1.700 |  | 1.998 | 1.918 | 2.082 |  | 1.982 | 1.902 | 2.065 |  | 2.147 | 2.061 | 2.237 |  | 2.148 | 2.062 | 2.238 |
| Age group: 70-79 | 1.878 | 1.805 | 1.954 |  | 3.266 | 3.139 | 3.398 |  | 3.236 | 3.111 | 3.368 |  | 2.964 | 2.849 | 3.085 |  | 2.965 | 2.850 | 3.086 |
| Age group: 80-89 | 1.798 | 1.728 | 1.871 |  | 5.653 | 5.432 | 5.885 |  | 5.626 | 5.406 | 5.857 |  | 7.294 | 7.008 | 7.593 |  | 7.399 | 7.109 | 7.703 |
| Age group: 90+ | 0.837 | 0.797 | 0.879 |  | 13.186 | 12.547 | 13.856 |  | 13.254 | 12.613 | 13.928 |  | 19.659 | 18.706 | 20.660 |  | 20.220 | 19.240 | 21.250 |
| Gender: male | 1.046 | 1.024 | 1.069 |  | 1.363 | 1.334 | 1.393 |  | 1.359 | 1.330 | 1.389 |  | 1.538 | 1.505 | 1.572 |  | 1.533 | 1.499 | 1.567 |
| Language: NL | 2.451 | 2.389 | 2.515 |  | 1.512 | 1.474 | 1.551 |  | 1.246 | 1.214 | 1.278 |  | 1.656 | 1.614 | 1.699 |  | 1.332 | 1.298 | 1.366 |
| Reason: Dementia | 0.199 | 0.177 | 0.223 |  | 0.198 | 0.176 | 0.222 |  | 0.198 | 0.176 | 0.223 |  | 0.198 | 0.176 | 0.222 |  | 0.198 | 0.176 | 0.222 |
| Reason: Multimorbidity | 0.306 | 0.297 | 0.316 |  | 0.304 | 0.294 | 0.314 |  | 0.304 | 0.295 | 0.314 |  | 0.304 | 0.294 | 0.313 |  | 0.304 | 0.294 | 0.314 |
| Reason: Nervous system diseases | 0.180 | 0.172 | 0.187 |  | 0.179 | 0.172 | 0.186 |  | 0.179 | 0.172 | 0.186 |  | 0.179 | 0.172 | 0.186 |  | 0.179 | 0.172 | 0.186 |
| Reason: Others | 0.104 | 0.094 | 0.115 |  | 0.103 | 0.093 | 0.114 |  | 0.103 | 0.093 | 0.114 |  | 0.103 | 0.093 | 0.114 |  | 0.103 | 0.093 | 0.114 |
| Reason: Psychiatric disorders | 0.373 | 0.336 | 0.413 |  | 0.388 | 0.350 | 0.430 |  | 0.388 | 0.350 | 0.430 |  | 0.387 | 0.349 | 0.429 |  | 0.388 | 0.349 | 0.430 |
| Reason: Specific diseases | 0.190 | 0.183 | 0.198 |  | 0.189 | 0.181 | 0.196 |  | 0.189 | 0.181 | 0.196 |  | 0.188 | 0.181 | 0.196 |  | 0.189 | 0.181 | 0.196 |
| Basis: advanced | 0.200 | 0.181 | 0.219 |  | 0.200 | 0.182 | 0.220 |  | 0.200 | 0.182 | 0.219 |  | 0.201 | 0.182 | 0.220 |  | 0.201 | 0.182 | 0.220 |
| Suffering: both | 2.013 | 1.962 | 2.067 |  | 2.016 | 1.964 | 2.069 |  | 2.017 | 1.965 | 2.070 |  | 2.013 | 1.961 | 2.066 |  | 2.013 | 1.961 | 2.066 |
| Suffering: mental | 0.629 | 0.586 | 0.674 |  | 0.623 | 0.580 | 0.668 |  | 0.623 | 0.581 | 0.668 |  | 0.625 | 0.583 | 0.670 |  | 0.625 | 0.583 | 0.670 |
| Term: Short term | 1.897 | 1.834 | 1.962 |  | 1.905 | 1.842 | 1.971 |  | 1.906 | 1.843 | 1.972 |  | 1.909 | 1.846 | 1.974 |  | 1.909 | 1.846 | 1.975 |
| Place: Hospital | 0.823 | 0.803 | 0.843 |  | 0.822 | 0.802 | 0.841 |  | 0.822 | 0.803 | 0.842 |  | 0.824 | 0.804 | 0.844 |  | 0.824 | 0.804 | 0.843 |
| Place: Nursing home | 0.461 | 0.446 | 0.477 |  | 0.457 | 0.442 | 0.473 |  | 0.457 | 0.442 | 0.473 |  | 0.458 | 0.442 | 0.474 |  | 0.458 | 0.442 | 0.474 |
| Place: Other | 0.132 | 0.122 | 0.143 |  | 0.130 | 0.120 | 0.141 |  | 0.130 | 0.120 | 0.141 |  | 0.130 | 0.120 | 0.141 |  | 0.130 | 0.120 | 0.141 |
| Place: Palliative care | 0.192 | 0.178 | 0.206 |  | 0.190 | 0.177 | 0.205 |  | 0.190 | 0.177 | 0.204 |  | 0.191 | 0.177 | 0.205 |  | 0.190 | 0.177 | 0.205 |

| Model 2. Interaction between year and reason for euthanasia | | | | | | | | | | | | | | | | | | | |
| --- | --- | --- | --- | --- | --- | --- | --- | --- | --- | --- | --- | --- | --- | --- | --- | --- | --- | --- | --- |
|  | Incidence rate ratio | | |  | Prevalence ratio | | |  | Prevalence ratio (sensitivity check) | | | | | | | | | | |
|  |  |  |  |  | Includng Brussels | | |  | 1. excluding Brussels | | |  | 2. baseline values, including Brussels | | |  | 3. baseline values, excluding Brussels | | |
|  | RR | 2.5 % | 97.5 % |  | PR | 2.5 % | 97.5 % |  | PR | 2.5 % | 97.5 % |  | PR | 2.5 % | 97.5 % |  | PR | 2.5 % | 97.5 % |
| (Intercept) | 0.612 | 0.571 | 0.655 |  | 0.000 | 0.000 | 0.000 |  | 0.000 | 0.000 | 0.000 |  | 0.000 | 0.000 | 0.000 |  | 0.000 | 0.000 | 0.000 |
| year | 1.070 | 1.067 | 1.073 |  | 1.054 | 1.051 | 1.056 |  | 1.054 | 1.051 | 1.056 |  | 1.070 | 1.068 | 1.073 |  | 1.070 | 1.068 | 1.073 |
| Reason: Dementia | 0.285 | 0.191 | 0.418 |  | 0.282 | 0.188 | 0.412 |  | 0.282 | 0.188 | 0.413 |  | 0.290 | 0.194 | 0.424 |  | 0.290 | 0.194 | 0.424 |
| Reason: Multimorbidity | 0.193 | 0.172 | 0.215 |  | 0.194 | 0.174 | 0.217 |  | 0.195 | 0.175 | 0.218 |  | 0.186 | 0.167 | 0.208 |  | 0.186 | 0.167 | 0.208 |
| Reason: Nervous system diseases | 0.233 | 0.205 | 0.266 |  | 0.228 | 0.199 | 0.259 |  | 0.228 | 0.199 | 0.259 |  | 0.233 | 0.204 | 0.265 |  | 0.233 | 0.205 | 0.266 |
| Reason: Others | 0.321 | 0.225 | 0.450 |  | 0.312 | 0.219 | 0.439 |  | 0.312 | 0.218 | 0.438 |  | 0.318 | 0.222 | 0.447 |  | 0.318 | 0.223 | 0.447 |
| Reason: Psychiatric disorders | 0.863 | 0.622 | 1.185 |  | 0.857 | 0.617 | 1.178 |  | 0.851 | 0.613 | 1.170 |  | 0.921 | 0.662 | 1.269 |  | 0.922 | 0.663 | 1.270 |
| Reason: Specific diseases | 0.254 | 0.223 | 0.289 |  | 0.253 | 0.222 | 0.288 |  | 0.254 | 0.223 | 0.289 |  | 0.246 | 0.216 | 0.280 |  | 0.246 | 0.216 | 0.280 |
| LanguageNL | 2.453 | 2.391 | 2.517 |  | 1.513 | 1.474 | 1.552 |  | 1.246 | 1.215 | 1.279 |  | 1.658 | 1.616 | 1.701 |  | 1.333 | 1.299 | 1.368 |
| Age group: 15-29 | 0.212 | 0.176 | 0.253 |  | 0.165 | 0.137 | 0.197 |  | 0.167 | 0.139 | 0.199 |  | 0.142 | 0.118 | 0.169 |  | 0.144 | 0.120 | 0.172 |
| Age group: 30-39 | 0.293 | 0.264 | 0.324 |  | 0.313 | 0.282 | 0.347 |  | 0.321 | 0.289 | 0.356 |  | 0.248 | 0.223 | 0.275 |  | 0.252 | 0.227 | 0.279 |
| Age group: 40-49 | 0.485 | 0.453 | 0.518 |  | 0.487 | 0.455 | 0.520 |  | 0.493 | 0.461 | 0.527 |  | 0.410 | 0.383 | 0.438 |  | 0.411 | 0.384 | 0.439 |
| Age group: 60-69 | 1.640 | 1.574 | 1.709 |  | 2.009 | 1.928 | 2.093 |  | 1.992 | 1.912 | 2.076 |  | 2.158 | 2.071 | 2.248 |  | 2.159 | 2.072 | 2.250 |
| Age group: 70-79 | 1.886 | 1.813 | 1.963 |  | 3.280 | 3.153 | 3.413 |  | 3.250 | 3.124 | 3.382 |  | 2.977 | 2.861 | 3.097 |  | 2.977 | 2.862 | 3.098 |
| Age group: 80-89 | 1.804 | 1.734 | 1.878 |  | 5.673 | 5.451 | 5.906 |  | 5.646 | 5.425 | 5.878 |  | 7.319 | 7.032 | 7.620 |  | 7.425 | 7.134 | 7.730 |
| Age group: 90+ | 0.837 | 0.797 | 0.879 |  | 13.167 | 12.529 | 13.836 |  | 13.235 | 12.594 | 13.909 |  | 19.666 | 18.712 | 20.667 |  | 20.227 | 19.246 | 21.258 |
| Gender: male | 1.046 | 1.024 | 1.069 |  | 1.364 | 1.335 | 1.394 |  | 1.360 | 1.331 | 1.390 |  | 1.539 | 1.506 | 1.573 |  | 1.534 | 1.500 | 1.568 |
| Basis: advanced | 0.200 | 0.181 | 0.219 |  | 0.200 | 0.182 | 0.219 |  | 0.200 | 0.181 | 0.219 |  | 0.200 | 0.182 | 0.220 |  | 0.201 | 0.182 | 0.220 |
| Suffering: both | 2.016 | 1.964 | 2.069 |  | 2.018 | 1.966 | 2.071 |  | 2.019 | 1.967 | 2.072 |  | 2.015 | 1.964 | 2.068 |  | 2.016 | 1.964 | 2.069 |
| Suffering: mental | 0.627 | 0.584 | 0.672 |  | 0.621 | 0.579 | 0.666 |  | 0.622 | 0.579 | 0.667 |  | 0.624 | 0.581 | 0.669 |  | 0.624 | 0.581 | 0.669 |
| Term: Short term | 1.904 | 1.841 | 1.969 |  | 1.912 | 1.849 | 1.978 |  | 1.913 | 1.850 | 1.979 |  | 1.916 | 1.853 | 1.982 |  | 1.917 | 1.853 | 1.983 |
| Place: Hospital | 0.822 | 0.803 | 0.842 |  | 0.821 | 0.802 | 0.841 |  | 0.822 | 0.802 | 0.841 |  | 0.823 | 0.804 | 0.843 |  | 0.823 | 0.804 | 0.843 |
| Place: Nursing home | 0.461 | 0.445 | 0.476 |  | 0.457 | 0.441 | 0.472 |  | 0.457 | 0.441 | 0.473 |  | 0.457 | 0.442 | 0.473 |  | 0.457 | 0.442 | 0.473 |
| Place: Other | 0.132 | 0.121 | 0.143 |  | 0.130 | 0.120 | 0.141 |  | 0.130 | 0.120 | 0.141 |  | 0.130 | 0.120 | 0.140 |  | 0.130 | 0.120 | 0.141 |
| Place: Palliative care | 0.191 | 0.177 | 0.205 |  | 0.189 | 0.176 | 0.203 |  | 0.189 | 0.176 | 0.203 |  | 0.189 | 0.176 | 0.203 |  | 0.189 | 0.176 | 0.203 |
| Reason: Dementia * Year | 0.977 | 0.954 | 1.002 |  | 0.978 | 0.955 | 1.002 |  | 0.978 | 0.955 | 1.002 |  | 0.976 | 0.954 | 1.000 |  | 0.976 | 0.954 | 1.000 |
| Reason: Multimorbidity * Year | 1.030 | 1.023 | 1.037 |  | 1.029 | 1.022 | 1.036 |  | 1.029 | 1.022 | 1.035 |  | 1.032 | 1.025 | 1.038 |  | 1.032 | 1.025 | 1.038 |
| Reason: Nervous system diseases * Year | 0.983 | 0.975 | 0.991 |  | 0.984 | 0.976 | 0.992 |  | 0.984 | 0.976 | 0.992 |  | 0.983 | 0.975 | 0.991 |  | 0.983 | 0.975 | 0.991 |
| Reason: Others * Year | 0.931 | 0.910 | 0.952 |  | 0.932 | 0.911 | 0.953 |  | 0.932 | 0.911 | 0.953 |  | 0.930 | 0.910 | 0.952 |  | 0.930 | 0.910 | 0.952 |
| Reason: Psychiatric disorders * Year | 0.946 | 0.926 | 0.966 |  | 0.949 | 0.929 | 0.969 |  | 0.949 | 0.930 | 0.970 |  | 0.944 | 0.925 | 0.965 |  | 0.944 | 0.925 | 0.965 |
| Reason: Specific diseases * Year | 0.981 | 0.973 | 0.989 |  | 0.981 | 0.973 | 0.989 |  | 0.981 | 0.973 | 0.989 |  | 0.982 | 0.974 | 0.991 |  | 0.982 | 0.974 | 0.991 |

| Model 3. Interaction between year and basis for euthanasia | | | | | | | | | | | | | | | | | | | |
| --- | --- | --- | --- | --- | --- | --- | --- | --- | --- | --- | --- | --- | --- | --- | --- | --- | --- | --- | --- |
|  | Incidence rate ratio | | |  | Prevalence ratio | | |  | Prevalence ratio (sensitivity check) | | | | | | | | | | |
|  |  |  |  |  | Includng Brussels | | |  | 1. excluding Brussels | | |  | 2. baseline values, including Brussels | | |  | 3. baseline values, excluding Brussels | | |
|  | RR | 2.5 % | 97.5 % |  | PR | 2.5 % | 97.5 % |  | PR | 2.5 % | 97.5 % |  | PR | 2.5 % | 97.5 % |  | PR | 2.5 % | 97.5 % |
| (Intercept) | 0.616 | 0.576 | 0.658 |  | 0.000 | 0.000 | 0.000 |  | 0.000 | 0.000 | 0.000 |  | 0.000 | 0.000 | 0.000 |  | 0.000 | 0.000 | 0.000 |
| year | 1.070 | 1.068 | 1.072 |  | 1.054 | 1.052 | 1.056 |  | 1.054 | 1.051 | 1.056 |  | 1.071 | 1.069 | 1.073 |  | 1.071 | 1.069 | 1.073 |
| Basis: advanced | 0.296 | 0.231 | 0.376 |  | 0.288 | 0.225 | 0.365 |  | 0.287 | 0.225 | 0.365 |  | 0.303 | 0.237 | 0.384 |  | 0.304 | 0.237 | 0.386 |
| Reason: Dementia | 0.199 | 0.177 | 0.223 |  | 0.198 | 0.176 | 0.222 |  | 0.198 | 0.176 | 0.222 |  | 0.198 | 0.176 | 0.222 |  | 0.198 | 0.176 | 0.222 |
| Reason: Multimorbidity | 0.306 | 0.297 | 0.316 |  | 0.304 | 0.294 | 0.314 |  | 0.304 | 0.294 | 0.314 |  | 0.303 | 0.294 | 0.313 |  | 0.304 | 0.294 | 0.314 |
| Reason: Nervous system diseases | 0.180 | 0.172 | 0.187 |  | 0.179 | 0.172 | 0.186 |  | 0.179 | 0.172 | 0.186 |  | 0.179 | 0.172 | 0.186 |  | 0.179 | 0.172 | 0.186 |
| Reason: Others | 0.104 | 0.094 | 0.115 |  | 0.103 | 0.093 | 0.114 |  | 0.103 | 0.093 | 0.114 |  | 0.103 | 0.093 | 0.114 |  | 0.103 | 0.093 | 0.114 |
| Reason: Psychiatric disorders | 0.373 | 0.336 | 0.413 |  | 0.389 | 0.350 | 0.430 |  | 0.388 | 0.350 | 0.430 |  | 0.387 | 0.349 | 0.429 |  | 0.388 | 0.350 | 0.430 |
| Reason: Specific diseases | 0.190 | 0.183 | 0.198 |  | 0.189 | 0.181 | 0.196 |  | 0.189 | 0.181 | 0.197 |  | 0.188 | 0.181 | 0.196 |  | 0.189 | 0.181 | 0.196 |
| LanguageNL | 2.450 | 2.388 | 2.513 |  | 1.511 | 1.473 | 1.550 |  | 1.245 | 1.213 | 1.277 |  | 1.655 | 1.613 | 1.698 |  | 1.331 | 1.297 | 1.365 |
| Age group: 15-29 | 0.212 | 0.176 | 0.253 |  | 0.165 | 0.138 | 0.197 |  | 0.168 | 0.139 | 0.200 |  | 0.142 | 0.118 | 0.170 |  | 0.144 | 0.120 | 0.172 |
| Age group: 30-39 | 0.292 | 0.263 | 0.324 |  | 0.313 | 0.281 | 0.346 |  | 0.320 | 0.288 | 0.355 |  | 0.248 | 0.223 | 0.274 |  | 0.251 | 0.226 | 0.278 |
| Age group: 40-49 | 0.487 | 0.455 | 0.520 |  | 0.489 | 0.457 | 0.523 |  | 0.495 | 0.463 | 0.530 |  | 0.412 | 0.385 | 0.440 |  | 0.413 | 0.386 | 0.441 |
| Age group: 60-69 | 1.633 | 1.567 | 1.701 |  | 2.000 | 1.920 | 2.084 |  | 1.983 | 1.904 | 2.067 |  | 2.149 | 2.063 | 2.240 |  | 2.150 | 2.064 | 2.241 |
| Age group: 70-79 | 1.879 | 1.806 | 1.955 |  | 3.267 | 3.141 | 3.400 |  | 3.237 | 3.112 | 3.369 |  | 2.966 | 2.851 | 3.086 |  | 2.967 | 2.852 | 3.087 |
| Age group: 80-89 | 1.801 | 1.730 | 1.874 |  | 5.661 | 5.440 | 5.893 |  | 5.634 | 5.414 | 5.865 |  | 7.307 | 7.020 | 7.607 |  | 7.412 | 7.121 | 7.717 |
| Age group: 90+ | 0.837 | 0.797 | 0.880 |  | 13.194 | 12.555 | 13.865 |  | 13.263 | 12.621 | 13.937 |  | 19.681 | 18.726 | 20.683 |  | 20.242 | 19.261 | 21.273 |
| Gender: male | 1.046 | 1.024 | 1.069 |  | 1.364 | 1.334 | 1.394 |  | 1.359 | 1.330 | 1.389 |  | 1.539 | 1.505 | 1.573 |  | 1.533 | 1.500 | 1.567 |
| Suffering: both | 2.009 | 1.957 | 2.062 |  | 2.011 | 1.960 | 2.064 |  | 2.013 | 1.961 | 2.066 |  | 2.008 | 1.957 | 2.061 |  | 2.008 | 1.957 | 2.061 |
| Suffering: mental | 0.627 | 0.585 | 0.673 |  | 0.622 | 0.579 | 0.666 |  | 0.622 | 0.580 | 0.667 |  | 0.624 | 0.581 | 0.669 |  | 0.624 | 0.581 | 0.669 |
| Term: Short term | 1.897 | 1.834 | 1.962 |  | 1.905 | 1.842 | 1.971 |  | 1.906 | 1.843 | 1.972 |  | 1.909 | 1.846 | 1.975 |  | 1.909 | 1.846 | 1.975 |
| Place: Hospital | 0.823 | 0.803 | 0.842 |  | 0.822 | 0.802 | 0.841 |  | 0.822 | 0.802 | 0.842 |  | 0.824 | 0.804 | 0.843 |  | 0.823 | 0.804 | 0.843 |
| Place: Nursing home | 0.461 | 0.446 | 0.477 |  | 0.457 | 0.442 | 0.473 |  | 0.457 | 0.442 | 0.473 |  | 0.458 | 0.442 | 0.473 |  | 0.458 | 0.442 | 0.473 |
| Place: Other | 0.132 | 0.122 | 0.143 |  | 0.130 | 0.120 | 0.141 |  | 0.130 | 0.120 | 0.141 |  | 0.130 | 0.120 | 0.141 |  | 0.130 | 0.120 | 0.141 |
| Place: Palliative care | 0.192 | 0.178 | 0.206 |  | 0.190 | 0.177 | 0.204 |  | 0.190 | 0.177 | 0.204 |  | 0.190 | 0.177 | 0.205 |  | 0.190 | 0.177 | 0.204 |
| Basis: advanced * Year | 0.969 | 0.952 | 0.987 |  | 0.972 | 0.954 | 0.989 |  | 0.972 | 0.954 | 0.990 |  | 0.968 | 0.950 | 0.986 |  | 0.968 | 0.950 | 0.986 |

| Model 4. Interaction between year and type of suffering | | | | | | | | | | | | | | | | | | | |
| --- | --- | --- | --- | --- | --- | --- | --- | --- | --- | --- | --- | --- | --- | --- | --- | --- | --- | --- | --- |
|  | Incidence rate ratio | | |  | Prevalence ratio | | |  | Prevalence ratio (sensitivity check) | | | | | | | | | | |
|  |  |  |  |  | Includng Brussels | | |  | 1. excluding Brussels | | |  | 2. baseline values, including Brussels | | |  | 3. baseline values, excluding Brussels | | |
|  | RR | 2.5 % | 97.5 % |  | PR | 2.5 % | 97.5 % |  | PR | 2.5 % | 97.5 % |  | PR | 2.5 % | 97.5 % |  | PR | 2.5 % | 97.5 % |
| (Intercept) | 0.735 | 0.677 | 0.798 |  | 0.000 | 0.000 | 0.000 |  | 0.000 | 0.000 | 0.000 |  | 0.000 | 0.000 | 0.000 |  | 0.000 | 0.000 | 0.000 |
| year | 1.057 | 1.052 | 1.061 |  | 1.041 | 1.036 | 1.045 |  | 1.040 | 1.036 | 1.045 |  | 1.057 | 1.053 | 1.062 |  | 1.057 | 1.053 | 1.061 |
| Suffering: both | 1.575 | 1.464 | 1.696 |  | 1.577 | 1.465 | 1.697 |  | 1.578 | 1.467 | 1.699 |  | 1.566 | 1.455 | 1.686 |  | 1.565 | 1.454 | 1.685 |
| Suffering: mental | 0.606 | 0.510 | 0.717 |  | 0.604 | 0.508 | 0.714 |  | 0.604 | 0.508 | 0.714 |  | 0.615 | 0.517 | 0.727 |  | 0.615 | 0.518 | 0.728 |
| Reason: Dementia | 0.200 | 0.178 | 0.225 |  | 0.200 | 0.177 | 0.224 |  | 0.200 | 0.177 | 0.224 |  | 0.200 | 0.177 | 0.224 |  | 0.200 | 0.178 | 0.224 |
| Reason: Multimorbidity | 0.307 | 0.297 | 0.316 |  | 0.304 | 0.295 | 0.314 |  | 0.304 | 0.295 | 0.314 |  | 0.304 | 0.294 | 0.314 |  | 0.304 | 0.294 | 0.314 |
| Reason: Nervous system diseases | 0.180 | 0.173 | 0.188 |  | 0.179 | 0.172 | 0.187 |  | 0.179 | 0.172 | 0.187 |  | 0.179 | 0.172 | 0.187 |  | 0.179 | 0.172 | 0.187 |
| Reason: Others | 0.104 | 0.093 | 0.115 |  | 0.103 | 0.093 | 0.114 |  | 0.103 | 0.093 | 0.114 |  | 0.103 | 0.092 | 0.113 |  | 0.103 | 0.092 | 0.114 |
| Reason: Psychiatric disorders | 0.376 | 0.338 | 0.416 |  | 0.391 | 0.352 | 0.433 |  | 0.391 | 0.352 | 0.433 |  | 0.390 | 0.351 | 0.433 |  | 0.391 | 0.352 | 0.433 |
| Reason: Specific diseases | 0.191 | 0.183 | 0.198 |  | 0.189 | 0.182 | 0.197 |  | 0.189 | 0.182 | 0.197 |  | 0.189 | 0.181 | 0.196 |  | 0.189 | 0.181 | 0.197 |
| LanguageNL | 2.450 | 2.388 | 2.514 |  | 1.511 | 1.473 | 1.551 |  | 1.245 | 1.213 | 1.277 |  | 1.656 | 1.614 | 1.699 |  | 1.331 | 1.297 | 1.366 |
| Age group: 15-29 | 0.213 | 0.177 | 0.253 |  | 0.166 | 0.138 | 0.198 |  | 0.168 | 0.140 | 0.200 |  | 0.143 | 0.119 | 0.170 |  | 0.145 | 0.120 | 0.172 |
| Age group: 30-39 | 0.292 | 0.263 | 0.323 |  | 0.312 | 0.281 | 0.346 |  | 0.320 | 0.288 | 0.354 |  | 0.247 | 0.222 | 0.274 |  | 0.251 | 0.226 | 0.278 |
| Age group: 40-49 | 0.487 | 0.455 | 0.521 |  | 0.490 | 0.458 | 0.524 |  | 0.496 | 0.463 | 0.530 |  | 0.412 | 0.385 | 0.440 |  | 0.413 | 0.386 | 0.442 |
| Age group: 60-69 | 1.631 | 1.566 | 1.700 |  | 1.998 | 1.918 | 2.082 |  | 1.981 | 1.901 | 2.064 |  | 2.146 | 2.060 | 2.236 |  | 2.148 | 2.061 | 2.238 |
| Age group: 70-79 | 1.881 | 1.808 | 1.957 |  | 3.270 | 3.143 | 3.403 |  | 3.240 | 3.115 | 3.372 |  | 2.969 | 2.854 | 3.089 |  | 2.970 | 2.854 | 3.090 |
| Age group: 80-89 | 1.802 | 1.731 | 1.875 |  | 5.666 | 5.444 | 5.898 |  | 5.639 | 5.418 | 5.870 |  | 7.310 | 7.023 | 7.610 |  | 7.415 | 7.124 | 7.720 |
| Age group: 90+ | 0.838 | 0.798 | 0.881 |  | 13.202 | 12.563 | 13.873 |  | 13.270 | 12.628 | 13.945 |  | 19.708 | 18.752 | 20.711 |  | 20.271 | 19.288 | 21.304 |
| Gender: male | 1.046 | 1.024 | 1.068 |  | 1.363 | 1.334 | 1.393 |  | 1.359 | 1.330 | 1.389 |  | 1.538 | 1.505 | 1.573 |  | 1.533 | 1.499 | 1.567 |
| Basis: advanced | 0.201 | 0.182 | 0.220 |  | 0.201 | 0.182 | 0.221 |  | 0.201 | 0.182 | 0.220 |  | 0.201 | 0.183 | 0.221 |  | 0.202 | 0.183 | 0.221 |
| Term: Short term | 1.902 | 1.839 | 1.968 |  | 1.910 | 1.847 | 1.976 |  | 1.911 | 1.848 | 1.977 |  | 1.914 | 1.851 | 1.980 |  | 1.915 | 1.851 | 1.980 |
| Place: Hospital | 0.822 | 0.803 | 0.842 |  | 0.821 | 0.802 | 0.841 |  | 0.821 | 0.802 | 0.841 |  | 0.823 | 0.804 | 0.843 |  | 0.823 | 0.804 | 0.843 |
| Place: Nursing home | 0.461 | 0.446 | 0.477 |  | 0.457 | 0.442 | 0.473 |  | 0.458 | 0.442 | 0.473 |  | 0.458 | 0.442 | 0.474 |  | 0.458 | 0.442 | 0.474 |
| Place: Other | 0.132 | 0.122 | 0.143 |  | 0.130 | 0.120 | 0.141 |  | 0.130 | 0.120 | 0.141 |  | 0.130 | 0.120 | 0.141 |  | 0.130 | 0.120 | 0.141 |
| Place: Palliative care | 0.191 | 0.178 | 0.205 |  | 0.190 | 0.176 | 0.204 |  | 0.190 | 0.176 | 0.204 |  | 0.190 | 0.176 | 0.204 |  | 0.190 | 0.176 | 0.204 |
| Suffering: both * Year | 1.017 | 1.012 | 1.022 |  | 1.017 | 1.012 | 1.022 |  | 1.017 | 1.012 | 1.022 |  | 1.017 | 1.013 | 1.022 |  | 1.018 | 1.013 | 1.022 |
| Suffering: mental * Year | 1.002 | 0.990 | 1.014 |  | 1.001 | 0.989 | 1.013 |  | 1.001 | 0.989 | 1.013 |  | 1.000 | 0.988 | 1.012 |  | 1.000 | 0.988 | 1.012 |

| Model 5. Interaction between year and expected term of death | | | | | | | | | | | | | | | | | | | |
| --- | --- | --- | --- | --- | --- | --- | --- | --- | --- | --- | --- | --- | --- | --- | --- | --- | --- | --- | --- |
|  | Incidence rate ratio | | |  | Prevalence ratio | | |  | Prevalence ratio (sensitivity check) | | | | | | | | | | |
|  |  |  |  |  | Includng Brussels | | |  | 1. excluding Brussels | | |  | 2. baseline values, including Brussels | | |  | 3. baseline values, excluding Brussels | | |
|  | RR | 2.5 % | 97.5 % |  | PR | 2.5 % | 97.5 % |  | PR | 2.5 % | 97.5 % |  | PR | 2.5 % | 97.5 % |  | PR | 2.5 % | 97.5 % |
| (Intercept) | 0.715 | 0.635 | 0.804 |  | 0.000 | 0.000 | 0.000 |  | 0.000 | 0.000 | 0.000 |  | 0.000 | 0.000 | 0.000 |  | 0.000 | 0.000 | 0.000 |
| year | 1.060 | 1.053 | 1.067 |  | 1.046 | 1.039 | 1.053 |  | 1.045 | 1.038 | 1.052 |  | 1.061 | 1.054 | 1.068 |  | 1.061 | 1.054 | 1.068 |
| Term: Short term | 1.620 | 1.448 | 1.814 |  | 1.675 | 1.497 | 1.876 |  | 1.674 | 1.497 | 1.875 |  | 1.627 | 1.454 | 1.822 |  | 1.629 | 1.456 | 1.824 |
| Reason: Dementia | 0.200 | 0.177 | 0.224 |  | 0.199 | 0.177 | 0.223 |  | 0.199 | 0.177 | 0.223 |  | 0.199 | 0.176 | 0.223 |  | 0.199 | 0.177 | 0.223 |
| Reason: Multimorbidity | 0.306 | 0.297 | 0.316 |  | 0.304 | 0.294 | 0.314 |  | 0.304 | 0.295 | 0.314 |  | 0.304 | 0.294 | 0.313 |  | 0.304 | 0.294 | 0.314 |
| Reason: Nervous system diseases | 0.180 | 0.172 | 0.187 |  | 0.179 | 0.171 | 0.186 |  | 0.179 | 0.171 | 0.186 |  | 0.179 | 0.171 | 0.186 |  | 0.179 | 0.171 | 0.186 |
| Reason: Others | 0.104 | 0.094 | 0.115 |  | 0.103 | 0.093 | 0.114 |  | 0.103 | 0.093 | 0.114 |  | 0.103 | 0.093 | 0.114 |  | 0.103 | 0.093 | 0.114 |
| Reason: Psychiatric disorders | 0.372 | 0.335 | 0.412 |  | 0.388 | 0.349 | 0.430 |  | 0.388 | 0.349 | 0.429 |  | 0.387 | 0.348 | 0.428 |  | 0.387 | 0.349 | 0.429 |
| Reason: Specific diseases | 0.190 | 0.183 | 0.198 |  | 0.189 | 0.181 | 0.196 |  | 0.189 | 0.181 | 0.196 |  | 0.188 | 0.181 | 0.196 |  | 0.188 | 0.181 | 0.196 |
| LanguageNL | 2.450 | 2.388 | 2.514 |  | 1.511 | 1.473 | 1.550 |  | 1.245 | 1.213 | 1.277 |  | 1.655 | 1.613 | 1.698 |  | 1.331 | 1.297 | 1.366 |
| Age group: 15-29 | 0.212 | 0.176 | 0.252 |  | 0.165 | 0.137 | 0.197 |  | 0.167 | 0.139 | 0.199 |  | 0.142 | 0.118 | 0.169 |  | 0.144 | 0.120 | 0.172 |
| Age group: 30-39 | 0.291 | 0.262 | 0.323 |  | 0.312 | 0.281 | 0.346 |  | 0.320 | 0.288 | 0.354 |  | 0.247 | 0.222 | 0.273 |  | 0.251 | 0.226 | 0.278 |
| Age group: 40-49 | 0.486 | 0.454 | 0.519 |  | 0.489 | 0.457 | 0.523 |  | 0.495 | 0.463 | 0.529 |  | 0.411 | 0.384 | 0.439 |  | 0.412 | 0.385 | 0.440 |
| Age group: 60-69 | 1.632 | 1.566 | 1.700 |  | 1.999 | 1.918 | 2.083 |  | 1.982 | 1.902 | 2.065 |  | 2.147 | 2.061 | 2.237 |  | 2.148 | 2.062 | 2.239 |
| Age group: 70-79 | 1.876 | 1.803 | 1.952 |  | 3.264 | 3.137 | 3.396 |  | 3.234 | 3.109 | 3.365 |  | 2.961 | 2.847 | 3.082 |  | 2.962 | 2.848 | 3.083 |
| Age group: 80-89 | 1.796 | 1.726 | 1.870 |  | 5.648 | 5.427 | 5.880 |  | 5.621 | 5.401 | 5.852 |  | 7.287 | 7.001 | 7.587 |  | 7.392 | 7.102 | 7.696 |
| Age group: 90+ | 0.836 | 0.796 | 0.878 |  | 13.170 | 12.532 | 13.839 |  | 13.238 | 12.597 | 13.911 |  | 19.628 | 18.676 | 20.628 |  | 20.188 | 19.209 | 21.216 |
| Gender: male | 1.046 | 1.024 | 1.069 |  | 1.364 | 1.334 | 1.394 |  | 1.359 | 1.330 | 1.389 |  | 1.539 | 1.505 | 1.573 |  | 1.533 | 1.500 | 1.567 |
| Basis: advanced | 0.200 | 0.182 | 0.219 |  | 0.200 | 0.182 | 0.220 |  | 0.200 | 0.182 | 0.220 |  | 0.201 | 0.182 | 0.220 |  | 0.201 | 0.183 | 0.221 |
| Suffering: both | 2.013 | 1.962 | 2.066 |  | 2.015 | 1.964 | 2.068 |  | 2.016 | 1.965 | 2.070 |  | 2.012 | 1.961 | 2.065 |  | 2.013 | 1.961 | 2.066 |
| Suffering: mental | 0.629 | 0.586 | 0.674 |  | 0.623 | 0.580 | 0.668 |  | 0.623 | 0.581 | 0.668 |  | 0.625 | 0.583 | 0.670 |  | 0.625 | 0.583 | 0.670 |
| Place: Hospital | 0.822 | 0.803 | 0.842 |  | 0.821 | 0.802 | 0.841 |  | 0.822 | 0.802 | 0.841 |  | 0.823 | 0.804 | 0.843 |  | 0.823 | 0.804 | 0.843 |
| Place: Nursing home | 0.461 | 0.446 | 0.477 |  | 0.457 | 0.442 | 0.473 |  | 0.457 | 0.442 | 0.473 |  | 0.458 | 0.442 | 0.473 |  | 0.458 | 0.442 | 0.473 |
| Place: Other | 0.132 | 0.122 | 0.143 |  | 0.130 | 0.120 | 0.141 |  | 0.130 | 0.120 | 0.141 |  | 0.130 | 0.120 | 0.141 |  | 0.130 | 0.120 | 0.141 |
| Place: Palliative care | 0.192 | 0.178 | 0.206 |  | 0.190 | 0.177 | 0.204 |  | 0.190 | 0.177 | 0.204 |  | 0.190 | 0.177 | 0.205 |  | 0.190 | 0.177 | 0.205 |
| Term: Short term * Year | 1.010 | 1.003 | 1.017 |  | 1.008 | 1.001 | 1.015 |  | 1.008 | 1.001 | 1.015 |  | 1.010 | 1.003 | 1.017 |  | 1.010 | 1.003 | 1.017 |

| Model 6. Interaction between year and place of death | | | | | | | | | | | | | | | | | | | |
| --- | --- | --- | --- | --- | --- | --- | --- | --- | --- | --- | --- | --- | --- | --- | --- | --- | --- | --- | --- |
|  | Incidence rate ratio | | |  | Prevalence ratio | | |  | Prevalence ratio (sensitivity check) | | | | | | | | | | |
|  |  |  |  |  | Includng Brussels | | |  | 1. excluding Brussels | | |  | 2. baseline values, including Brussels | | |  | 3. baseline values, excluding Brussels | | |
|  | RR | 2.5 % | 97.5 % |  | PR | 2.5 % | 97.5 % |  | PR | 2.5 % | 97.5 % |  | PR | 2.5 % | 97.5 % |  | PR | 2.5 % | 97.5 % |
| (Intercept) | 0.504 | 0.468 | 0.543 |  | 0.000 | 0.000 | 0.000 |  | 0.000 | 0.000 | 0.000 |  | 0.000 | 0.000 | 0.000 |  | 0.000 | 0.000 | 0.000 |
| year | 1.084 | 1.081 | 1.088 |  | 1.068 | 1.065 | 1.071 |  | 1.068 | 1.064 | 1.071 |  | 1.085 | 1.081 | 1.088 |  | 1.085 | 1.081 | 1.088 |
| Place: Hospital | 1.309 | 1.223 | 1.401 |  | 1.303 | 1.218 | 1.395 |  | 1.304 | 1.218 | 1.395 |  | 1.302 | 1.217 | 1.393 |  | 1.302 | 1.216 | 1.393 |
| Place: Nursing home | 0.459 | 0.406 | 0.518 |  | 0.464 | 0.410 | 0.524 |  | 0.466 | 0.412 | 0.526 |  | 0.441 | 0.390 | 0.498 |  | 0.440 | 0.390 | 0.497 |
| Place: Other | 0.236 | 0.183 | 0.302 |  | 0.230 | 0.178 | 0.295 |  | 0.230 | 0.177 | 0.295 |  | 0.228 | 0.176 | 0.292 |  | 0.228 | 0.176 | 0.292 |
| Place: Palliative care | 0.133 | 0.093 | 0.184 |  | 0.130 | 0.091 | 0.180 |  | 0.130 | 0.091 | 0.180 |  | 0.133 | 0.093 | 0.185 |  | 0.133 | 0.094 | 0.185 |
| Reason: Dementia | 0.200 | 0.178 | 0.224 |  | 0.199 | 0.177 | 0.224 |  | 0.200 | 0.177 | 0.224 |  | 0.199 | 0.177 | 0.223 |  | 0.199 | 0.177 | 0.224 |
| Reason: Multimorbidity | 0.306 | 0.297 | 0.316 |  | 0.304 | 0.294 | 0.314 |  | 0.304 | 0.294 | 0.314 |  | 0.304 | 0.294 | 0.313 |  | 0.304 | 0.294 | 0.314 |
| Reason: Nervous system diseases | 0.180 | 0.172 | 0.187 |  | 0.179 | 0.172 | 0.186 |  | 0.179 | 0.172 | 0.186 |  | 0.179 | 0.172 | 0.186 |  | 0.179 | 0.172 | 0.186 |
| Reason: Others | 0.105 | 0.094 | 0.116 |  | 0.104 | 0.093 | 0.115 |  | 0.104 | 0.094 | 0.115 |  | 0.103 | 0.093 | 0.115 |  | 0.104 | 0.093 | 0.115 |
| Reason: Psychiatric disorders | 0.372 | 0.336 | 0.412 |  | 0.388 | 0.349 | 0.430 |  | 0.388 | 0.349 | 0.429 |  | 0.387 | 0.348 | 0.428 |  | 0.388 | 0.349 | 0.429 |
| Reason: Specific diseases | 0.190 | 0.183 | 0.198 |  | 0.189 | 0.181 | 0.196 |  | 0.189 | 0.181 | 0.196 |  | 0.189 | 0.181 | 0.196 |  | 0.189 | 0.181 | 0.196 |
| LanguageNL | 2.456 | 2.394 | 2.520 |  | 1.515 | 1.476 | 1.554 |  | 1.248 | 1.216 | 1.281 |  | 1.659 | 1.617 | 1.703 |  | 1.334 | 1.300 | 1.369 |
| Agegroup15-29 | 0.212 | 0.176 | 0.253 |  | 0.165 | 0.137 | 0.197 |  | 0.168 | 0.139 | 0.200 |  | 0.142 | 0.118 | 0.169 |  | 0.144 | 0.120 | 0.171 |
| Agegroup30-39 | 0.290 | 0.261 | 0.321 |  | 0.310 | 0.279 | 0.344 |  | 0.318 | 0.286 | 0.352 |  | 0.246 | 0.221 | 0.272 |  | 0.250 | 0.225 | 0.276 |
| Agegroup40-49 | 0.485 | 0.453 | 0.519 |  | 0.487 | 0.455 | 0.521 |  | 0.493 | 0.461 | 0.527 |  | 0.410 | 0.383 | 0.438 |  | 0.411 | 0.384 | 0.439 |
| Agegroup60-69 | 1.628 | 1.563 | 1.696 |  | 1.995 | 1.915 | 2.079 |  | 1.978 | 1.899 | 2.061 |  | 2.143 | 2.057 | 2.233 |  | 2.144 | 2.058 | 2.234 |
| Agegroup70-79 | 1.880 | 1.808 | 1.957 |  | 3.269 | 3.143 | 3.402 |  | 3.240 | 3.114 | 3.371 |  | 2.969 | 2.854 | 3.090 |  | 2.970 | 2.855 | 3.091 |
| Agegroup80-89 | 1.802 | 1.731 | 1.875 |  | 5.667 | 5.445 | 5.900 |  | 5.640 | 5.419 | 5.872 |  | 7.313 | 7.026 | 7.613 |  | 7.418 | 7.127 | 7.723 |
| Agegroup90+ | 0.837 | 0.797 | 0.879 |  | 13.184 | 12.546 | 13.854 |  | 13.253 | 12.611 | 13.927 |  | 19.680 | 18.726 | 20.682 |  | 20.241 | 19.259 | 21.271 |
| Gendermale | 1.045 | 1.023 | 1.068 |  | 1.362 | 1.333 | 1.392 |  | 1.358 | 1.329 | 1.388 |  | 1.537 | 1.504 | 1.571 |  | 1.531 | 1.498 | 1.565 |
| basisadvanced | 0.199 | 0.181 | 0.219 |  | 0.200 | 0.181 | 0.219 |  | 0.199 | 0.181 | 0.219 |  | 0.200 | 0.182 | 0.220 |  | 0.200 | 0.182 | 0.220 |
| sufferingboth | 2.016 | 1.964 | 2.069 |  | 2.018 | 1.966 | 2.071 |  | 2.019 | 1.968 | 2.072 |  | 2.015 | 1.963 | 2.068 |  | 2.015 | 1.964 | 2.069 |
| sufferingmental | 0.621 | 0.578 | 0.665 |  | 0.615 | 0.573 | 0.659 |  | 0.615 | 0.573 | 0.660 |  | 0.617 | 0.575 | 0.661 |  | 0.617 | 0.575 | 0.661 |
| termShort term | 1.905 | 1.842 | 1.970 |  | 1.913 | 1.849 | 1.978 |  | 1.914 | 1.850 | 1.979 |  | 1.916 | 1.852 | 1.982 |  | 1.916 | 1.853 | 1.982 |
| year:PlaceHospital | 0.968 | 0.963 | 0.972 |  | 0.968 | 0.964 | 0.972 |  | 0.968 | 0.964 | 0.972 |  | 0.968 | 0.964 | 0.973 |  | 0.968 | 0.964 | 0.973 |
| year:PlaceNursing home | 0.999 | 0.992 | 1.007 |  | 0.998 | 0.991 | 1.006 |  | 0.998 | 0.990 | 1.005 |  | 1.002 | 0.994 | 1.009 |  | 1.002 | 0.994 | 1.009 |
| year:PlaceOther | 0.962 | 0.946 | 0.978 |  | 0.962 | 0.947 | 0.978 |  | 0.962 | 0.947 | 0.978 |  | 0.963 | 0.947 | 0.979 |  | 0.963 | 0.947 | 0.979 |
| year:PlacePalliative care | 1.018 | 1.000 | 1.037 |  | 1.019 | 1.001 | 1.038 |  | 1.019 | 1.001 | 1.038 |  | 1.018 | 1.000 | 1.037 |  | 1.017 | 1.000 | 1.037 |

| Model 7. Interaction between year and age group | | | | | | | | | | | | | | | | | | | |
| --- | --- | --- | --- | --- | --- | --- | --- | --- | --- | --- | --- | --- | --- | --- | --- | --- | --- | --- | --- |
|  | Incidence rate ratio | | |  | Prevalence ratio | | |  | Prevalence ratio (sensitivity check) | | | | | | | | | | |
|  |  |  |  |  | Includng Brussels | | |  | 1. excluding Brussels | | |  | 2. baseline values, including Brussels | | |  | 3. baseline values, excluding Brussels | | |
|  | RR | 2.5 % | 97.5 % |  | PR | 2.5 % | 97.5 % |  | PR | 2.5 % | 97.5 % |  | PR | 2.5 % | 97.5 % |  | PR | 2.5 % | 97.5 % |
| (Intercept) | 0.929 | 0.841 | 1.025 |  | 0.000 | 0.000 | 0.000 |  | 0.000 | 0.000 | 0.000 |  | 0.000 | 0.000 | 0.000 |  | 0.000 | 0.000 | 0.000 |
| year | 1.038 | 1.032 | 1.044 |  | 1.028 | 1.022 | 1.035 |  | 1.029 | 1.023 | 1.035 |  | 1.039 | 1.033 | 1.045 |  | 1.039 | 1.033 | 1.045 |
| Age group: 15-29 | 0.253 | 0.148 | 0.417 |  | 0.174 | 0.102 | 0.287 |  | 0.177 | 0.103 | 0.291 |  | 0.167 | 0.097 | 0.274 |  | 0.169 | 0.098 | 0.278 |
| Age group: 30-39 | 0.319 | 0.245 | 0.412 |  | 0.310 | 0.238 | 0.399 |  | 0.316 | 0.243 | 0.408 |  | 0.275 | 0.211 | 0.355 |  | 0.279 | 0.214 | 0.360 |
| Age group: 40-49 | 0.539 | 0.456 | 0.635 |  | 0.445 | 0.377 | 0.526 |  | 0.446 | 0.377 | 0.526 |  | 0.455 | 0.385 | 0.536 |  | 0.456 | 0.386 | 0.538 |
| Age group: 60-69 | 1.288 | 1.156 | 1.435 |  | 1.804 | 1.619 | 2.012 |  | 1.813 | 1.626 | 2.021 |  | 1.687 | 1.515 | 1.879 |  | 1.687 | 1.515 | 1.879 |
| Age group: 70-79 | 1.165 | 1.049 | 1.294 |  | 2.058 | 1.854 | 2.287 |  | 2.076 | 1.869 | 2.307 |  | 1.845 | 1.661 | 2.050 |  | 1.846 | 1.663 | 2.051 |
| Age group: 80-89 | 0.883 | 0.793 | 0.985 |  | 2.991 | 2.679 | 3.340 |  | 3.034 | 2.718 | 3.389 |  | 3.611 | 3.241 | 4.026 |  | 3.651 | 3.276 | 4.070 |
| Age group: 90+ | 0.356 | 0.302 | 0.419 |  | 10.399 | 8.814 | 12.251 |  | 10.689 | 9.060 | 12.591 |  | 7.954 | 6.748 | 9.361 |  | 8.079 | 6.855 | 9.507 |
| Gendermale | 1.044 | 1.022 | 1.066 |  | 1.360 | 1.330 | 1.390 |  | 1.356 | 1.326 | 1.385 |  | 1.536 | 1.502 | 1.570 |  | 1.530 | 1.497 | 1.564 |
| LanguageNL | 2.460 | 2.398 | 2.524 |  | 1.512 | 1.473 | 1.551 |  | 1.246 | 1.214 | 1.278 |  | 1.662 | 1.620 | 1.706 |  | 1.336 | 1.302 | 1.371 |
| Reason: Dementia | 0.198 | 0.176 | 0.222 |  | 0.198 | 0.175 | 0.222 |  | 0.198 | 0.176 | 0.222 |  | 0.197 | 0.175 | 0.221 |  | 0.198 | 0.175 | 0.222 |
| Reason: Multimorbidity | 0.305 | 0.296 | 0.315 |  | 0.303 | 0.294 | 0.313 |  | 0.303 | 0.294 | 0.313 |  | 0.302 | 0.293 | 0.312 |  | 0.303 | 0.293 | 0.312 |
| Reason: Nervous system diseases | 0.179 | 0.172 | 0.187 |  | 0.178 | 0.171 | 0.186 |  | 0.178 | 0.171 | 0.186 |  | 0.178 | 0.171 | 0.186 |  | 0.178 | 0.171 | 0.186 |
| Reason: Others | 0.104 | 0.093 | 0.115 |  | 0.103 | 0.092 | 0.114 |  | 0.103 | 0.093 | 0.114 |  | 0.103 | 0.092 | 0.113 |  | 0.103 | 0.092 | 0.114 |
| Reason: Psychiatric disorders | 0.377 | 0.340 | 0.418 |  | 0.393 | 0.354 | 0.435 |  | 0.393 | 0.354 | 0.435 |  | 0.392 | 0.353 | 0.434 |  | 0.392 | 0.353 | 0.435 |
| Reason: Specific diseases | 0.190 | 0.182 | 0.198 |  | 0.188 | 0.181 | 0.196 |  | 0.189 | 0.181 | 0.196 |  | 0.188 | 0.181 | 0.196 |  | 0.188 | 0.181 | 0.196 |
| Basis: advanced | 0.196 | 0.178 | 0.215 |  | 0.198 | 0.180 | 0.217 |  | 0.198 | 0.180 | 0.217 |  | 0.197 | 0.179 | 0.217 |  | 0.198 | 0.179 | 0.217 |
| Suffering: both | 2.024 | 1.972 | 2.077 |  | 2.023 | 1.972 | 2.077 |  | 2.024 | 1.973 | 2.078 |  | 2.024 | 1.972 | 2.077 |  | 2.024 | 1.973 | 2.078 |
| Suffering: mental | 0.629 | 0.586 | 0.674 |  | 0.622 | 0.580 | 0.667 |  | 0.622 | 0.580 | 0.667 |  | 0.626 | 0.583 | 0.671 |  | 0.626 | 0.583 | 0.671 |
| Term: Short term | 1.892 | 1.830 | 1.957 |  | 1.902 | 1.839 | 1.967 |  | 1.903 | 1.840 | 1.968 |  | 1.904 | 1.841 | 1.969 |  | 1.904 | 1.841 | 1.969 |
| Place: Hospital | 0.822 | 0.803 | 0.842 |  | 0.821 | 0.802 | 0.841 |  | 0.821 | 0.802 | 0.841 |  | 0.823 | 0.804 | 0.843 |  | 0.823 | 0.804 | 0.843 |
| Place: Nursing home | 0.461 | 0.446 | 0.477 |  | 0.458 | 0.442 | 0.474 |  | 0.458 | 0.442 | 0.474 |  | 0.458 | 0.443 | 0.474 |  | 0.458 | 0.443 | 0.474 |
| Place: Other | 0.132 | 0.121 | 0.143 |  | 0.130 | 0.119 | 0.140 |  | 0.130 | 0.119 | 0.140 |  | 0.130 | 0.119 | 0.140 |  | 0.130 | 0.119 | 0.140 |
| Place: Palliative care | 0.190 | 0.177 | 0.204 |  | 0.189 | 0.176 | 0.203 |  | 0.189 | 0.176 | 0.203 |  | 0.189 | 0.176 | 0.203 |  | 0.189 | 0.175 | 0.203 |
| Age group: 15-29 * Year | 0.989 | 0.956 | 1.025 |  | 0.997 | 0.964 | 1.033 |  | 0.997 | 0.963 | 1.033 |  | 0.990 | 0.957 | 1.026 |  | 0.991 | 0.957 | 1.026 |
| Age group: 30-39 * Year | 0.993 | 0.975 | 1.011 |  | 1.000 | 0.982 | 1.018 |  | 1.000 | 0.982 | 1.019 |  | 0.991 | 0.973 | 1.010 |  | 0.992 | 0.974 | 1.010 |
| Age group: 40-49 * Year | 0.992 | 0.980 | 1.003 |  | 1.006 | 0.994 | 1.018 |  | 1.007 | 0.995 | 1.019 |  | 0.992 | 0.980 | 1.004 |  | 0.992 | 0.980 | 1.003 |
| Age group: 60-69 * Year | 1.018 | 1.011 | 1.026 |  | 1.009 | 1.001 | 1.016 |  | 1.008 | 1.000 | 1.015 |  | 1.018 | 1.011 | 1.026 |  | 1.018 | 1.011 | 1.026 |
| Age group: 70-79 * Year | 1.035 | 1.028 | 1.043 |  | 1.034 | 1.027 | 1.041 |  | 1.033 | 1.025 | 1.040 |  | 1.035 | 1.028 | 1.042 |  | 1.035 | 1.028 | 1.042 |
| Age group: 80-89 * Year | 1.052 | 1.044 | 1.059 |  | 1.046 | 1.038 | 1.054 |  | 1.045 | 1.037 | 1.052 |  | 1.051 | 1.043 | 1.059 |  | 1.051 | 1.044 | 1.059 |
| Age group: 90+ * Year | 1.059 | 1.049 | 1.070 |  | 1.019 | 1.008 | 1.029 |  | 1.017 | 1.007 | 1.028 |  | 1.063 | 1.052 | 1.074 |  | 1.064 | 1.053 | 1.075 |

| Model 8. Interaction between year and gender | | | | | | | | | | | | | | | | | | | |
| --- | --- | --- | --- | --- | --- | --- | --- | --- | --- | --- | --- | --- | --- | --- | --- | --- | --- | --- | --- |
|  | Incidence rate ratio | | |  | Prevalence ratio | | |  | Prevalence ratio (sensitivity check) | | | | | | | | | | |
|  |  |  |  |  | Includng Brussels | | |  | 1. excluding Brussels | | |  | 2. baseline values, including Brussels | | |  | 3. baseline values, excluding Brussels | | |
|  | RR | 2.5 % | 97.5 % |  | PR | 2.5 % | 97.5 % |  | PR | 2.5 % | 97.5 % |  | PR | 2.5 % | 97.5 % |  | PR | 2.5 % | 97.5 % |
| (Intercept) | 0.589 | 0.548 | 0.634 |  | 0.000 | 0.000 | 0.000 |  | 0.000 | 0.000 | 0.000 |  | 0.000 | 0.000 | 0.000 |  | 0.000 | 0.000 | 0.000 |
| year | 1.073 | 1.070 | 1.076 |  | 1.057 | 1.054 | 1.060 |  | 1.057 | 1.054 | 1.060 |  | 1.071 | 1.068 | 1.074 |  | 1.071 | 1.068 | 1.074 |
| Gender: male | 1.150 | 1.079 | 1.226 |  | 1.508 | 1.414 | 1.608 |  | 1.500 | 1.407 | 1.599 |  | 1.565 | 1.468 | 1.669 |  | 1.562 | 1.465 | 1.666 |
| Age group: 15-29 | 0.212 | 0.176 | 0.253 |  | 0.165 | 0.137 | 0.197 |  | 0.168 | 0.139 | 0.200 |  | 0.142 | 0.118 | 0.170 |  | 0.144 | 0.120 | 0.172 |
| Age group: 30-39 | 0.292 | 0.263 | 0.323 |  | 0.312 | 0.281 | 0.346 |  | 0.320 | 0.288 | 0.355 |  | 0.247 | 0.223 | 0.274 |  | 0.251 | 0.226 | 0.278 |
| Age group: 40-49 | 0.486 | 0.455 | 0.520 |  | 0.489 | 0.457 | 0.523 |  | 0.495 | 0.463 | 0.529 |  | 0.411 | 0.385 | 0.440 |  | 0.412 | 0.386 | 0.441 |
| Age group: 60-69 | 1.631 | 1.566 | 1.700 |  | 1.999 | 1.919 | 2.083 |  | 1.982 | 1.903 | 2.066 |  | 2.147 | 2.061 | 2.237 |  | 2.148 | 2.062 | 2.239 |
| Age group: 70-79 | 1.878 | 1.805 | 1.954 |  | 3.268 | 3.141 | 3.400 |  | 3.238 | 3.113 | 3.370 |  | 2.964 | 2.850 | 3.085 |  | 2.965 | 2.851 | 3.086 |
| Age group: 80-89 | 1.798 | 1.728 | 1.872 |  | 5.659 | 5.438 | 5.891 |  | 5.633 | 5.412 | 5.864 |  | 7.294 | 7.008 | 7.594 |  | 7.400 | 7.109 | 7.704 |
| Age group: 90+ | 0.837 | 0.797 | 0.879 |  | 13.172 | 12.534 | 13.842 |  | 13.241 | 12.600 | 13.915 |  | 19.656 | 18.703 | 20.657 |  | 20.216 | 19.236 | 21.246 |
| LanguageNL | 2.451 | 2.389 | 2.515 |  | 1.512 | 1.473 | 1.551 |  | 1.245 | 1.214 | 1.278 |  | 1.656 | 1.614 | 1.699 |  | 1.332 | 1.298 | 1.366 |
| Reason: Dementia | 0.199 | 0.177 | 0.224 |  | 0.199 | 0.176 | 0.223 |  | 0.199 | 0.177 | 0.223 |  | 0.198 | 0.176 | 0.222 |  | 0.198 | 0.176 | 0.222 |
| Reason: Multimorbidity | 0.306 | 0.297 | 0.316 |  | 0.304 | 0.294 | 0.314 |  | 0.304 | 0.294 | 0.314 |  | 0.304 | 0.294 | 0.313 |  | 0.304 | 0.294 | 0.314 |
| Reason: Nervous system diseases | 0.180 | 0.173 | 0.187 |  | 0.179 | 0.172 | 0.186 |  | 0.179 | 0.172 | 0.186 |  | 0.179 | 0.172 | 0.186 |  | 0.179 | 0.172 | 0.186 |
| Reason: Others | 0.104 | 0.094 | 0.115 |  | 0.103 | 0.093 | 0.114 |  | 0.103 | 0.093 | 0.114 |  | 0.103 | 0.093 | 0.114 |  | 0.103 | 0.093 | 0.114 |
| Reason: Psychiatric disorders | 0.372 | 0.335 | 0.412 |  | 0.388 | 0.349 | 0.429 |  | 0.388 | 0.349 | 0.429 |  | 0.387 | 0.349 | 0.429 |  | 0.388 | 0.349 | 0.430 |
| Reason: Specific diseases | 0.190 | 0.183 | 0.198 |  | 0.189 | 0.181 | 0.196 |  | 0.189 | 0.181 | 0.196 |  | 0.188 | 0.181 | 0.196 |  | 0.189 | 0.181 | 0.196 |
| Basis: advanced | 0.200 | 0.182 | 0.220 |  | 0.200 | 0.182 | 0.220 |  | 0.200 | 0.182 | 0.220 |  | 0.201 | 0.182 | 0.220 |  | 0.201 | 0.183 | 0.221 |
| Suffering: both | 2.014 | 1.963 | 2.067 |  | 2.016 | 1.965 | 2.069 |  | 2.017 | 1.966 | 2.071 |  | 2.013 | 1.961 | 2.066 |  | 2.013 | 1.962 | 2.066 |
| Suffering: mental | 0.630 | 0.587 | 0.676 |  | 0.624 | 0.582 | 0.669 |  | 0.625 | 0.582 | 0.670 |  | 0.626 | 0.583 | 0.671 |  | 0.626 | 0.583 | 0.671 |
| Term: Short term | 1.897 | 1.834 | 1.962 |  | 1.905 | 1.842 | 1.971 |  | 1.906 | 1.843 | 1.972 |  | 1.909 | 1.846 | 1.974 |  | 1.909 | 1.846 | 1.975 |
| Place: Hospital | 0.823 | 0.803 | 0.842 |  | 0.822 | 0.802 | 0.841 |  | 0.822 | 0.802 | 0.842 |  | 0.824 | 0.804 | 0.844 |  | 0.824 | 0.804 | 0.843 |
| Place: Nursing home | 0.462 | 0.446 | 0.478 |  | 0.458 | 0.442 | 0.473 |  | 0.458 | 0.442 | 0.474 |  | 0.458 | 0.442 | 0.474 |  | 0.458 | 0.442 | 0.474 |
| Place: Other | 0.132 | 0.122 | 0.143 |  | 0.130 | 0.120 | 0.141 |  | 0.130 | 0.120 | 0.141 |  | 0.130 | 0.120 | 0.141 |  | 0.130 | 0.120 | 0.141 |
| Place: Palliative care | 0.192 | 0.178 | 0.206 |  | 0.190 | 0.177 | 0.205 |  | 0.190 | 0.177 | 0.205 |  | 0.191 | 0.177 | 0.205 |  | 0.191 | 0.177 | 0.205 |
| Gender: male. * Year | 0.994 | 0.989 | 0.998 |  | 0.993 | 0.989 | 0.997 |  | 0.993 | 0.989 | 0.997 |  | 0.999 | 0.995 | 1.003 |  | 0.999 | 0.995 | 1.003 |

| Model 9. Interaction between year and language/region | | | | | | | | | | | | | | | | | | | |
| --- | --- | --- | --- | --- | --- | --- | --- | --- | --- | --- | --- | --- | --- | --- | --- | --- | --- | --- | --- |
|  | Incidence rate ratio | | |  | Prevalence ratio | | |  | Prevalence ratio (sensitivity check) | | | | | | | | | | |
|  |  |  |  |  | Includng Brussels | | |  | 1. excluding Brussels | | |  | 2. baseline values, including Brussels | | |  | 3. baseline values, excluding Brussels | | |
|  | RR | 2.5 % | 97.5 % |  | PR | 2.5 % | 97.5 % |  | PR | 2.5 % | 97.5 % |  | PR | 2.5 % | 97.5 % |  | PR | 2.5 % | 97.5 % |
| (Intercept) | 0.542 | 0.494 | 0.594 |  | 0.000 | 0.000 | 0.000 |  | 0.000 | 0.000 | 0.000 |  | 0.000 | 0.000 | 0.000 |  | 0.000 | 0.000 | 0.000 |
| year | 1.079 | 1.074 | 1.084 |  | 1.068 | 1.063 | 1.073 |  | 1.067 | 1.062 | 1.072 |  | 1.079 | 1.074 | 1.084 |  | 1.080 | 1.075 | 1.085 |
| LanguageNL | 2.884 | 2.655 | 3.136 |  | 1.961 | 1.805 | 2.132 |  | 1.593 | 1.466 | 1.733 |  | 1.938 | 1.783 | 2.108 |  | 1.569 | 1.444 | 1.706 |
| Age group: 15-29 | 0.212 | 0.176 | 0.253 |  | 0.165 | 0.137 | 0.197 |  | 0.168 | 0.139 | 0.200 |  | 0.142 | 0.118 | 0.170 |  | 0.144 | 0.120 | 0.172 |
| Age group: 30-39 | 0.292 | 0.262 | 0.323 |  | 0.312 | 0.281 | 0.345 |  | 0.320 | 0.288 | 0.354 |  | 0.247 | 0.222 | 0.274 |  | 0.251 | 0.226 | 0.278 |
| Age group: 40-49 | 0.486 | 0.454 | 0.519 |  | 0.488 | 0.456 | 0.521 |  | 0.494 | 0.462 | 0.528 |  | 0.411 | 0.384 | 0.439 |  | 0.412 | 0.385 | 0.440 |
| Age group: 60-69 | 1.633 | 1.567 | 1.701 |  | 2.000 | 1.919 | 2.084 |  | 1.982 | 1.903 | 2.065 |  | 2.148 | 2.062 | 2.238 |  | 2.149 | 2.063 | 2.240 |
| Age group: 70-79 | 1.877 | 1.805 | 1.954 |  | 3.263 | 3.137 | 3.396 |  | 3.233 | 3.108 | 3.364 |  | 2.964 | 2.849 | 3.084 |  | 2.965 | 2.850 | 3.085 |
| Age group: 80-89 | 1.798 | 1.727 | 1.871 |  | 5.658 | 5.437 | 5.890 |  | 5.630 | 5.409 | 5.860 |  | 7.292 | 7.006 | 7.592 |  | 7.397 | 7.107 | 7.701 |
| Age group: 90+ | 0.836 | 0.796 | 0.878 |  | 13.187 | 12.548 | 13.857 |  | 13.253 | 12.612 | 13.927 |  | 19.644 | 18.692 | 20.644 |  | 20.207 | 19.227 | 21.236 |
| Gender: male | 1.046 | 1.023 | 1.068 |  | 1.363 | 1.334 | 1.393 |  | 1.359 | 1.329 | 1.389 |  | 1.538 | 1.505 | 1.572 |  | 1.532 | 1.499 | 1.566 |
| Reason: Dementia | 0.199 | 0.177 | 0.223 |  | 0.198 | 0.176 | 0.222 |  | 0.198 | 0.176 | 0.223 |  | 0.198 | 0.176 | 0.222 |  | 0.198 | 0.176 | 0.222 |
| Reason: Multimorbidity | 0.306 | 0.296 | 0.316 |  | 0.304 | 0.294 | 0.313 |  | 0.304 | 0.294 | 0.314 |  | 0.303 | 0.294 | 0.313 |  | 0.304 | 0.294 | 0.313 |
| Reason: Nervous system diseases | 0.180 | 0.172 | 0.187 |  | 0.179 | 0.171 | 0.186 |  | 0.179 | 0.171 | 0.186 |  | 0.179 | 0.171 | 0.186 |  | 0.179 | 0.172 | 0.186 |
| Reason: Others | 0.104 | 0.094 | 0.116 |  | 0.104 | 0.093 | 0.115 |  | 0.104 | 0.093 | 0.115 |  | 0.103 | 0.093 | 0.114 |  | 0.103 | 0.093 | 0.114 |
| Reason: Psychiatric disorders | 0.373 | 0.336 | 0.413 |  | 0.388 | 0.350 | 0.430 |  | 0.388 | 0.350 | 0.430 |  | 0.387 | 0.349 | 0.429 |  | 0.388 | 0.349 | 0.430 |
| Reason: Specific diseases | 0.190 | 0.183 | 0.198 |  | 0.188 | 0.181 | 0.196 |  | 0.189 | 0.181 | 0.196 |  | 0.188 | 0.181 | 0.196 |  | 0.188 | 0.181 | 0.196 |
| Basis: advanced | 0.199 | 0.181 | 0.219 |  | 0.199 | 0.181 | 0.219 |  | 0.199 | 0.181 | 0.219 |  | 0.200 | 0.182 | 0.220 |  | 0.200 | 0.182 | 0.220 |
| Suffering: both | 2.015 | 1.963 | 2.068 |  | 2.017 | 1.966 | 2.070 |  | 2.018 | 1.967 | 2.071 |  | 2.014 | 1.962 | 2.067 |  | 2.014 | 1.962 | 2.067 |
| Suffering: mental | 0.628 | 0.586 | 0.674 |  | 0.622 | 0.580 | 0.667 |  | 0.623 | 0.580 | 0.668 |  | 0.625 | 0.582 | 0.670 |  | 0.625 | 0.582 | 0.670 |
| Term: Short term | 1.898 | 1.835 | 1.963 |  | 1.907 | 1.844 | 1.973 |  | 1.908 | 1.845 | 1.974 |  | 1.910 | 1.847 | 1.976 |  | 1.910 | 1.847 | 1.976 |
| Place: Hospital | 0.823 | 0.804 | 0.843 |  | 0.822 | 0.803 | 0.842 |  | 0.822 | 0.803 | 0.842 |  | 0.824 | 0.805 | 0.844 |  | 0.824 | 0.805 | 0.844 |
| Place: Nursing home | 0.461 | 0.446 | 0.477 |  | 0.457 | 0.442 | 0.473 |  | 0.457 | 0.442 | 0.473 |  | 0.458 | 0.442 | 0.474 |  | 0.458 | 0.442 | 0.474 |
| Place: Other | 0.132 | 0.122 | 0.143 |  | 0.130 | 0.120 | 0.141 |  | 0.130 | 0.120 | 0.141 |  | 0.130 | 0.120 | 0.141 |  | 0.130 | 0.120 | 0.141 |
| Place: Palliative care | 0.192 | 0.178 | 0.206 |  | 0.190 | 0.176 | 0.204 |  | 0.190 | 0.176 | 0.204 |  | 0.190 | 0.177 | 0.204 |  | 0.190 | 0.177 | 0.204 |
| year:LanguageNL | 0.989 | 0.984 | 0.994 |  | 0.983 | 0.978 | 0.988 |  | 0.984 | 0.979 | 0.989 |  | 0.990 | 0.985 | 0.995 |  | 0.989 | 0.984 | 0.994 |

### Supplementary file S4. Marginal effects in the fully adjusted model

#### Model 1. Marginal effects of year

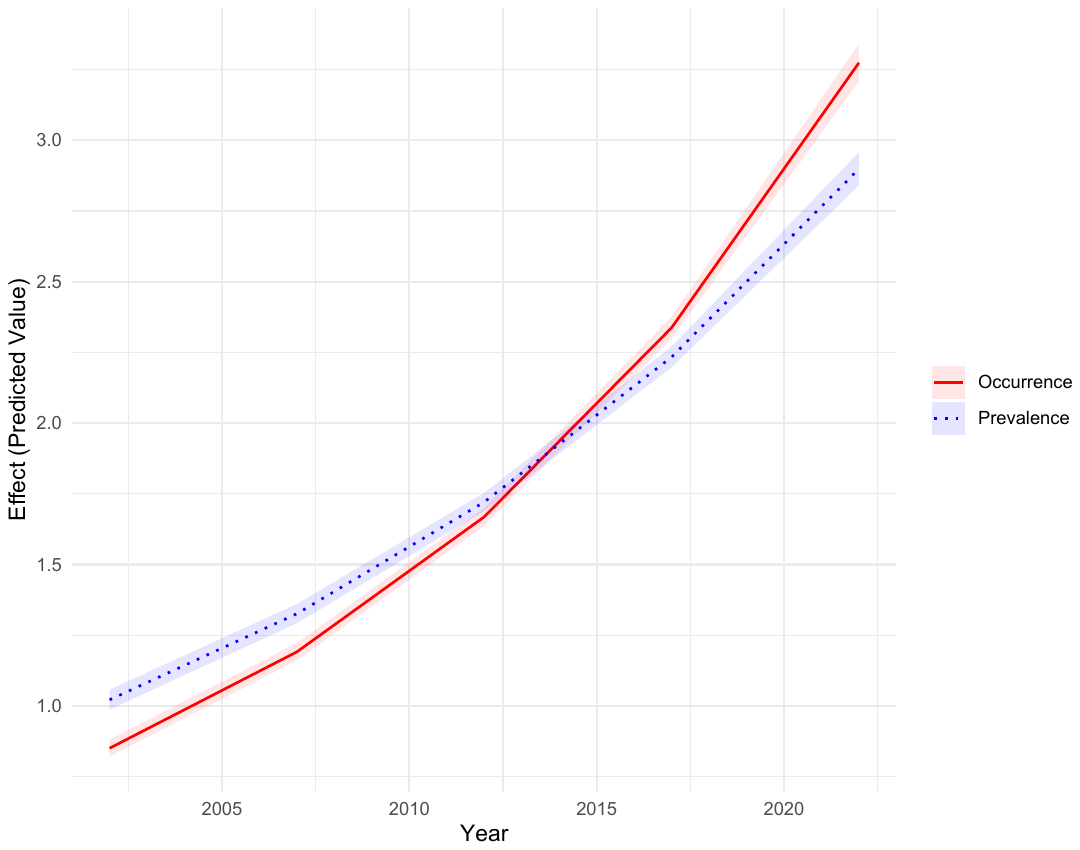

#### Model 2. Marginal effects of the interaction between age group and year

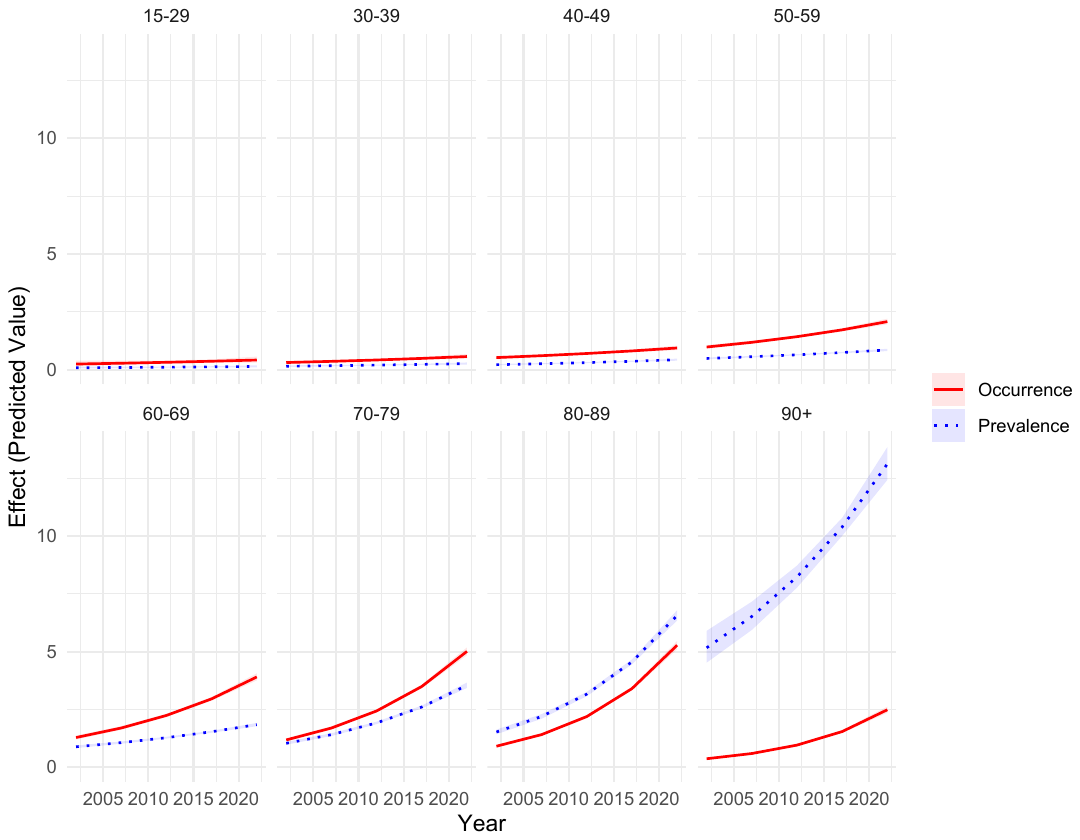

#### Model 3. Marginal effects of the interaction between gender and year

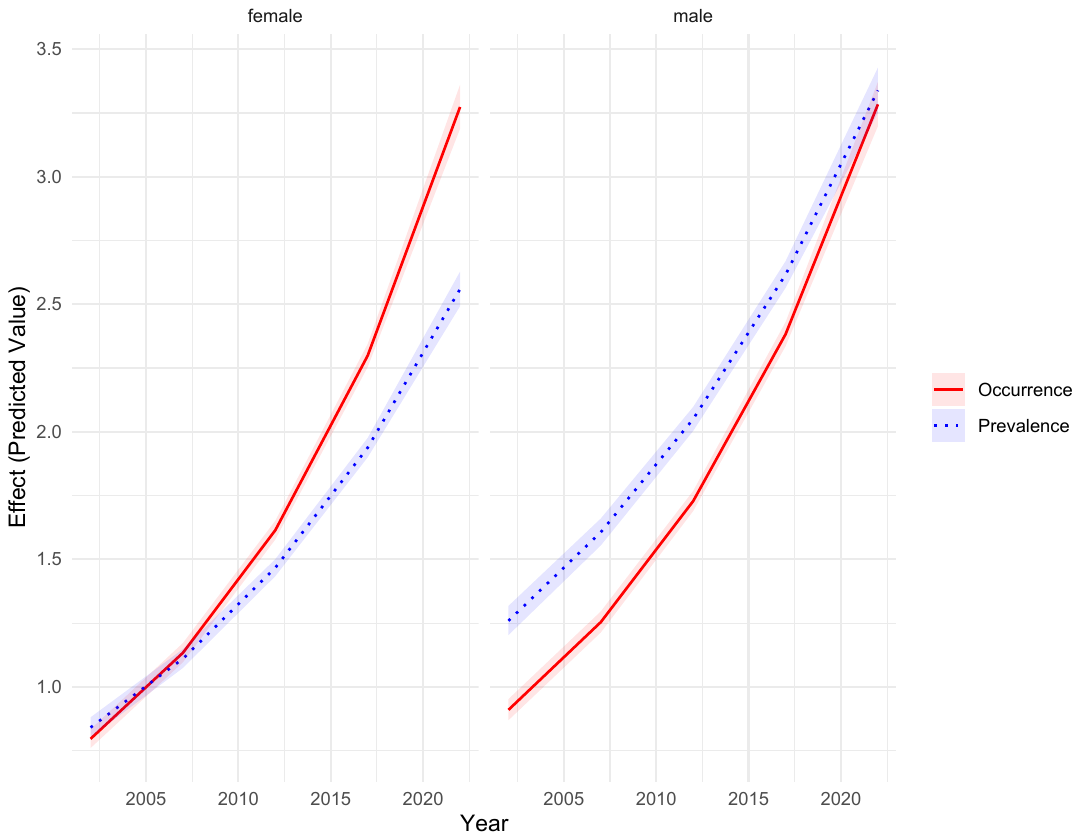

#### Model 4. Marginal effects of the interaction between language/region and year

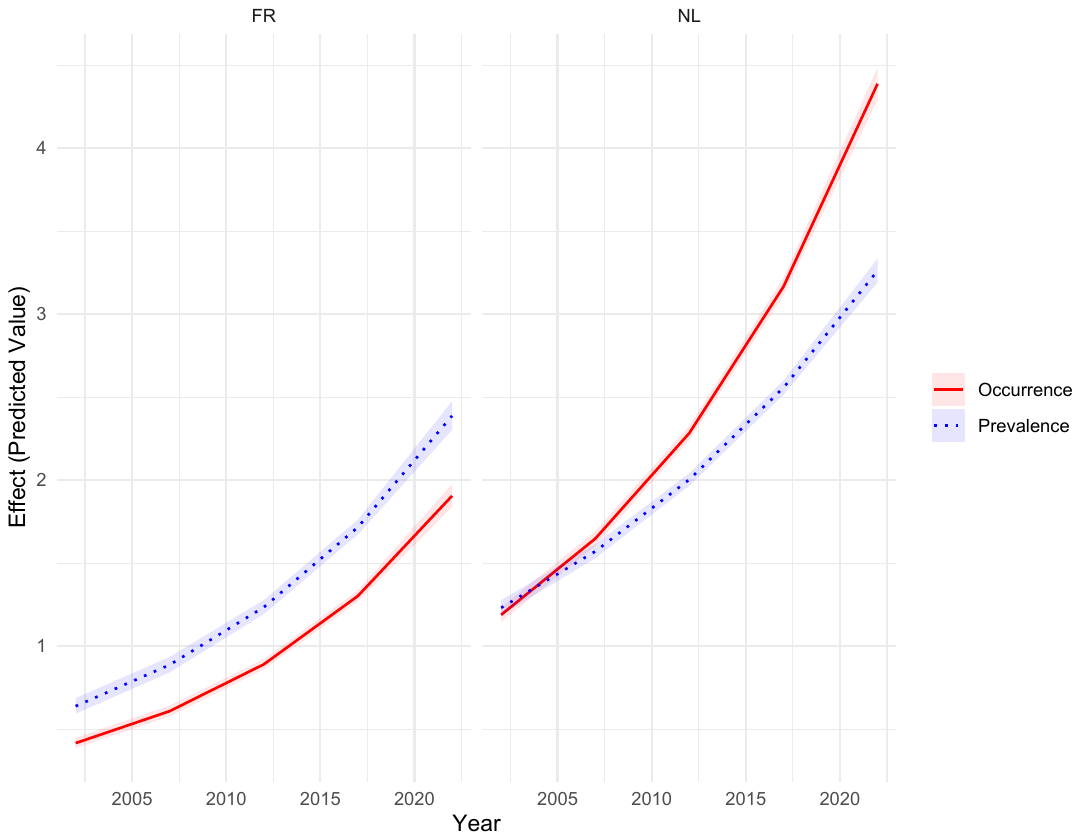

#### Model 5. Marginal effects of the interaction between reason for euthanasia and year

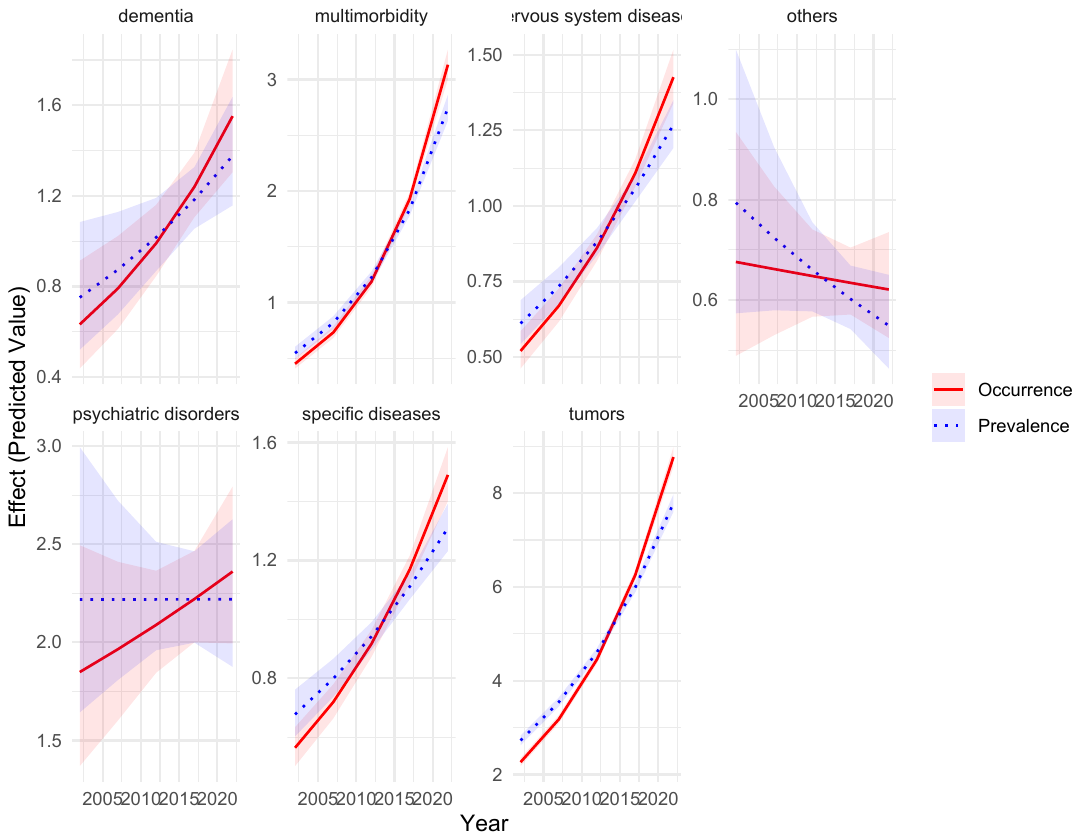

#### Model 6. Marginal effects of the interaction between basis of euthanasia and year

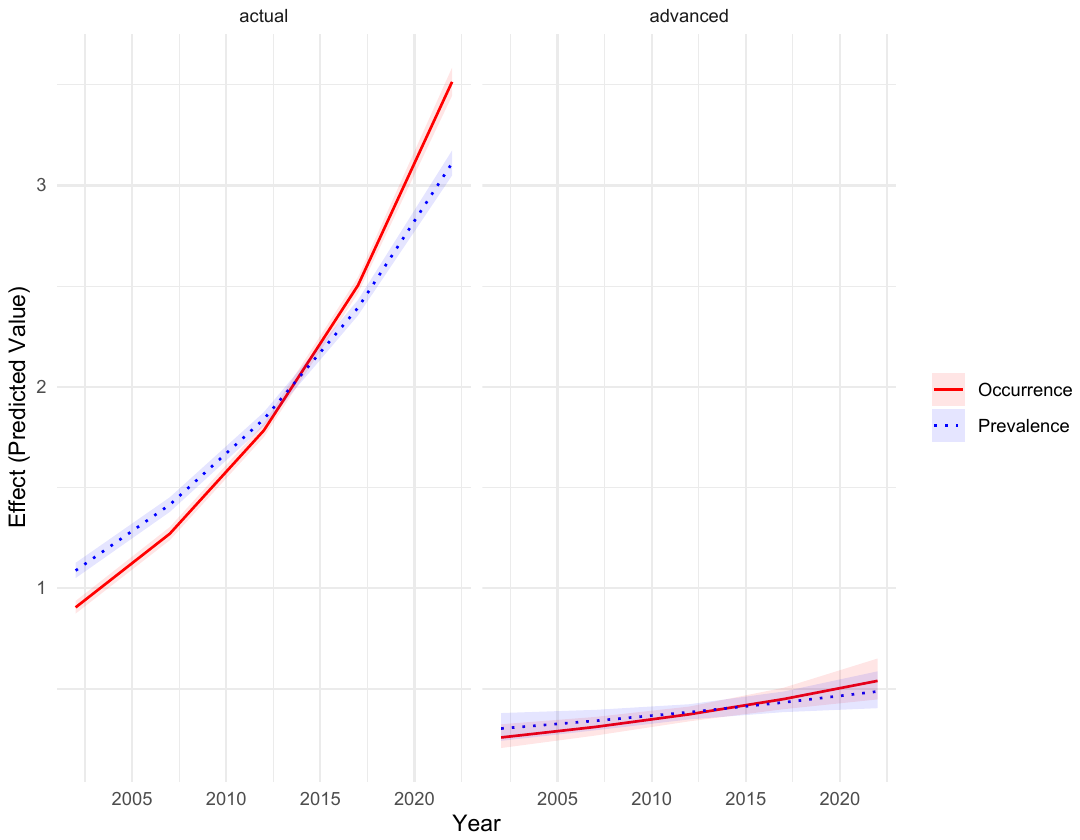

#### Model 7. Marginal effects of the interaction between type of suffering and year

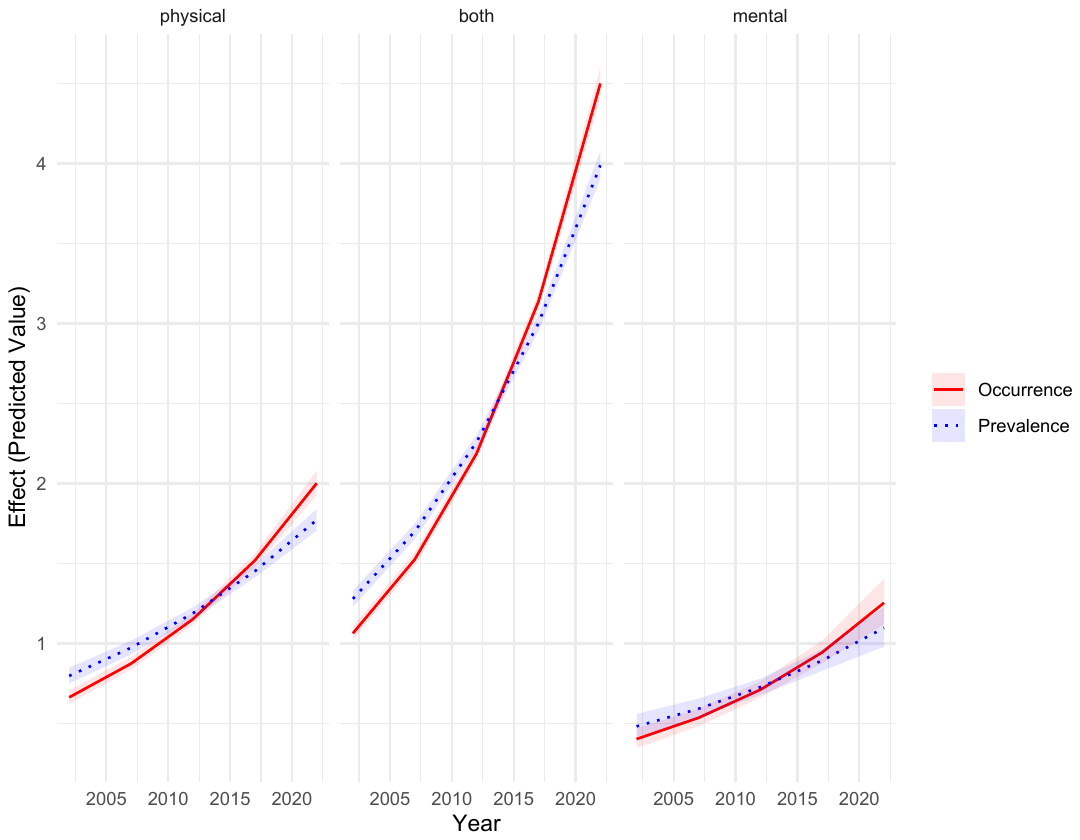

#### Model 8. Marginal effects of the interaction between term of death and year

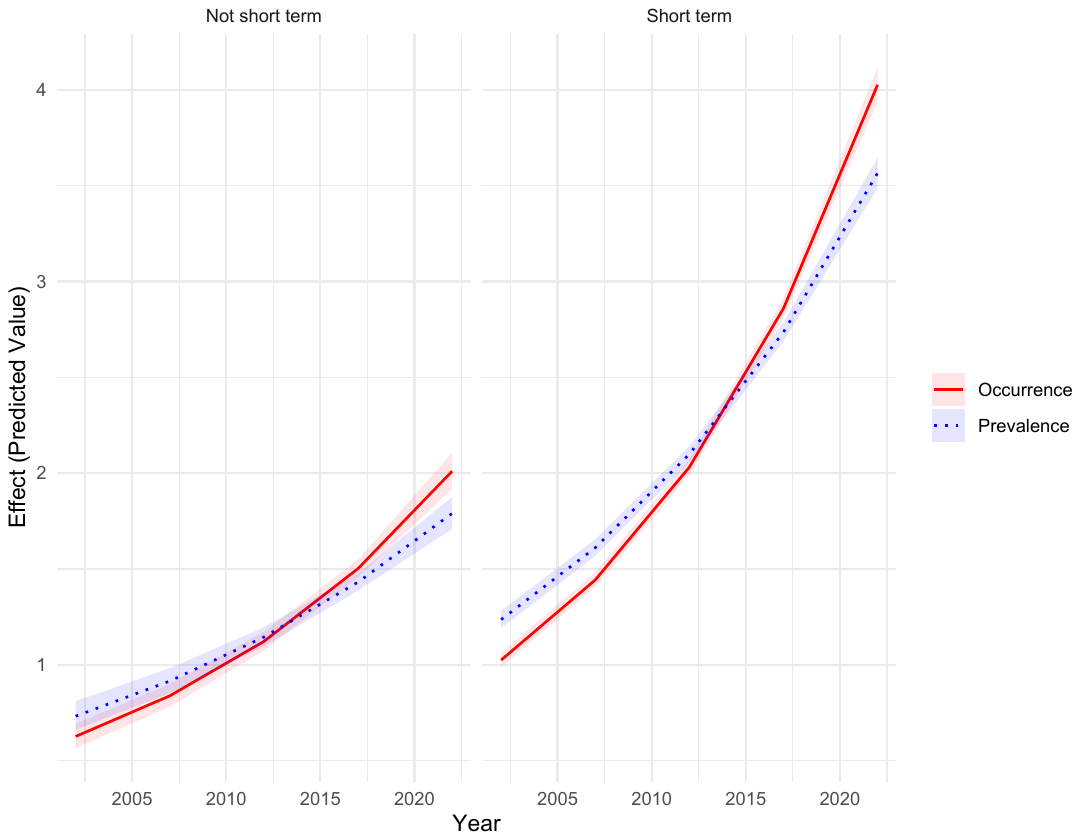

#### Model 9. Marginal effects of the interaction between place of death and year

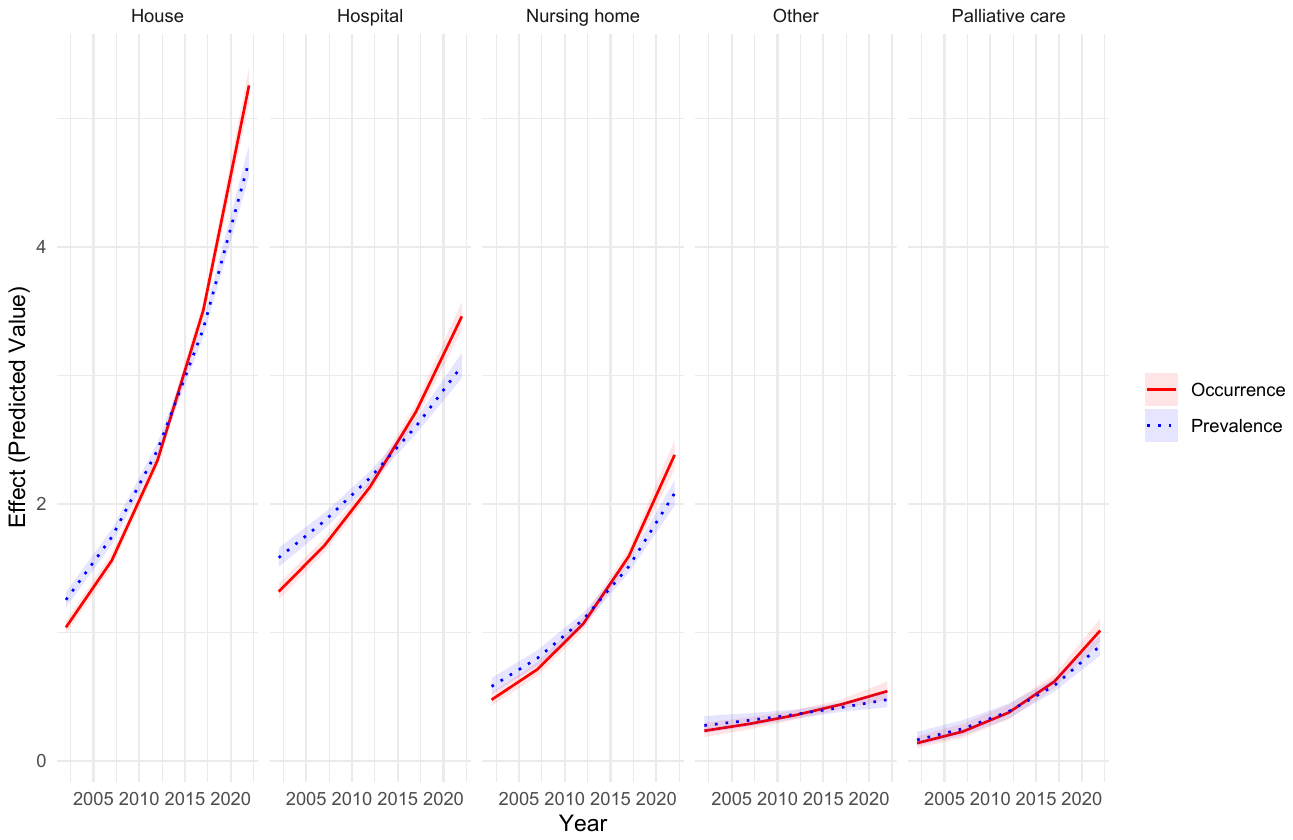

### Supplementary file S5. Marginal effects (year as categorical)

#### Model 1. Marginal effects of year

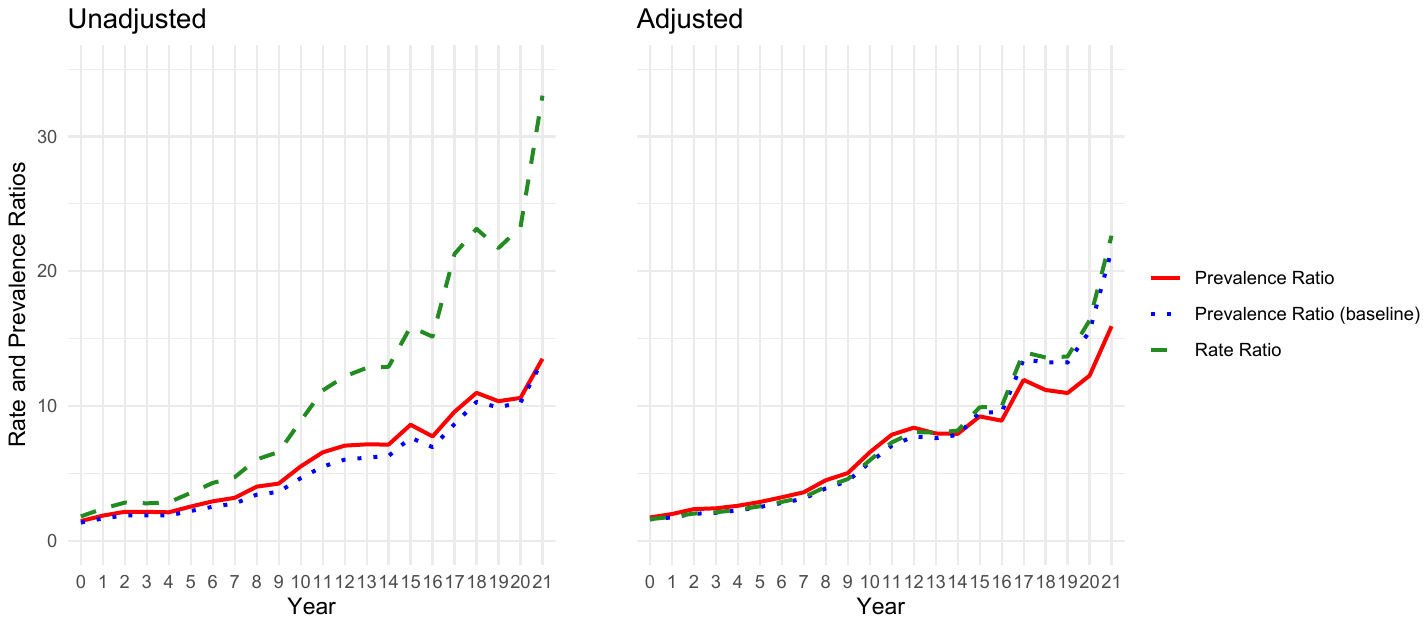

#### Model 2. Marginal effects of the interaction between age group and year

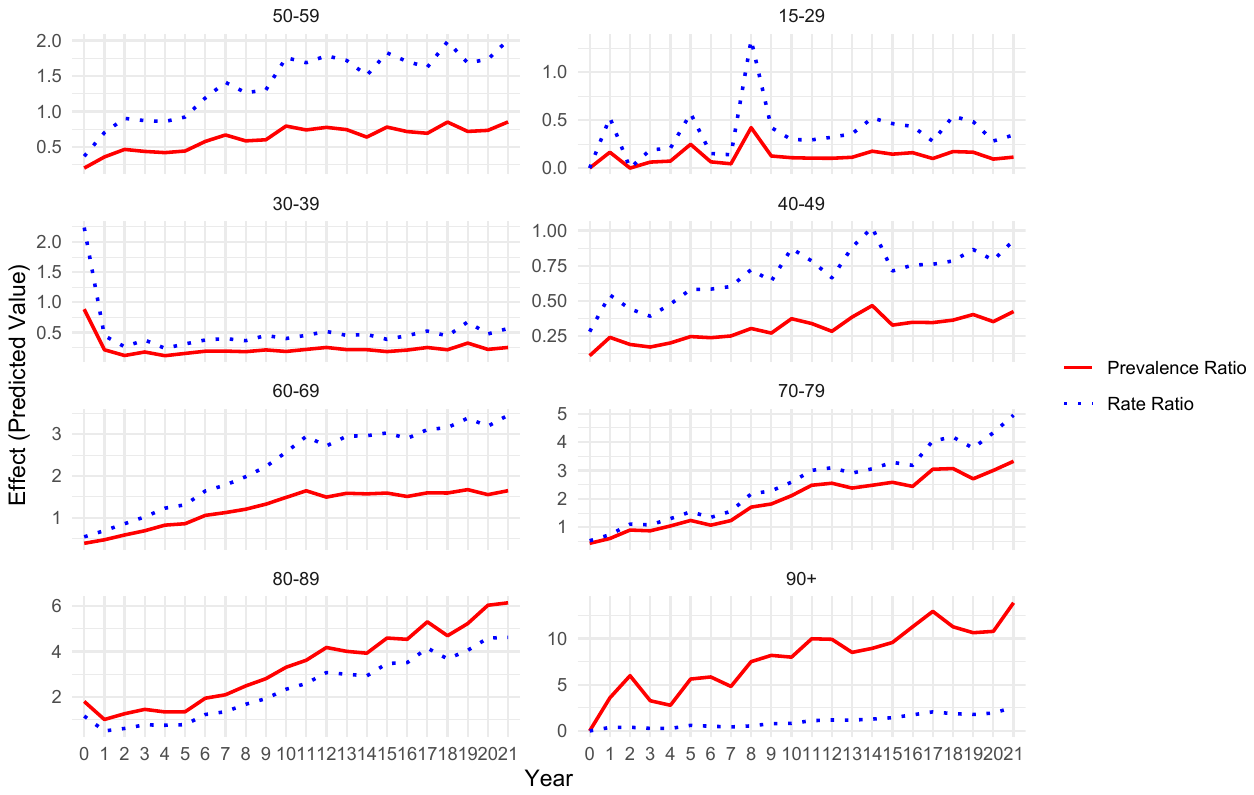

#### Model 3. Marginal effects of the interaction between gender and year

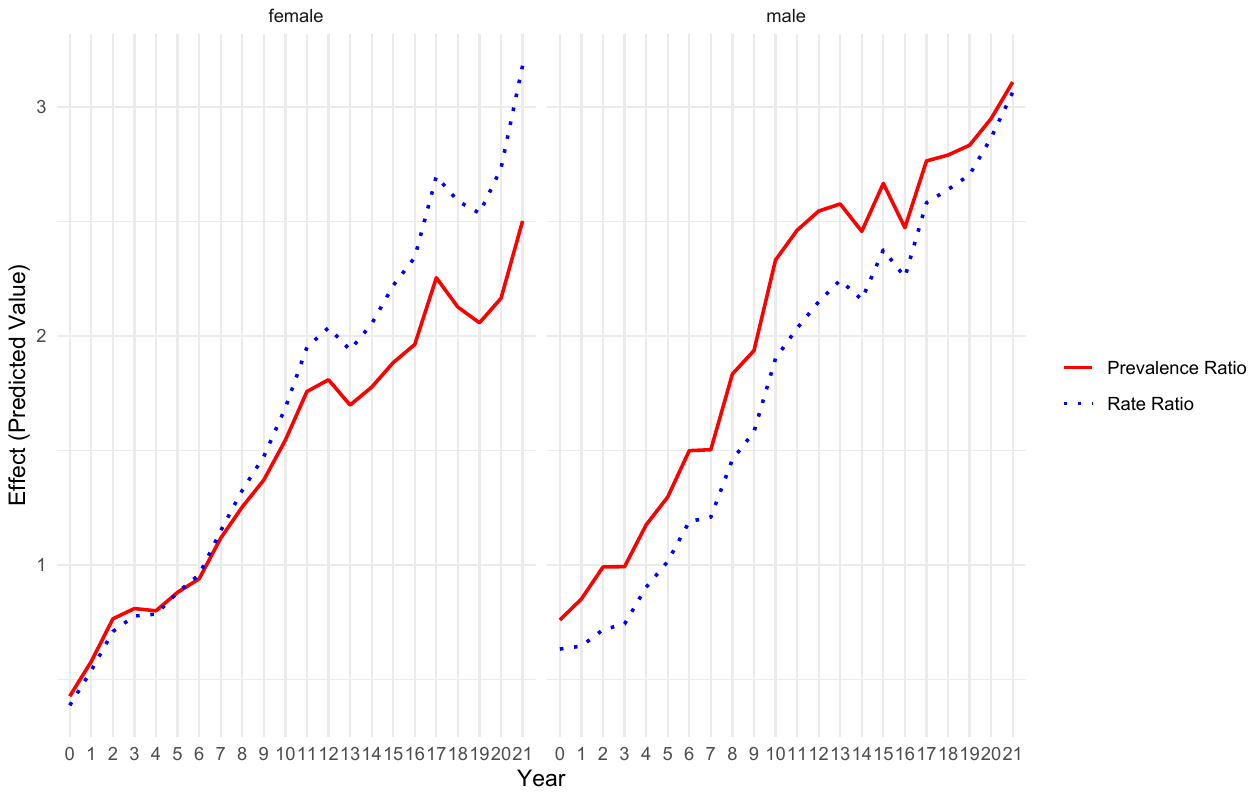

#### Model 4. Marginal effects of the interaction between language/region and year

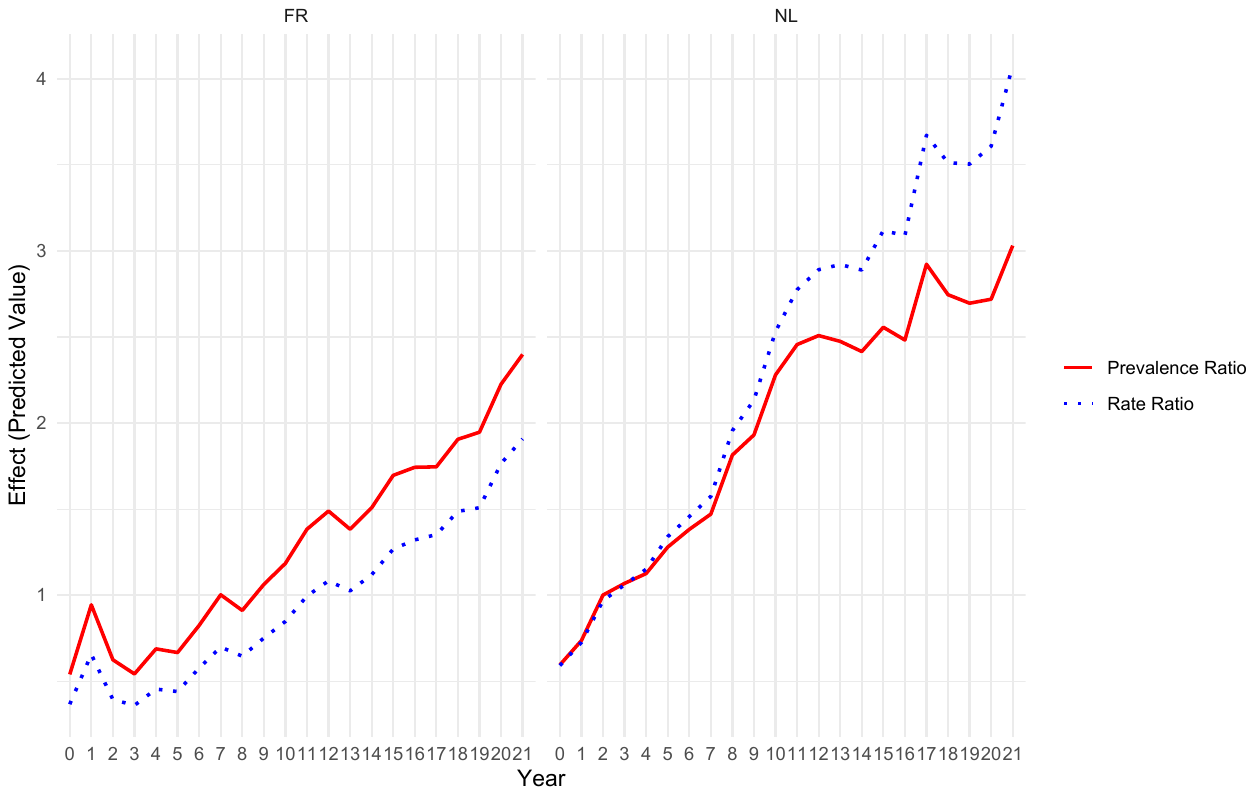

#### Model 5. Marginal effects of the interaction between reason for euthanasia and year

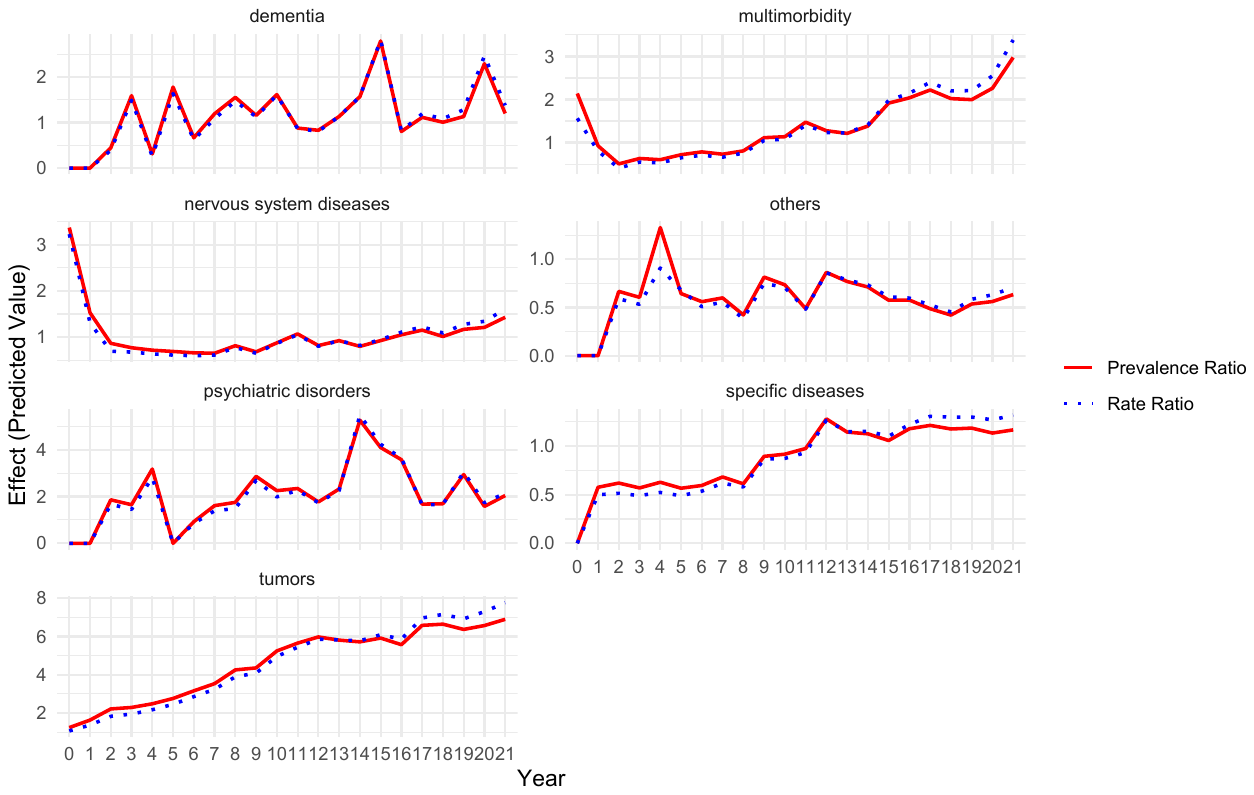

#### Model 6. Marginal effects of the interaction between basis of euthanasia and year

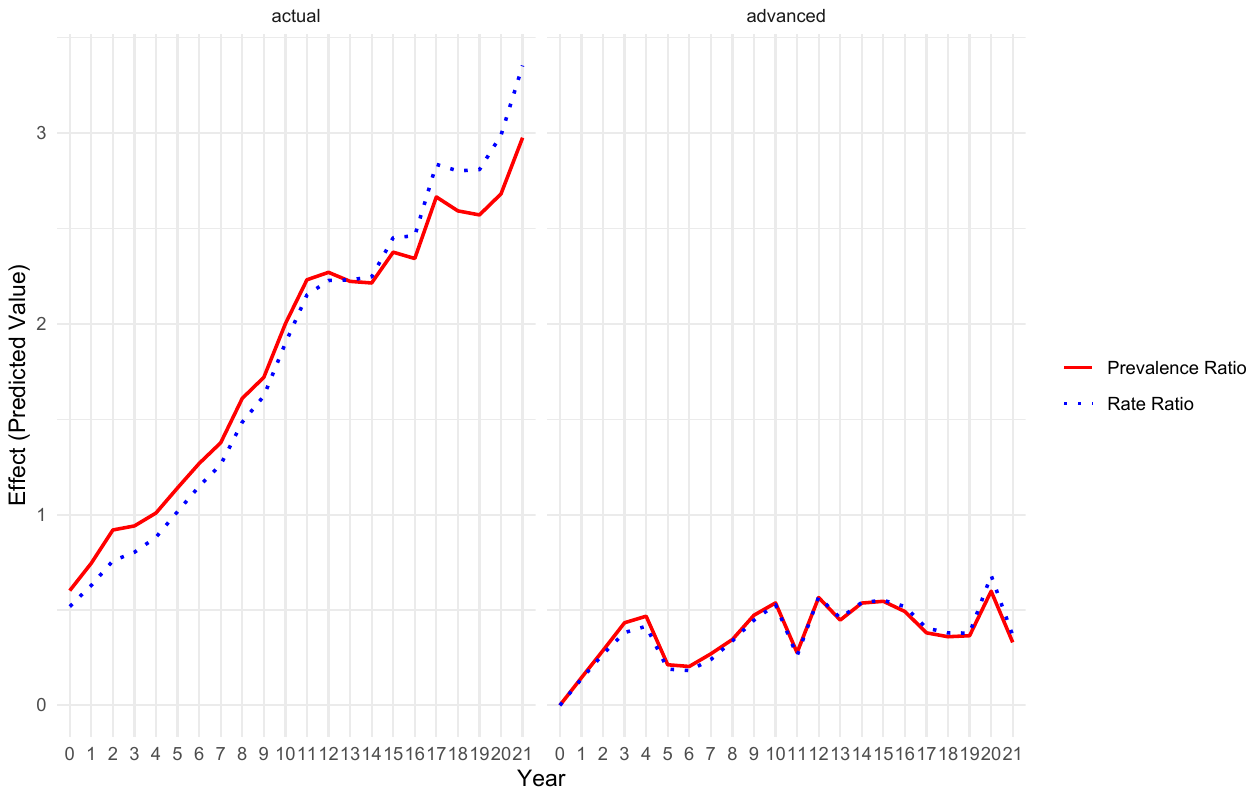

#### Model 7. Marginal effects of the interaction between type of suffering and year

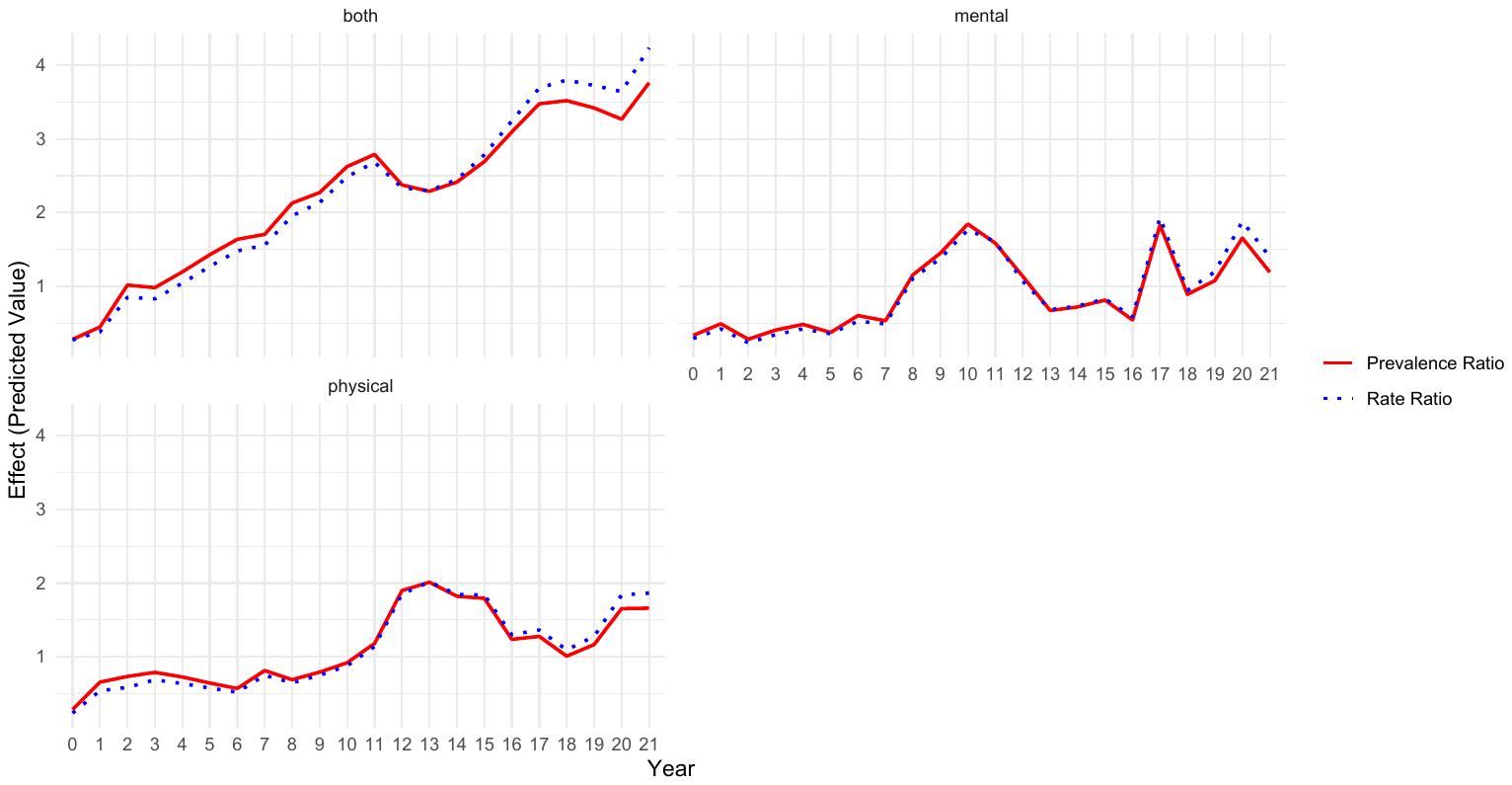

#### Model 8. Marginal effects of the interaction between term of death and year

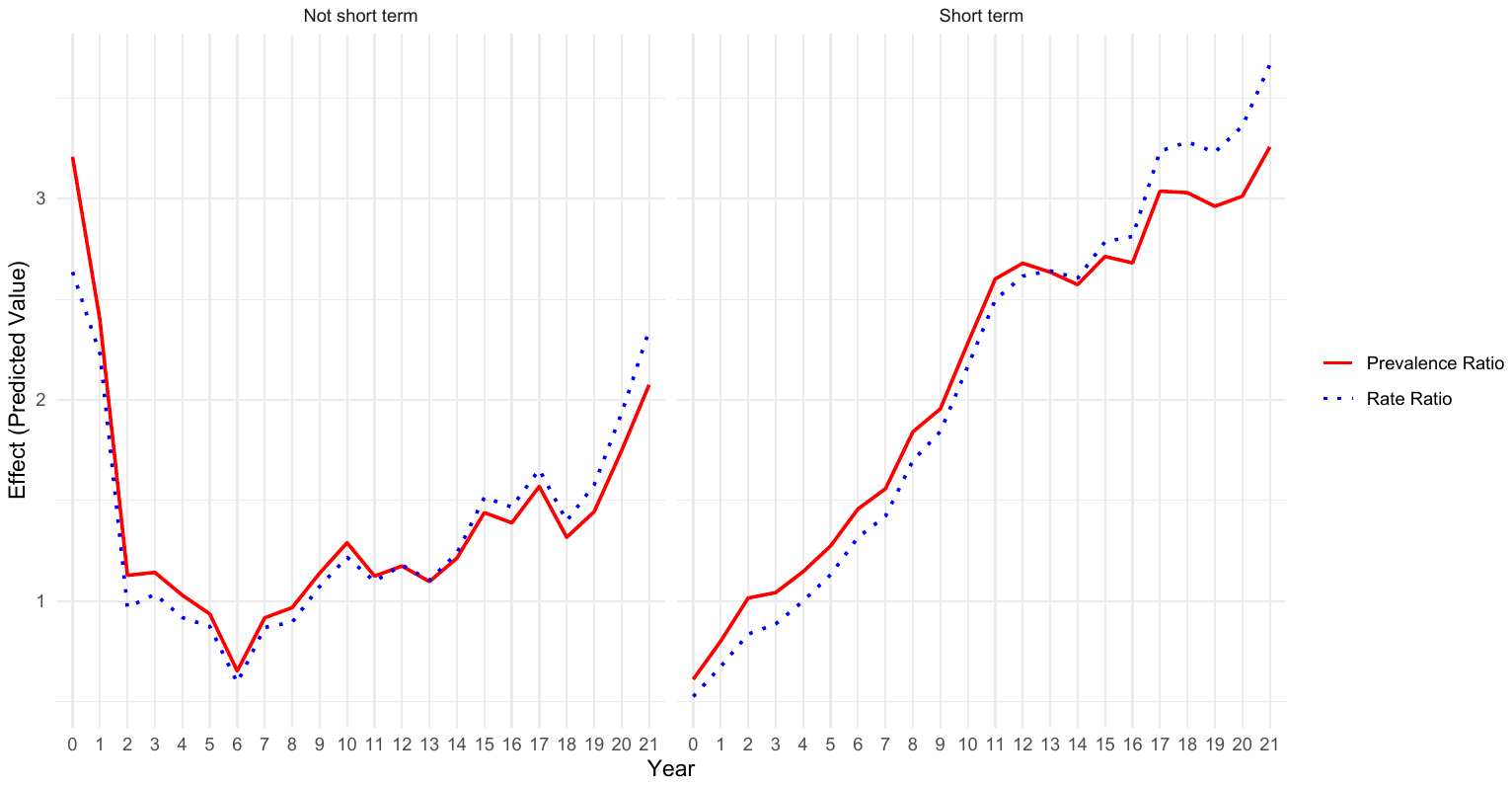

#### Model 9. Marginal effects of the interaction between place of death and year

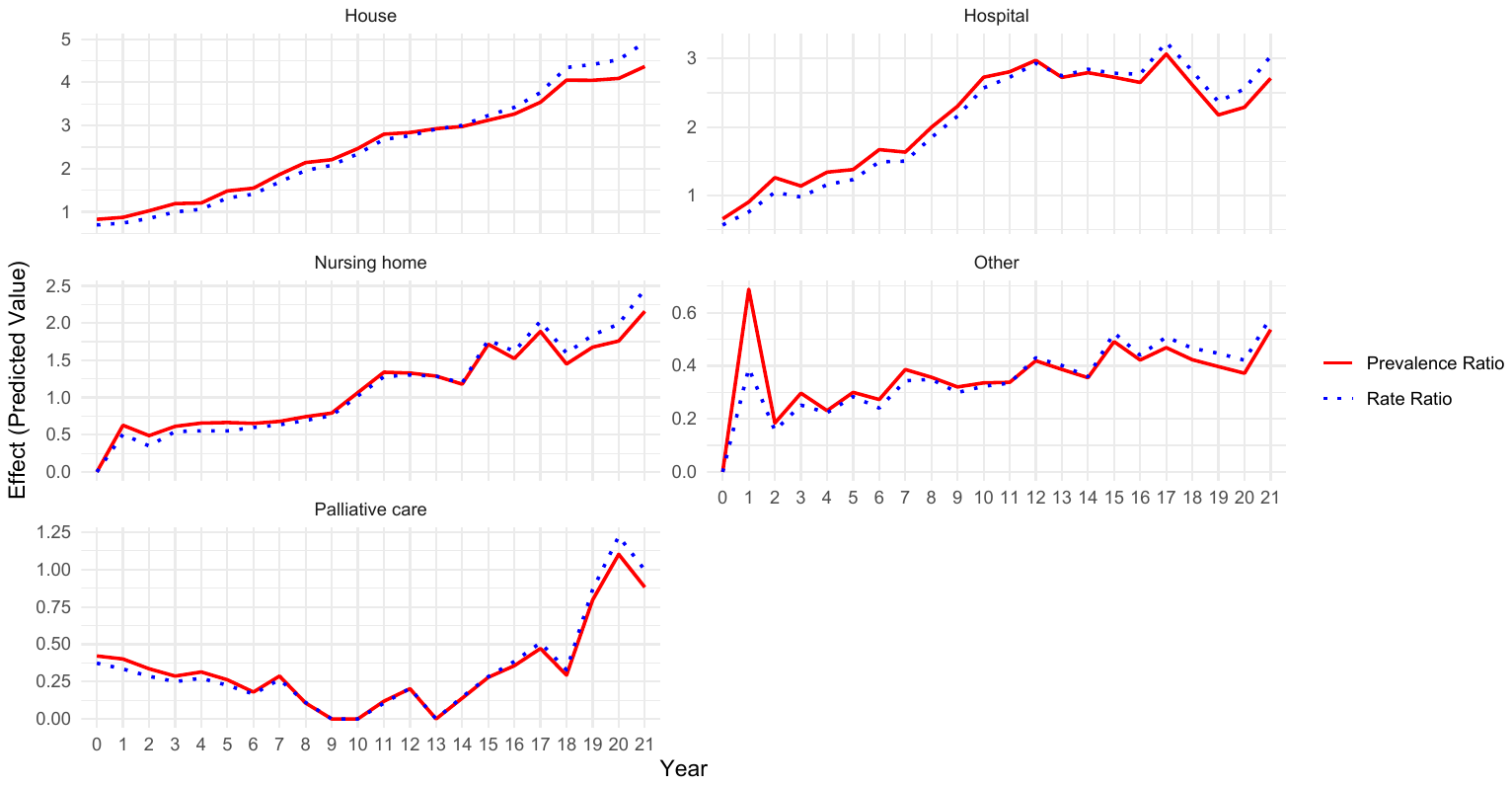

### Supplementary file S6. Linear trends of year by time period (2003-2015; 2016-2022), fully adjusted model without interaction

|  | **2003-2015** | | | | | | |  | **2016-2022** | | | | | | |
| --- | --- | --- | --- | --- | --- | --- | --- | --- | --- | --- | --- | --- | --- | --- | --- |
|  | **Incidence rate ratio** | | |  | **Prevalence ratio** | | |  | **Incidence rate ratio** | | |  | **Prevalence ratio** | | |
|  | RR | 2.5 % | 97.5 % |  | PR | 2.5 % | 97.5 % |  | RR | 2.5 % | 97.5 % |  | PR | 2.5 % | 97.5 % |
| (Intercept) | 0.435 | 0.389 | 0.487 |  | 0.000 | 0.000 | 0.000 |  | 0.781 | 0.686 | 0.889 |  | 0.000 | 0.000 | 0.000 |
| year | **1.118** | **1.112** | **1.124** |  | **1.100** | **1.094** | **1.106** |  | **1.048** | **1.042** | **1.055** |  | **1.033** | **1.027** | **1.039** |
| Age group: 15-29 | 0.198 | 0.148 | 0.259 |  | 0.147 | 0.110 | 0.193 |  | 0.227 | 0.178 | 0.284 |  | 0.182 | 0.142 | 0.228 |
| Age group: 30-39 | 0.294 | 0.254 | 0.339 |  | 0.307 | 0.265 | 0.354 |  | 0.282 | 0.242 | 0.326 |  | 0.310 | 0.266 | 0.359 |
| Age group: 40-49 | 0.488 | 0.444 | 0.536 |  | 0.455 | 0.414 | 0.500 |  | 0.471 | 0.427 | 0.518 |  | 0.508 | 0.461 | 0.559 |
| Age group: 60-69 | 1.501 | 1.413 | 1.595 |  | 1.936 | 1.822 | 2.058 |  | 1.786 | 1.689 | 1.889 |  | 2.114 | 1.999 | 2.237 |
| Age group: 70-79 | 1.564 | 1.474 | 1.659 |  | 2.776 | 2.618 | 2.946 |  | 2.191 | 2.077 | 2.313 |  | 3.779 | 3.582 | 3.990 |
| Age group: 80-89 | 1.368 | 1.288 | 1.454 |  | 4.447 | 4.184 | 4.728 |  | 2.196 | 2.082 | 2.319 |  | 6.733 | 6.381 | 7.109 |
| Age group: 90+ | 0.563 | 0.515 | 0.614 |  | 11.545 | 10.565 | 12.606 |  | 1.033 | 0.970 | 1.099 |  | 14.654 | 13.765 | 15.605 |
| Gender: male | 1.084 | 1.047 | 1.123 |  | 1.398 | 1.349 | 1.448 |  | 1.016 | 0.989 | 1.044 |  | 1.333 | 1.296 | 1.370 |
| Language: NL | 2.773 | 2.651 | 2.902 |  | 1.770 | 1.692 | 1.852 |  | 2.339 | 2.267 | 2.414 |  | 1.410 | 1.367 | 1.455 |
| Reason: Dementia | 0.222 | 0.181 | 0.270 |  | 0.220 | 0.179 | 0.267 |  | 0.196 | 0.169 | 0.226 |  | 0.197 | 0.170 | 0.226 |
| Reason: Multimorbidity | 0.246 | 0.232 | 0.262 |  | 0.242 | 0.227 | 0.257 |  | 0.331 | 0.318 | 0.343 |  | 0.330 | 0.317 | 0.342 |
| Reason: Nervous system diseases | 0.186 | 0.174 | 0.200 |  | 0.186 | 0.173 | 0.199 |  | 0.174 | 0.165 | 0.183 |  | 0.173 | 0.164 | 0.182 |
| Reason: Others | 0.151 | 0.127 | 0.177 |  | 0.147 | 0.124 | 0.173 |  | 0.088 | 0.077 | 0.100 |  | 0.087 | 0.076 | 0.099 |
| Reason: Psychiatric disorders | 0.368 | 0.314 | 0.429 |  | 0.384 | 0.328 | 0.448 |  | 0.364 | 0.316 | 0.418 |  | 0.382 | 0.332 | 0.438 |
| Reason: Specific diseases | 0.204 | 0.191 | 0.218 |  | 0.201 | 0.188 | 0.215 |  | 0.179 | 0.171 | 0.189 |  | 0.179 | 0.170 | 0.188 |
| Basis: advanced | 0.201 | 0.178 | 0.227 |  | 0.201 | 0.177 | 0.226 |  | 0.182 | 0.156 | 0.211 |  | 0.184 | 0.158 | 0.213 |
| Suffering: both | 1.835 | 1.761 | 1.912 |  | 1.837 | 1.763 | 1.914 |  | 2.179 | 2.107 | 2.253 |  | 2.177 | 2.105 | 2.252 |
| Suffering: mental | 0.649 | 0.585 | 0.719 |  | 0.645 | 0.581 | 0.714 |  | 0.597 | 0.542 | 0.657 |  | 0.587 | 0.532 | 0.645 |
| Term: Short term | 1.939 | 1.824 | 2.063 |  | 1.960 | 1.844 | 2.085 |  | 1.938 | 1.862 | 2.018 |  | 1.943 | 1.866 | 2.023 |
| Place: Hospital | 1.014 | 0.977 | 1.053 |  | 1.009 | 0.972 | 1.047 |  | 0.708 | 0.686 | 0.730 |  | 0.709 | 0.687 | 0.732 |
| Place: Nursing home | 0.477 | 0.448 | 0.509 |  | 0.466 | 0.437 | 0.497 |  | 0.443 | 0.425 | 0.461 |  | 0.442 | 0.424 | 0.460 |
| Place: Other | 0.154 | 0.135 | 0.176 |  | 0.150 | 0.131 | 0.172 |  | 0.117 | 0.106 | 0.129 |  | 0.116 | 0.104 | 0.128 |
| Place: Palliative care | 0.188 | 0.145 | 0.239 |  | 0.187 | 0.144 | 0.237 |  | 0.192 | 0.178 | 0.207 |  | 0.191 | 0.177 | 0.206 |
